# A national-scale digital atlas of recorded care patterns across more than 1000 diagnostic categories using real-world health data: a retrospective observational study

**DOI:** 10.64898/2026.09.16.26363186

**Authors:** Markus Haug, Kerli Mooses, Marek Oja, Silver Heinsar, Eno-Martin Lotman, Mariliis Põld, Priit Pauklin, Laura Lõo, Pilvi Ilves, Tuuli Ruus, Gerhard Grents, Anneli Uusküla, Mihkel Arrak, Sulev Reisberg, Jaak Vilo, Raivo Kolde

## Abstract

**Background:** Routinely collected health data can reveal recorded care around diagnosis, but disease-specific curation does not scale across thousands of conditions. We scaled a previously developed automated framework that reduces heterogeneous clinical events into standardised, diagnosis-centred summaries of real-world care patterns.

**Methods:** In this retrospective observational study, we used EST-Health-30, a pseudonymised 30% random sample of Estonian residents with health-care records from 2012 to 2024. We constructed cohorts for ICD-10 three-character diagnostic categories using the first eligible observed diagnosis after a 3-year diagnosis-free observable lookback. For each category, we summarised recorded clinical events from a 90-day pre-index window, a 30-day post-index window, and a 365-day post-index window. An enrichment-based workflow compared earlier self-comparator periods and matched population controls, then filtered and aggregated concepts across clinical domains. Diagnosis-concept relationships were assessed by rubric-guided language-model classification, and atlas plausibility and utility by ten experts.

**Findings:** Of 1645 observed diagnostic categories, 1080 met inclusion criteria, yielding 3240 diagnosis-window summaries across 509,856 individuals. Each summary characterised diagnosis-associated events by prevalence, enrichment, timing, co-occurrence, and variation between patient groups after reducing candidate concepts by 97–98%. Among 296,303 retained diagnosis–concept pairs, the language model classified 24% as directly related, 55% as indirectly related, and 21% as noisy. Experts rated the atlas highly for generating research questions (median 6·5 of 7 [IQR 6–7]), providing information difficult to obtain conventionally (6 [5–7]), and supporting observational study design (5·5 [5–6]).

**Interpretation:** The atlas makes multidimensional diagnosis-centred patterns in longitudinal health records inspectable at population scale in order to support clinical orientation, hypothesis generation, and observational study planning. Outputs represent recorded care rather than individual patient trajectories or causal effects.

**Funding:** Estonian Research Council; European Union; Estonian Ministry of Education and Research; Innovative Medicines Initiative 2 Joint Undertaking; EFPIA.

**Research in context:** *Evidence before this study:* We searched PubMed and medRxiv from database inception to Aug, 2026, without language restrictions, using terms for electronic health records, real-world data, disease trajectories, treatment and care pathways, treatment patterns, clinical-event analysis, OMOP, and population-scale atlases. We also examined relevant reviews and reference lists. Previous population-scale studies have mapped disease trajectories, treatment pathways, or care intensity, generally focusing on diagnoses alone, prespecified events, selected disease cohorts, or a single summary measure of care. OMOP-based treatment-pattern studies have likewise concentrated on specific disease groups. We found no previous population-scale resource that automatically reduced multidomain clinical events surrounding an index diagnosis across more than 1000 diagnostic categories while retaining prevalence, enrichment, timing, co-occurrence, and patient-level heterogeneity.

*Added value of this study:* We applied a previously clinically benchmarked concept-reduction workflow to longitudinal OMOP-mapped health data across 1080 ICD-10 diagnostic categories and three observation windows, producing 3240 diagnosis-window summaries from 509,856 individuals. The workflow reduced candidate clinical concepts by 97–98% and generated compact summaries spanning diagnoses, procedures, medications, measurements, observations, visits, and deaths. The resulting interactive atlas combines concept prevalence and enrichment with timing, co-occurrence, and exploratory patient clustering. An internal formative expert evaluation supported its use for generating research questions and planning observational studies.

*Implications of all the available evidence:* Automated reduction of high-dimensional health-record data can provide a scalable starting point for clinical orientation, hypothesis generation, and observational study design without requiring a separately curated event set for every disease. Applying the same workflow to other OMOP databases could enable direct comparison of diagnosis-centred treatment and care patterns between regions and health systems, while helping distinguish differences in clinical practice from differences in data capture. The atlas is exploratory and should not be interpreted as evidence of causal effects, guideline adherence, or individual patient trajectories.

## INTRODUCTION

The rapid growth of routinely collected health data has created new opportunities to characterise care at scale.^1^ Yet for a given diagnostic group, it remains difficult to determine which treatments, investigations, comorbid events, and patterns of health-care use occur around diagnosis, when they occur, and how they vary between patients.^2^ These records span heterogeneous and overlapping diagnoses, procedures, measurements, observations, medications, and visits.^2–4^ Making such data clinically interpretable typically requires disease-specific code lists and expert-defined events, an approach that is labour-intensive and difficult to standardise across hundreds or thousands of conditions.

Population-scale resources have shown the value of standardised, queryable summaries from routine health data.^5–9^ The Danish Disease Trajectory Browser, for example, maps enriched directional diagnosis pairs at population scale,^5,8^ while Observational Health Data Sciences and Informatics (OHDSI) studies have characterised treatment pathways for selected diseases.^10,11^ These approaches address important but different questions: disease trajectories focus mainly on how diagnoses follow one another, whereas treatment-pathway analyses generally begin with prespecified cohorts and events. Neither is designed primarily to provide a diagnosis-wide view of the broader recorded care surrounding an index diagnosis across multiple clinical domains.

We previously developed CohortContrast, an enrichment-based workflow for identifying diagnosis- and time-window-relevant concepts in Observational Medical Outcomes Partnership (OMOP) mapped real-world data.^12^ The method contrasts events around an index diagnosis with both a pre-index self-comparator period and matched population controls, then reduces non-specific and redundant concepts through filtering and aggregation. For each identified event it reports their prevalence, enrichment, timing relative to index, co-occurrence, and variation between patient groups. The identification of the relevant events and their characterisation within the cohort is driven by the real world patterns in the data and does not require additional expert input.

In this study, we apply the framework at national scale across ICD-10 three-character categories and three prespecified observation windows in OMOP-mapped health data.^13–15^ For each diagnosis-window combination, we identify and summarise diagnosis-associated concepts across procedures, medications, measurements, observations, visits, and diagnoses, and characterise their temporal alignment, co-occurrence, and cluster-level variation. The resulting interactive atlas provides a reusable resource for characterising diagnosis-associated care, exploring patient subgroups, generating hypotheses, and informing observational study design, together with a deployable framework for other OMOP-mapped data resources.

## METHODS

### Data source

We used EST-Health-30, a pseudonymised 30% random sample of Estonian residents with recorded health-care use. The dataset links individual-level health-care provision claims, health insurance coverage, prescription, national health information system, mortality, and cancer registry records, harmonised to the OHDSI OMOP Common Data Model (CDM) using the published Estonian transformation methodology.^16^ The study population included 509,856 individuals with records available from Jan 1, 2012, to Dec 31, 2024.^17^

### Study design and atlas construction

For each ICD-10 three-character category, the index date was defined as the first eligible diagnosis recorded in outpatient or inpatient care after a three-year diagnosis-free lookback without a recorded diagnosis in the same ICD-10 three-character category; this date was defined as the index date. Individuals could contribute to multiple diagnosis cohorts, but only once per diagnosis category. Eligible categories required at least 100 individuals with three years observable lookback and, post-index, at least 365 days observable follow-up unless death occurred earlier. We analysed three prespecified windows relative to index: PRE90 [−90, 0], POST30 [0, +30], and POST365 [0, +365] days, with day 0 included in all windows. Follow-up ended at window end or death, whichever occurred first. Event timing was expressed as days from the start of each observation window. Thus, for PRE90, day 0 corresponds to 90 days before the index diagnosis and day 90 to the index date; for POST30 and POST365, day 0 corresponds to the index date. We refer to each diagnosis category evaluated within one prespecified observation window as a diagnosis-window summary, representing a disease-level care signature derived from longitudinal OMOP records. For each diagnosis-window combination, we generated one concept-reduced summary and assembled these summaries into an interactive atlas (Figure 1).

**Figure 1.**
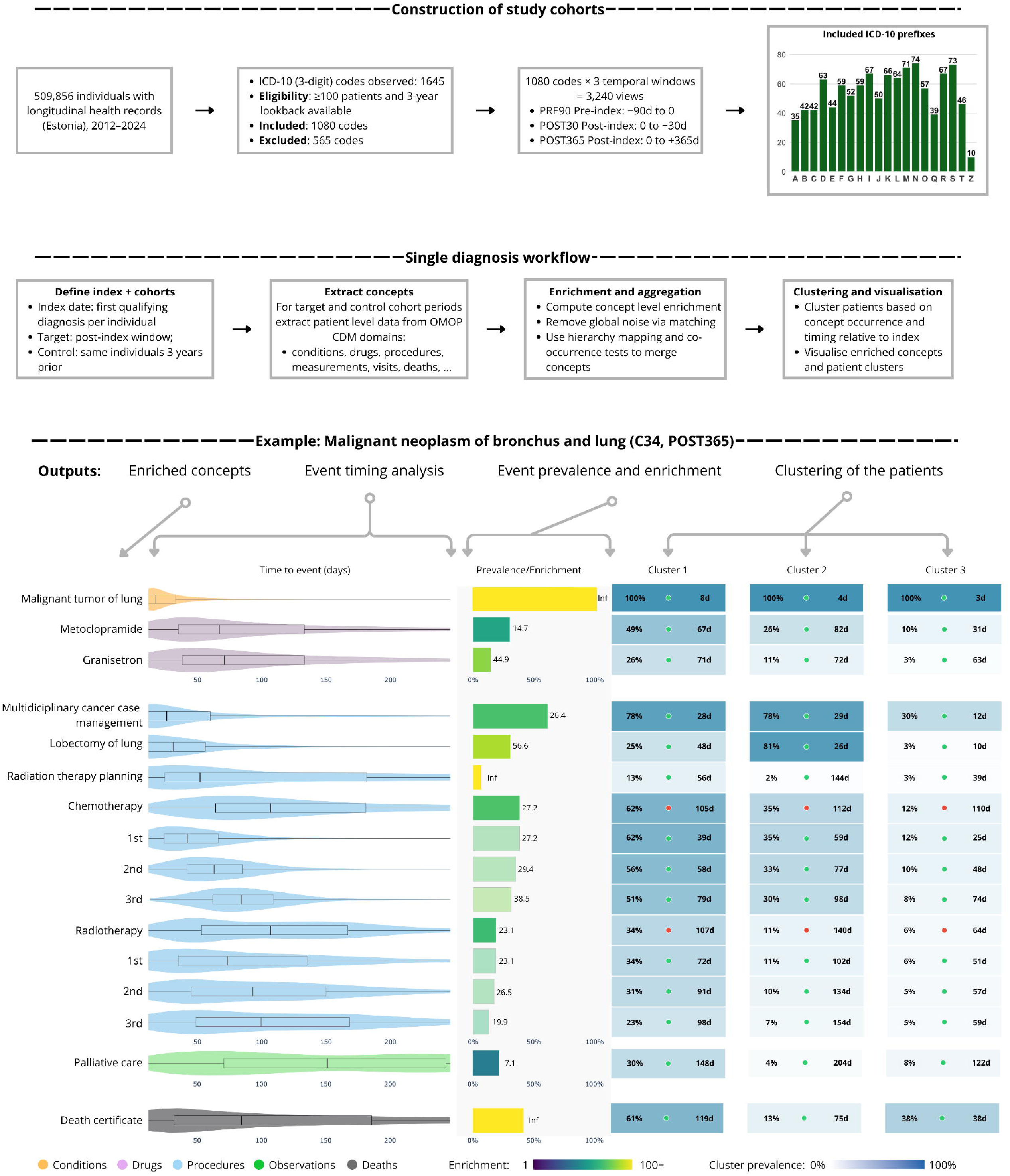
Overview workflow, per-diagnosis processing, and example output (ICD-10 C34, malignant neoplasm of lung).

### Concept summarisation and atlas generation

We applied the previously described CohortContrast concept-reduction workflow^12^ to each eligible diagnosis–window combination. Patient-level recorded events were extracted from the major OMOP clinical domains and represented as standardised clinical concepts. For each clinical concept, enrichment was expressed as the risk ratio of patient-level concept prevalence in the target window to its prevalence among the same patients in an equal-length self-comparator window. The self-comparator window was shifted 1,095 days earlier and aligned to the latest qualifying visit on or before the shifted anchor. Concept prevalence was defined as the proportion of patients with at least one recorded occurrence of the concept during the specified window. Concepts were retained when target prevalence exceeded comparator prevalence and either a Yates-corrected two-proportion χ² test or univariable logistic-regression Wald test met a Bonferroni-adjusted α of 0·05. Temporal background concepts were then removed by applying the same criteria against 1:1 age- and sex-matched population controls assigned the corresponding target calendar interval and observable throughout it. Redundancy was reduced by mapping less-prevalent descendants to the nearest retained vocabulary ancestor and greedily merging within-domain pairs with patient-level presence correlation ≥0·70 and median temporal separation ≤0 days. Repeated events were additionally represented by first, second, and third occurrences. Retained concepts supported precomputed summaries of frequency, enrichment, timing, co-occurrence, demographics, and cluster-level patterns served through the interactive atlas.^18^

Pairwise co-occurrence was calculated from patient-level binary concept-presence indicators. We reported the phi coefficient, equivalent to Pearson correlation for two binary variables and ranging from −1 to 1, and the overlap coefficient, n(A∩B)/min[n(A),n(B)], ranging from 0 to 1. Phi measures association relative to the concepts’ marginal prevalences, whereas overlap measures containment of the less-prevalent concept within the more-prevalent concept.

Exploratory patient clustering was performed within each diagnosis-window summary using all of the retained concepts, represented by whether each retained concept was present, the timing of its first occurrence relative to the index date, and log-transformed occurrence count.^12^ Absent events were coded as zero, features were standardised, and dimensionality was reduced to 20 principal components. Clustering was performed using k-medoids, candidate solutions with k=2–5 were evaluated. For the exploratory cross-disease analysis, each diagnosis–window view was represented using retained-concept presence and frequency, together with cohort-level median age and male proportion. Features were standardised and embedded into two dimensions using Uniform Manifold Approximation and Projection (UMAP, n_neighbors = 30, min_dist = 0·5, metric = “cosine”, random seed = 2025).^19^ ICD-10 subchapter coherence was assessed using silhouette widths.^20^ All publicly reported aggregate results were subject to small-cell disclosure control with k=5, with counts of 1–5 reported as 5.

### Validation

To evaluate the clinical plausibility of retained concepts at atlas scale, we applied the direct–indirect–noisy relationship rubric developed and clinically benchmarked in our preceding methods study.^12^ In that study, agreement between medical-doctor validators and the language-model classifier was comparable to inter-doctor agreement, supporting its use for scalable assessment of diagnosis-concept plausibility. Direct relationships were defined as concepts closely tied to the indexed diagnosis, indirect relationships as clinically plausible but less diagnosis-specific concepts, and noisy relationships as concepts without a meaningful judged relationship to the index diagnosis. Because full manual adjudication of all retained diagnosis–concept pairs was infeasible, we classified the pairs using Gemini 3 Pro Preview with zero-shot prompting (Appendix 1) and temperature 0.^21^ These classifications were used to assess broad clinical plausibility of the atlas outputs, not as gold-standard adjudication of individual pairs, and not as an additional filtering step.

To assess expert-perceived plausibility and utility, ten domain experts each reviewed three atlas views. They rated clinical plausibility, usefulness for research planning, and ease of interpretation on 7-point Likert-type scales, followed by overall ratings of clinical orientation, research question generation, study-design support, and information value. Free-text feedback covered useful insights, research applications, and potentially misleading outputs. Ratings were summarised using medians and IQRs.^22^ (Appendix 2)

## RESULTS

### Atlas coverage and dimensionality reduction

We applied the published concept-reduction workflow to each eligible ICD-10 three-character diagnosis across the three prespecified windows: PRE90, POST30, and POST365.^12^ Among 1,645 observed ICD-10 three-character categories, 1,080 met inclusion criteria, yielding 3,240 diagnosis-window summaries across 509,856 individuals. Cohort sizes were right-skewed, with median cohort sizes of 1,477 in PRE90, 1,518 in POST30, and 1,518 in POST365 (Appendix 3).

Across windows, the workflow reduced high-dimensional candidate event spaces by approximately 97–98%, reducing average candidate concept counts from 2,770, 2,188, and 4,120 per diagnosis-window summary to 76·4, 94·7, and 103·3 retained concepts in PRE90, POST30, and POST365, respectively; median retained concept counts were 59, 71·5, and 75 (Appendix 3). This compaction converted heterogeneous recorded events into interpretable diagnosis-centred care summaries while preserving information on timing, prevalence, enrichment, and cluster structure.

Concept aggregation also contributed to compactness and interpretability. A median of 10 concepts were merged per diagnosis-window summary: 8 in PRE90, 12 in POST30, and 10 in POST365. Hierarchy-based mapping occurred in 90·3% of views and primarily reduced vocabulary granularity, whereas co-occurrence-based mapping occurred in 85·9% and mainly combined concepts reflecting the same care episode or documentation workflow (Appendix 4).

### Clinical plausibility of retained diagnosis–concept relationships

We classified 296,303 retained diagnosis–concept pairs using the direct, indirect, and noisy relationship rubric. Across all windows, 70,952 pairs (24·0%) were classified as direct, 163,016 (55·0%) as indirect, and 62,335 (21·0%) as noisy (Table 1). Direct relationships were most common in PRE90 (34·2%), whereas indirect relationships dominated POST30 and POST365, accounting for 58·8% and 58·0% of pairs, respectively (Appendix 5).

**Table 1.** Rubric-based classification of retained diagnosis–concept relationships by observation window.

| Window | Direct |  | Indirect |  | Noise |  |
| --- | --- | --- | --- | --- | --- | --- |
| POST365 | 21894 | 19·6% | 64722 | 58·0% | 24904 | 22·3% |
| POST30 | 20868 | 20·4% | 60111 | 58·8% | 21305 | 20·8% |
| PRE90 | 28190 | 34·2% | 38183 | 46·3% | 16126 | 19·5% |
| Total | 70952 | 24·0% | 163016 | 55·0% | 62335 | 21·0% |

Distributional summaries supported this interpretation: across windows, direct concepts showed higher median enrichment than noisy concepts, including in POST30, where median enrichment was 19·37 for direct concepts versus 5·10 for noisy concepts (Appendix 6). The higher proportion of direct concepts in PRE90 likely reflects concentration of diagnostic workup near first recorded diagnosis, whereas post-index windows capture follow-up care, monitoring, comorbidity management, and general health-care utilisation.

### Illustrative diagnosis-window summary

Cerebral infarction illustrates a clear transition from acute stroke management before diagnosis to rehabilitation-oriented and complication-related care during follow-up (Figure 2). In PRE90, retained concepts were concentrated close to the index diagnosis and reflected an acute stroke diagnostic and treatment pathway. High-variance and high-prevalence concepts included emergency room and inpatient visits, intensive care, non-contrast and contrast-enhanced brain imaging, cerebral angiography or CT angiography, electrocardiographic monitoring, coagulation testing, thrombolytic therapy, and ischaemic stroke. Several of these concepts were highly prevalent, including non-contrast brain CT or radiology of two body areas (84·5%), emergency room and inpatient visits (75·5%), and electrocardiographic monitoring (74·9%). Overall, this profile was consistent with acute stroke care, in which rapid emergency assessment, urgent brain imaging, vascular imaging when thrombectomy is considered, and thrombolysis in eligible patients are central parts of the recorded pathway.^23,24^

**Figure 2.**
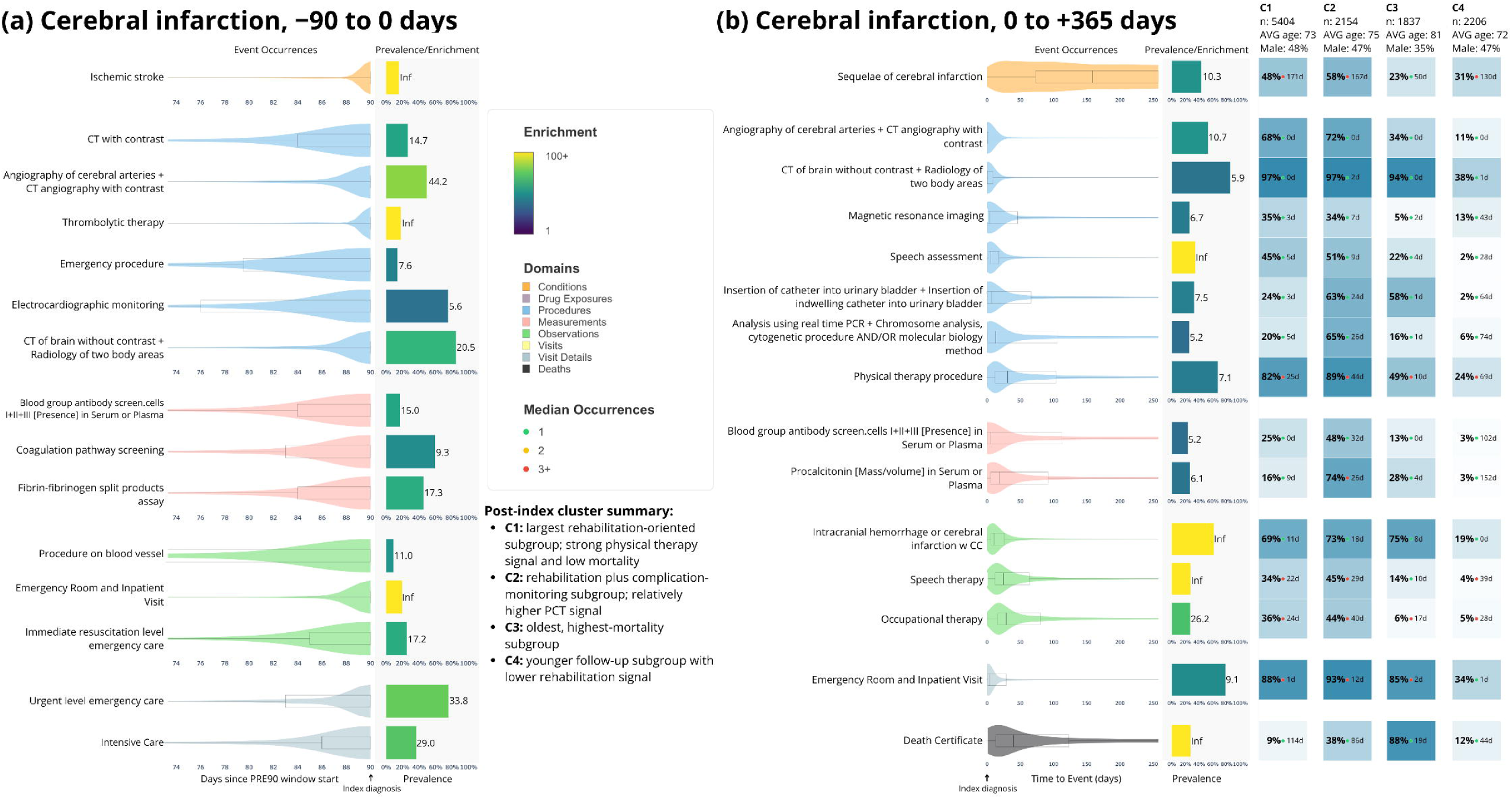
PRE90 (A) and POST365 (B) for Cerebral Infarction (ICD-10 I63). Both panels present the 15 highest between-cluster variance concepts for both observation windows. Event-occurrence plots show concept timing across the observation period. The prevalence/enrichment panel summarises prevalence in the target cohort and enrichment versus controls (Inf indicates no control occurrences). The cluster panel shows cluster-specific prevalence and median occurrence counts.

In POST365, the atlas shifted from acute diagnostic workup to a broader post-stroke care profile (Figure 2). Imaging and acute-care concepts remained frequent, but the retained summaries also captured downstream consequences and rehabilitation, including sequelae of cerebral infarction, physical therapy, speech assessment, speech therapy, and occupational therapy. This broader profile was consistent with stroke rehabilitation guidance recommending multidisciplinary assessment and therapy for post-stroke functional, communication, swallowing, cognitive, and mobility needs.^25^ The co-occurrence of urinary catheterisation, procalcitonin, emergency or inpatient care, and death additionally identified severe or complicated post-stroke courses, including patterns compatible with infection-related deterioration and higher mortality.^26^

Cluster-level summaries further separated cerebral infarction patients along axes of rehabilitation intensity, complication monitoring, and mortality (Figure 2). C1 and C2 represented broadly similar post-stroke profiles, both combining substantial inpatient and imaging-related care with therapies, but C2 showed more complication-associated monitoring and mortality: procalcitonin was present in 74% of C2 versus 16% of C1, and death certificate prevalence was 38% versus 9%, respectively.^26,27^ C3 represented the oldest and highest-mortality subgroup (mean age 81 years), with death certificate recorded in 88% of patients and a median time to death of about 19 days after index, together with high prevalence of emergency or inpatient care, urinary catheterisation, and complication-coded cerebral infarction. C4 showed a more mixed and lower-intensity profile, with less rehabilitation and fewer complication-related concepts. Taken together, these clusters suggested that the atlas could distinguish rehabilitation-oriented recovery, complication-associated care, early high-mortality patterns, and lower-intensity follow-up within a single diagnosis-window summary.

In the POST365 cerebral infarction summary, co-occurrence analyses identified distinct acute-care, rehabilitation, and mortality-associated care modules (Figure 3). Brain CT and cerebral angiography or CT angiography were among the earliest recorded procedures and showed notable co-occurrence with each other (phi 0·43; overlap coefficient 1·00), consistent with their role in acute stroke workup. Rehabilitation-related procedures followed later and clustered with the acute-care pathway, whereas death certificate showed its strongest positive association with urinary catheterisation (phi 0·33; overlap coefficient 0·58) and weak or negative associations with several rehabilitation-related procedures. Cluster-specific temporal summaries were consistent with this pattern: acute imaging occurred near diagnosis across patient groups, whereas repeated physical therapy was more prominent in lower-mortality survivor groups and less prominent in the early high-mortality cluster. Together, these results show that a single atlas view can combine event timing, prevalence, co-occurrence, and exploratory patient clustering to reveal clinically interpretable variation in recorded care patterns.

**Figure 3.**
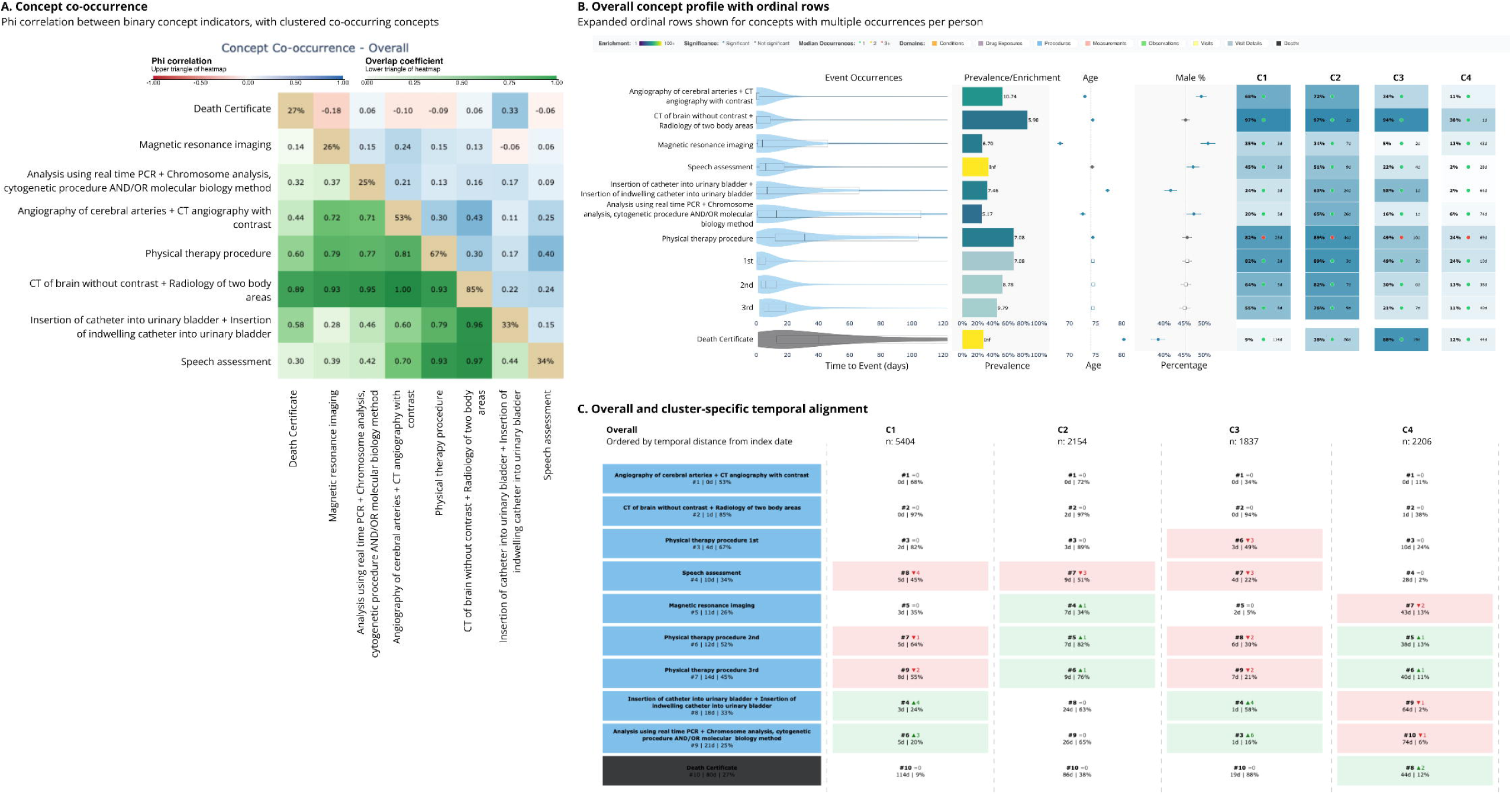
Co-occurrence, overall concept profiles, and cluster-specific temporal alignment for cerebral infarction in POST365. Procedure and death concepts from the POST365 cerebral infarction view are shown to illustrate how the atlas captures care-process structure beyond concept prevalence alone. A, Pairwise concept co-occurrence among binary concept indicators. The upper triangle shows phi correlations, the lower triangle shows overlap coefficients, and the diagonal shows concept prevalence. B, Overall concept profiles from the interactive atlas, including event-occurrence distributions, prevalence and enrichment, cohort age and sex summaries, and cluster-level concept summaries. Ordinal rows show repeated occurrences of the same concept where multiple occurrences per person were retained. C, Overall and cluster-specific temporal alignment of the same selected concepts, ordered by temporal distance from the index diagnosis. Together, the panels show that retained concepts formed interpretable care-process modules, including acute imaging, rehabilitation, and complication or mortality-related care, which were expressed differently across patient clusters.

### Within-disease patient structure

Within diagnosis-window summaries, patient clustering most often selected two groups. The highest silhouette score on average was k=2 in 76·8% of PRE90, 67·5% of POST30, and 80·6% of POST365 views; higher-order solutions were less common. Average best-clustering silhouette scores were modest at 0·224, 0·234, and 0·229, respectively, with the strongest separation in POST30. These findings suggest that within-disease variation was usually organised along one dominant care-intensive axis rather than many sharply separated patient clusters, consistent with the expected complexity of longitudinal clinical data.^2,12^

The concepts most often driving clustering were general markers of health-care utilisation and acute clinical workup, including inpatient and emergency visits, electrocardiographic monitoring, and common laboratory measurements such as electrolytes, glucose, bilirubin, inflammatory markers, and blood counts. The observation windows shared 80% of the most prevalent top ten separators over the ICD-10 codes (Appendix 7). Thus, in many diagnosis-window summaries, unsupervised patient clusters appeared to reflect differences in care intensity, acute severity, and data continuity rather than narrowly disease-specific patient clusters.

### Cross-disease structure

We also examined whether diagnosis-window summaries with similar retained-concept profiles formed coherent disease-space groupings. Overall separation of ICD-10 subchapters was weak across all three windows, indicating that recorded care patterns did not generally reproduce the ICD-10 hierarchy. However, several subchapters formed coherent local groups, including burns and corrosions, disorders of the gallbladder, biliary tract and pancreas, and pregnancy with abortive outcome (Figure 4; Appendix 8).

**Figure 4.**
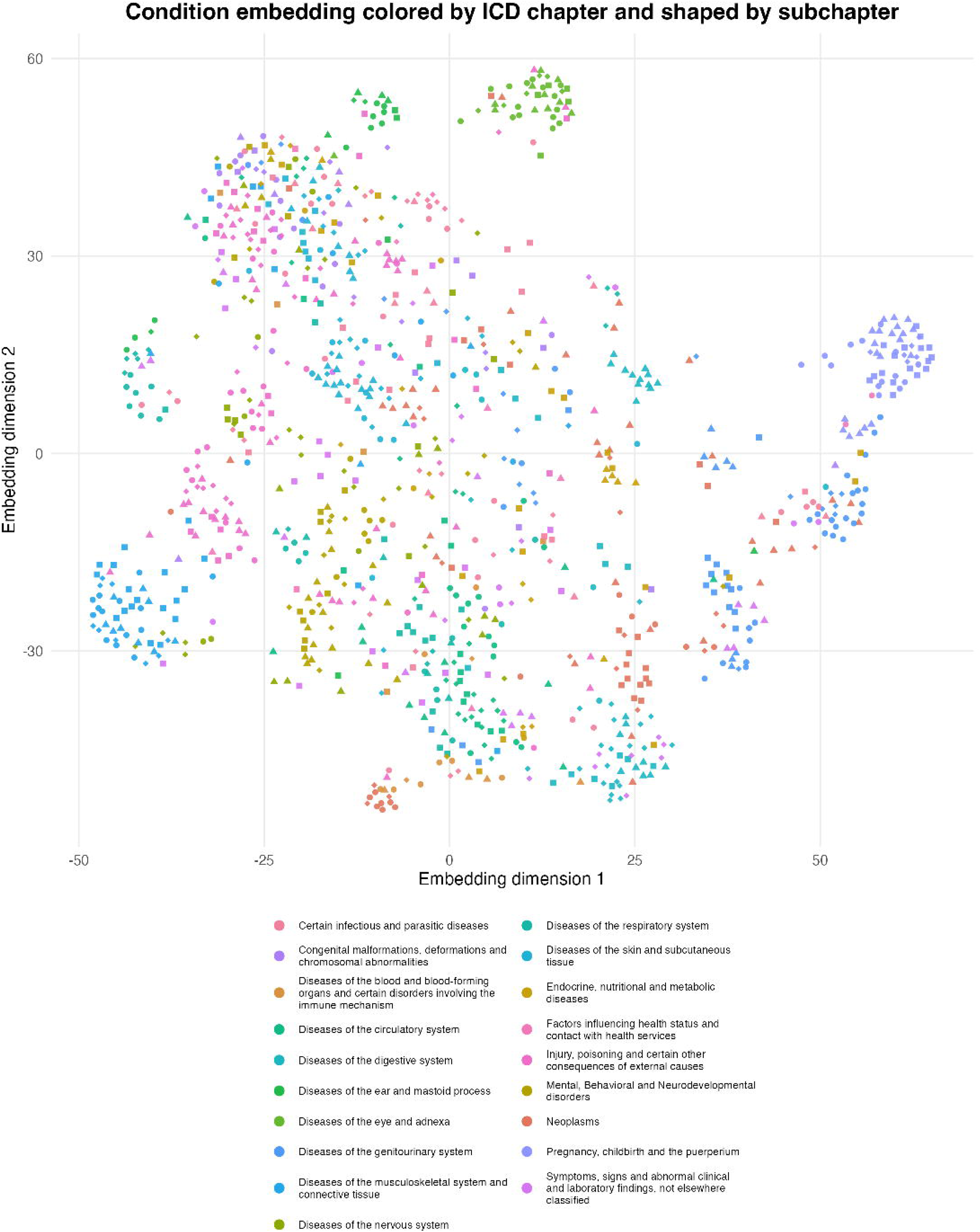
Cross-disease embedding of diagnosis-window summaries by ICD-10 chapter and subchapter for POST365. Each point represents an eligible ICD-10 three-character diagnosis category embedded using retained-concept presence and frequency, together with cohort-level median age and male proportion. Colours indicate ICD-10 chapters and point shapes indicate ICD-10 subchapters. The two-dimensional projection was generated using UMAP for visual exploration of whether diagnoses with similar retained care-pattern summaries formed local groupings. Axis orientation and absolute distances are arbitrary; local proximity should be interpreted as approximate similarity in the atlas-derived feature space.

These outputs form a precomputed interactive atlas across all 3,240 diagnosis-window summaries, including retained concept occurrences, temporal alignment, demographics, co-occurrence, and cluster-level summaries.^18^

In the structured formative evaluation, all ten experts rated the atlas positively for clinical orientation, research-question generation, and providing information difficult to obtain from conventional sources, with every rating at least 5/7. Median ratings were 5·5/7 (IQR 5–7) for clinical orientation, 6·5/7 (IQR 6–7) for research-question generation, 5·5/7 (IQR 5–6) for observational study-design support, and 6/7 (IQR 5–7) for providing otherwise difficult-to-obtain information. Eight of ten respondents also rated its usefulness for study design at least 5/7. Across 30 diagnosis-level reviews, usefulness as a starting point for disease-specific research received a median rating of 5·5/7, whereas ease of interpretation without additional explanation was lower at 5/7. Free-text feedback supported hypothesis generation and cohort planning, while lower ratings sometimes reflected requests for finer ICD-10 cohort definitions or difficulty interpreting atlas-specific timelines, clustering, terminology, and aggregation. (Table 2, Appendix 2)

**Table 2.** Expert evaluation of atlas views using 7-point Likert-type scales.

| Rating item | n | Mean | Median | SD | IQR |
| --- | --- | --- | --- | --- | --- |
| Clinical plausibility | 30 | 5·33 | 5 | 1·60 | 4-7 |
| Useful starting point for disease-specific research | 30 | 5·13 | 5·5 | 1·61 | 4-6·5 |
| Easy to interpret without additional explanation | 30 | 4·50 | 5 | 1·28 | 4-5·5 |
| Overall usefulness for clinical orientation | 10 | 5·80 | 5·5 | 0·92 | 5-7 |
| Overall usefulness for generating research questions | 10 | 6·30 | 6·5 | 0·82 | 6-7 |
| Overall usefulness for refining observational study design | 10 | 5.40 | 5.5 | 0.97 | 5-6 |
| Provides information difficult to obtain conventionally | 10 | 6.00 | 6 | 0.82 | 5-7 |

## DISCUSSION

This study extends the CohortContrast workflow from disease-specific analysis to a diagnosis-wide resource. Unlike disease-trajectory approaches, which characterise relationships between diagnoses over time, and treatment-pathway approaches that generally begin with prespecified cohorts and events, the atlas characterises the broader recorded care surrounding an index diagnosis across multiple clinical domains.^5,8,10–12^ It therefore provides a scalable starting point for characterising diagnosis-associated investigation and treatment, identifying temporally relevant clinical events and patient heterogeneity, and selecting candidate events for subsequent cohort definitions or trajectory modelling.

Routinely collected health data do not contain clinical pathways in a ready-to-analyse form. Events surrounding a diagnosis include disease-specific investigation and treatment alongside monitoring, complications, comorbidity management, follow-up, and health-care contact. The atlas reduces this event space while preserving prevalence, enrichment, timing, mappings, and co-occurrence. Its summaries are intended to structure recorded diagnostic context for clinical review and subsequent study design, not to define a disease or reproduce a guideline pathway.

Reduction and aggregation are necessary for diagnosis-wide analysis, but they introduce an important trade-off. Keeping concepts at their original granularity preserves detail but makes comparison across conditions difficult; broader parent or composite concepts improve readability but can combine distinctions that are clinically important. The expert evaluation illustrated both sides of this problem. Reviewers identified useful care patterns, but also pointed to the fixed three-character ICD granularity, missing source information, and concepts that had been combined too broadly. These observations support the use of automated summarisation as a way to narrow the space requiring review, rather than as a replacement for disease- or specialty-specific curation.

The patient clustering showed how recorded care varied within a diagnosis. Separation was often associated with acute investigation, hospital use, laboratory monitoring, severity-related care, and data continuity rather than narrowly disease-specific interventions. This is consistent with informed-presence effects in electronic health records, whereby available information depends partly on health-care contact.^28,29^ These clusters should therefore not be interpreted as fixed clinical phenotypes, but as indicators of heterogeneity relevant to defining study populations, covariates, exposures, or outcomes.

The expert rating (median 6 of 7) of the atlas as providing information “difficult to obtain conventionally” reflects a real practical barrier. No public source reports prevalence, enrichment, timing, and co-occurrence of recorded clinical events across more than a thousand diagnostic categories at population scale. Obtaining such information for even one diagnosis entails a full epidemiologic study, requiring a separate ethics application, a purpose-built cohort and analysis plan, while dealing with strict data privacy requirements. The atlas does not remove these requirements for downstream studies, but by offering precomputed summaries across the full diagnostic space under k>5 small-cell disclosure control, it makes population-scale patterns publicly inspectable without exposing individual-level data.

The expert evaluation further clarifies the intended role of the atlas. Reviewers generated questions concerning treatment choice and timing, patient subgroups, monitoring, outcomes, and follow-up periods, while also identifying outputs that were difficult to interpret or did not match clinical expectation. In routinely collected data, unexpected patterns may be clinically relevant, but may also arise from coding practice, incomplete data capture, concept mapping, or differences in health-care contact. Making such discrepancies visible before cohort definitions and downstream analytical choices are fixed is invaluable while developing more specific real-world studies on similar data.

The Estonian implementation demonstrates the value of applying a common analysis across linked national data sources while also highlighting the distinction between local findings and a transferable analytical framework. Claims, prescription records, national health information system data, mortality records, and cancer-registry data are represented within the same OMOP framework, allowing researchers to examine recorded care around different diagnoses using a common procedure. However, these care patterns should not be assumed to represent other health systems because coding, access, reimbursement, clinical practice, and source-data coverage differ between settings.

OMOP creates the opportunity to apply the same workflow in other data environments while retaining local governance of patient-level data.^30^ Cross-database comparison could identify which diagnosis-centred patterns are reproducible and which reflect local health systems or data capture, and could test whether proposed cohort definitions, observation windows, covariates, and outcomes behave similarly before larger multi-database studies. External implementation is therefore the next test of the framework and would distinguish transferable features of the atlas from those specific to the Estonian setting.

### Limitations

Because the atlas relies on recorded health-care activity around diagnosis, its results reflect a mixture of underlying disease processes, treatment patterns, and the structure, quality, and completeness of the underlying real-world data. Local coding practices, reimbursement rules, data-source coverage, documentation workflows, health-care access, and patient contact with the health system all influence the results. Therefore, these care patterns should not be assumed to be fully transferable to other health systems, although the standardised OMOP workflow reduces some differences in coding and vocabulary representation.

Concept reduction and aggregation make diagnosis-wide analysis feasible, but can obscure distinctions that are important for specific diseases. The retained atlas outputs were subsequently assessed for broad clinical plausibility using a rubric-guided large language model rather than exhaustive expert adjudication. Model-specific bias and misclassification therefore remain possible in the plausibility classifications themselves.

Finally, the expert evaluation was internal and formative. The ten evaluators subsequently contributed to interpretation and revision of the study and are co-authors; their assessments should therefore not be considered independent external validation.

## CONCLUSION

This study demonstrates that OMOP-mapped real-world health data can be transformed into a scalable, privacy-protected, diagnosis-wide atlas of recorded care patterns. By making diagnostics, treatments, monitoring, complications, utilisation, and follow-up inspectable across more than 1,000 diagnostic categories, the atlas provides a reusable resource for clinicians, researchers, and health-system partners to orient disease-specific research, refine cohort definitions, generate hypotheses, and design follow-up studies.

## Contributors

All authors participated in the revision process and have approved the submitted version. The work reported in the paper has been performed by the authors, unless clearly specified in the text. Conceptualization – Markus Haug, Raivo Kolde. Data curation – Markus Haug, Raivo Kolde, Marek Oja, Kerli Mooses, Sulev Reisberg, Jaak Vilo. Formal Analysis – Markus Haug, Raivo Kolde. Funding acquisition – Raivo Kolde, Jaak Vilo. Methodology – Markus Haug, Raivo Kolde. Project administration – Markus Haug, Raivo Kolde. Resources – Markus Haug, Raivo Kolde. Software – Markus Haug, Raivo Kolde. Supervision – Raivo Kolde. Validation – Markus Haug, Silver Heinsar, Eno-Martin Lotman, Mariliis Põld, Priit Pauklin, Laura Lõo, Pilvi Ilves, Tuuli Ruus, Gerhard Grents, Anneli Uusküla. Visualization – Markus Haug, Raivo Kolde. Writing – original draft – Markus Haug. Writing – All authors.

Markus Haug and Raivo Kolde accessed and verified the underlying data reported in the manuscript.

## Data sharing

There are legal restrictions on sharing de-identified data. In accordance with legislative and data protection requirements in Estonia, the authors cannot publicly release data obtained from the Estonian health data registers. To protect patient privacy, concept-level metrics are rounded up and reported as 5 whenever a concept is present for 1–5 patients, whether that count appears as a stand-alone value or in the numerator or denominator of a derived measure. The atlas is available at http://omop-apps.cloud.ut.ee/CohortContrastAtlas and the code for reproduction on OMOP CDM databases is documented in the GitHub repository https://github.com/HealthInformaticsUT/CohortContrast.

## ETHICS STATEMENT

The study was approved by the Research Ethics Committee of the University of Tartu (300/T-23 and No. 330/T-10) and the Estonian Committee on Bioethics and Human Research (1.1-12/653) and the requirement for informed consent was waived.

## Data Availability

There are legal restrictions on sharing de-identified data. In accordance with legislative and data protection requirements in Estonia, the authors cannot publicly release data obtained from the Estonian health data registers. To protect patient privacy, concept-level metrics are rounded up and reported as 5 whenever a concept is present for 1-5 patients, whether that count appears as a stand-alone value or in the numerator or denominator of a derived measure. The atlas is available at http://omop-apps.cloud.ut.ee/CohortContrastAtlas and the code for reproduction on OMOP CDM databases is documented in the GitHub repository https://github.com/HealthInformaticsUT/CohortContrast.

## Acknowledgments

This work was supported by the Estonian Research Council (PRG1844, PRG2078, PSG1216, PRG2218). The study was funded by the European Union and co-funded by the Ministry of Education and Research (TEM-TA72). The European Union funded the project under its Horizon Europe research and innovation programme (grant agreement No 101060011, TeamPerMed) and co-funded the research through the European Regional Development Fund (Project No. 2021-2027.1.01.24-0444). Views and opinions expressed are however those of the author(s) only and do not necessarily reflect those of the European Union or the European Research Executive Agency. Neither the European Union nor the granting authority can be held responsible for them. This work was also supported by the Estonian Centre of Excellence in Artificial Intelligence (EXAI), funded by the Estonian Ministry of Education and Research grant TK213. This work was further supported by the OPTIMA project (grant agreement No. 101034347) through IMI2 Joint Undertaking supported by European Union’s Horizon 2020 research and innovation programme and the European Federation of Pharmaceutical Industries and Associations (EFPIA).

## Declaration of generative AI and AI-assisted technologies in the manuscript preparation process

During manuscript preparation, the authors used ChatGPT to improve language, readability, and wording. After using this tool, the authors reviewed and edited the content as needed and take full responsibility for the accuracy, integrity, and final version of the publication.

---

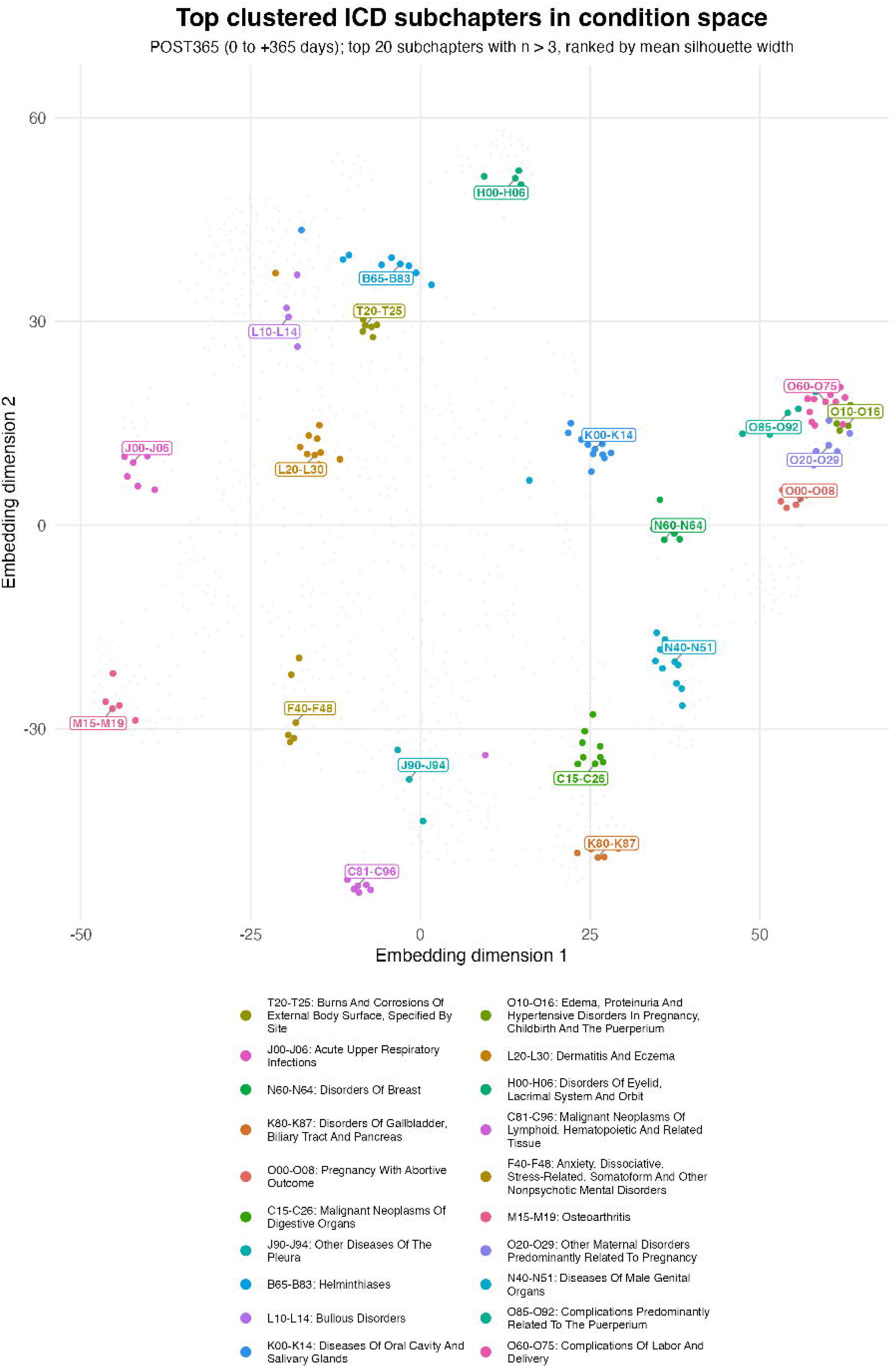

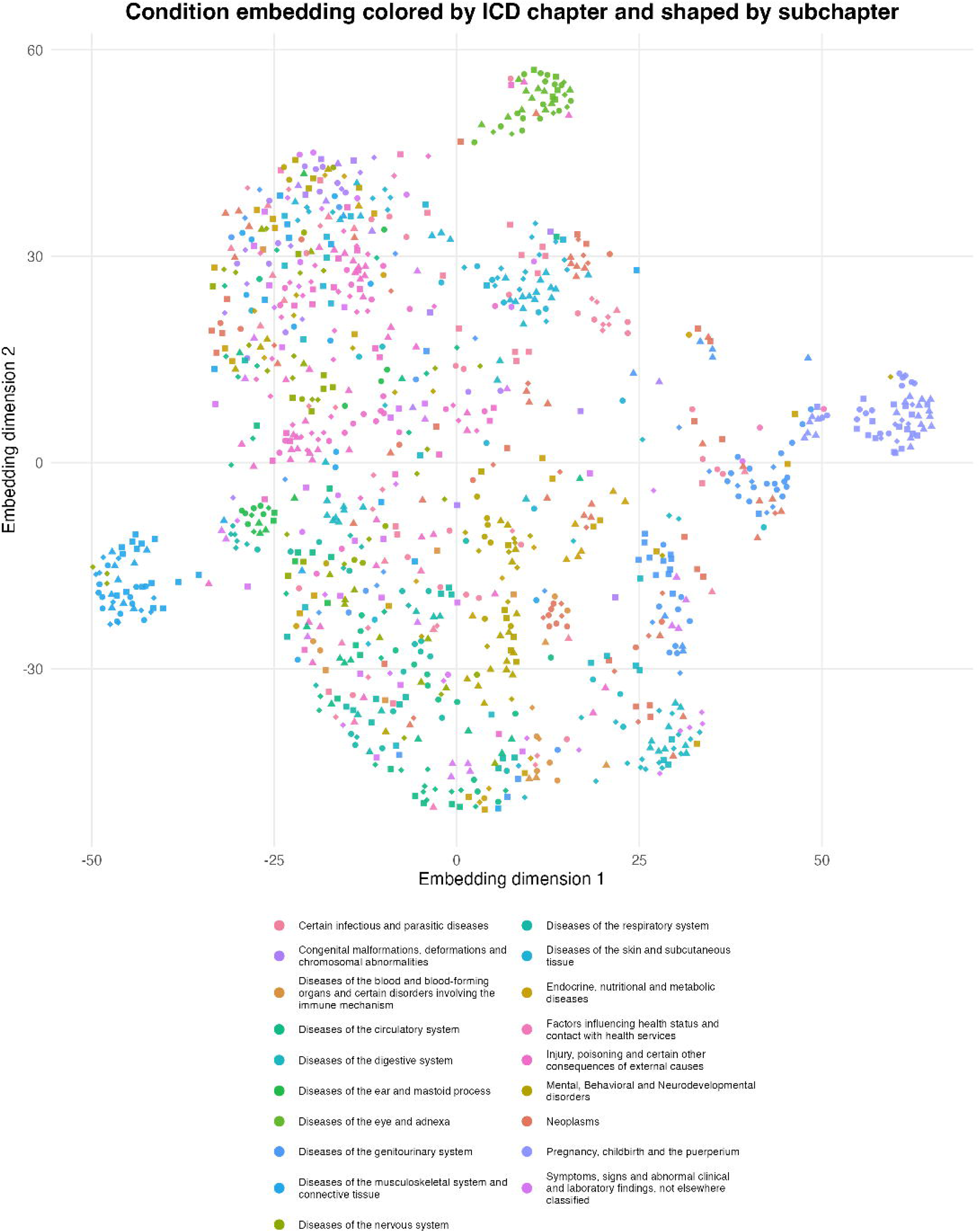

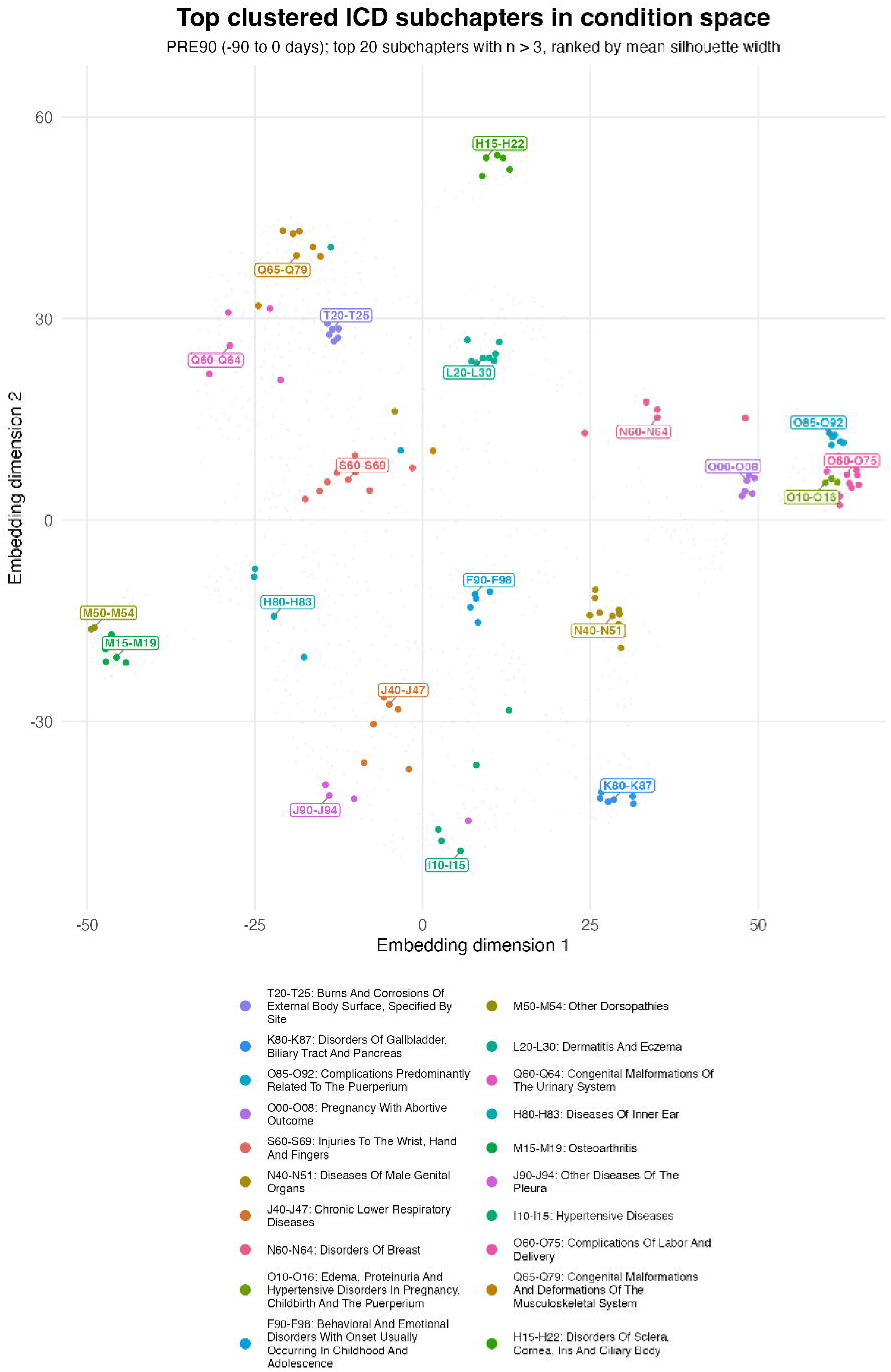

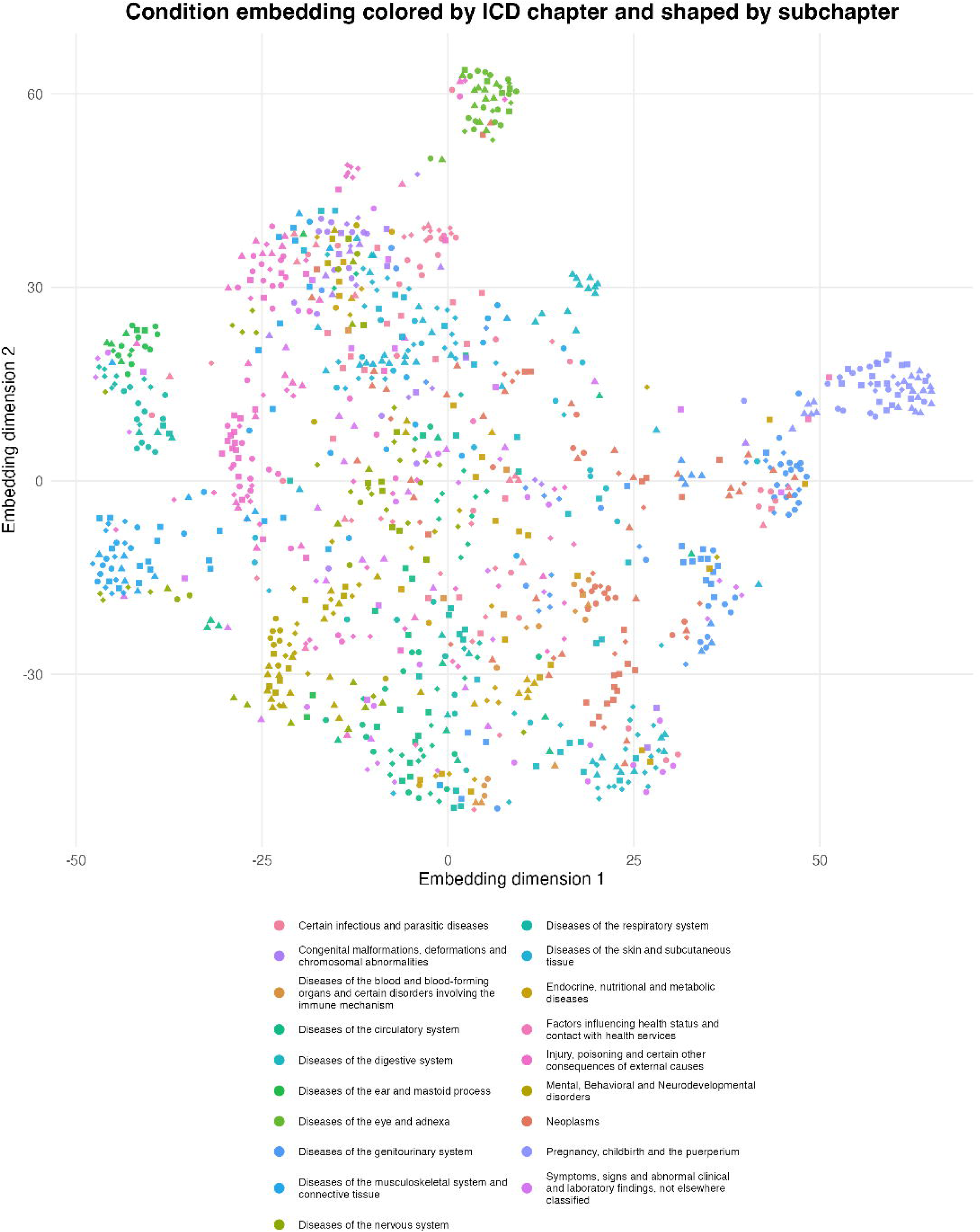

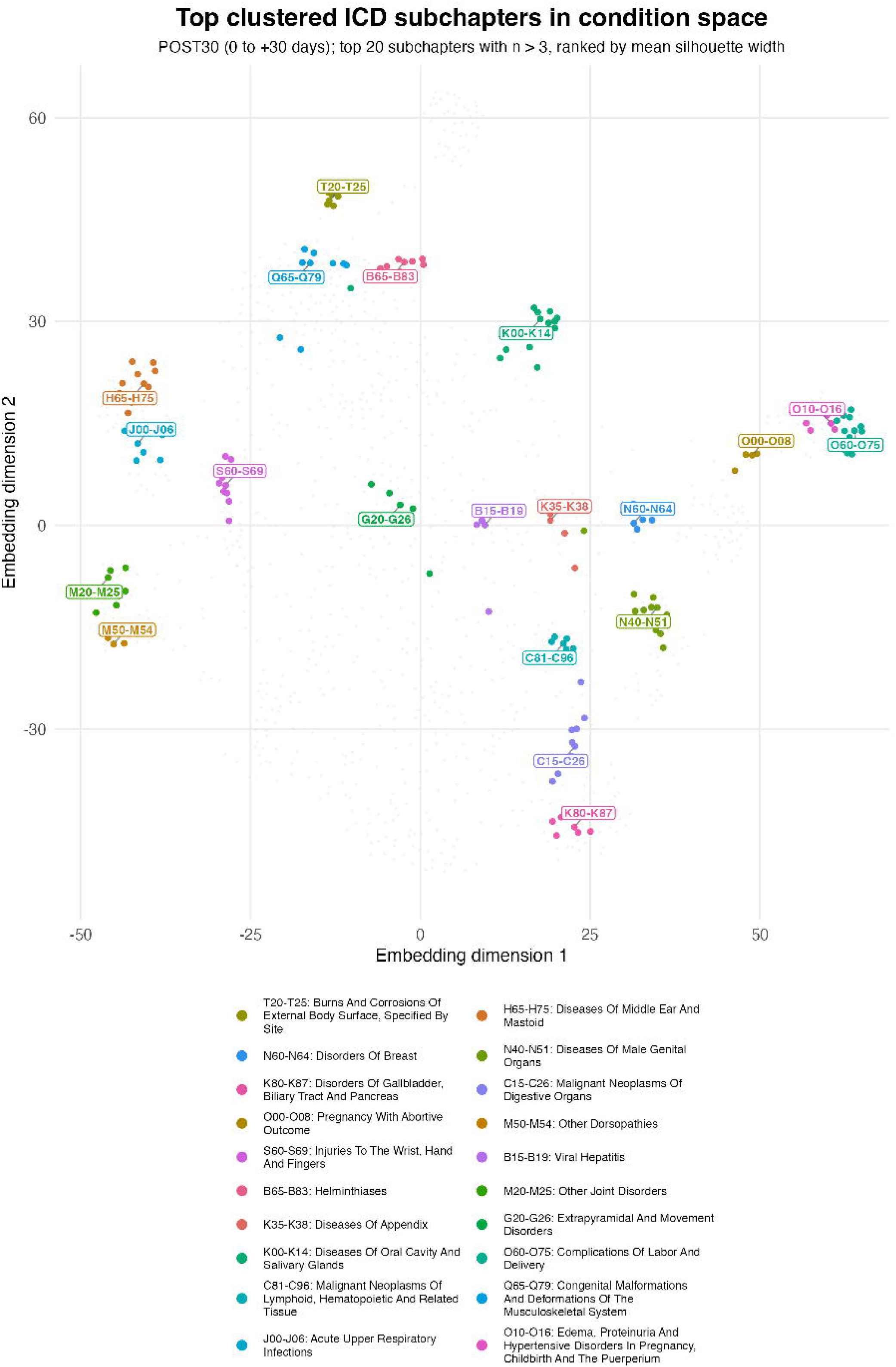

