## Appendix 2 for "A national-scale digital atlas of recorded care patterns across more than 1000 diagnostic categories using real-world health data: a retrospective observational study"

**Appendix 2: Expert evaluation of the CohortContrast Atlas**

### Survey instrument

All rating items used a 7-point Likert-type scale; higher values indicate greater agreement. The diagnosis-level block was repeated three times for each respondent.

#### Diagnosis-level evaluation

1. Diagnosis reviewed (ICD-10 code).
2. Was this diagnosis within your clinical specialty or area of expertise?
3. How much time did you spend reviewing this view?
4. The concepts and temporal patterns shown in this view were clinically plausible for this diagnosis.
5. This view was useful as a starting point for disease-specific research planning.
6. The atlas view was easy to interpret without needing additional explanation.
7. Could any concept, pattern, or visual output in this view be misleading?
8. What useful clinical insight, if any, did this view provide?
9. Please list any concrete research question, hypothesis, or cohort-design application suggested by this view.
10. If you had any additional thoughts while reviewing the disease, express them here.

#### Overall evaluation

11. Overall, the atlas is useful for clinical orientation to recorded care patterns around a diagnosis.
12. Overall, the atlas is useful for generating disease-specific research questions.
13. Overall, the atlas is useful for refining observational study design, such as cohort definitions, covariate selection, outcome definitions, or follow-up windows.
14. The atlas provides information that would be difficult to obtain from conventional tables, registry extracts, or code lists alone.
15. What was the main way in which the atlas could support research planning?
16. What was the main limitation or risk of misinterpretation when using the atlas?
17. Any additional comments about the atlas overall?

### Descriptive summary of ratings

| **Rating item** | **n** | **Mean** | **Median** | **SD** |
| --- | --- | --- | --- | --- |
| Clinical plausibility | 30 | 5.33 | 5 | 1.60 |
| Useful starting point for disease-specific research | 30 | 5.13 | 5.5 | 1.61 |
| Easy to interpret without additional explanation | 30 | 4.50 | 5 | 1.28 |
| Overall usefulness for clinical orientation | 10 | 5.80 | 5.5 | 0.92 |
| Overall usefulness for generating research questions | 10 | 6.30 | 6.5 | 0.82 |
| Overall usefulness for refining observational study design | 10 | 5.40 | 5.5 | 0.97 |
| Provides information difficult to obtain conventionally | 10 | 6.00 | 6 | 0.82 |

### Diagnosis-level ratings pool 30 reviews contributed by ten respondents; overall ratings use ten respondents. SD denotes the sample standard deviation. Statistics are descriptive because each respondent contributed three diagnosis-level reviews.

### Anonymized responses

Respondent identifiers are used only to group the three diagnosis-level reviews with the corresponding overall evaluation.

### Respondent R01

*Three diagnosis-level reviews followed by an overall evaluation.*

#### Review 1: I48

| **Diagnosis** | **Within expertise** | **Time spent** | **Plausibility** | **Research usefulness** | **Interpretability** |
| --- | --- | --- | --- | --- | --- |
| I48 | Yes | >30 minutes | 5/7 | 5/7 | 4/7 |

**Could any concept, pattern, or visual output in this view be misleading?**

What is "emergency room and inpatient visit"? Is this only ER visits? Or is it also ambulatory care?

**What useful clinical insight, if any, did this view provide?**

It showed me the main pattern of anticoagulant treatments we are currently using.

**Please list any concrete research question, hypothesis, or cohort-design application suggested by this view.**

How does atrial fibrillation provoked by systemic infection (I saw it from the procalcitonin measurement difference between groups) differ from what is not associated with systemic infection.

**Additional thoughts while reviewing this diagnosis.**

None to express

#### Review 2: I21

| **Diagnosis** | **Within expertise** | **Time spent** | **Plausibility** | **Research usefulness** | **Interpretability** |
| --- | --- | --- | --- | --- | --- |
| i21 | Yes | 15-30 minutes | 5/7 | 6/7 | 3/7 |

**Could any concept, pattern, or visual output in this view be misleading?**

Is difficult to understand what does cluster reference line mean when u look at for example age or male%.

**What useful clinical insight, if any, did this view provide?**

Did not receive specific clinical insight

**Please list any concrete research question, hypothesis, or cohort-design application suggested by this view.**

None to derive from here.

**Additional thoughts while reviewing this diagnosis.**

Would be good if this would also have information which we base our decisions on (cardiac function etc).

#### Review 3: Z95

| **Diagnosis** | **Within expertise** | **Time spent** | **Plausibility** | **Research usefulness** | **Interpretability** |
| --- | --- | --- | --- | --- | --- |
| z95 | Yes | <15 minutes | 1/7 | 1/7 | 5/7 |

**Could any concept, pattern, or visual output in this view be misleading?**

This is not a good example. The diagnostic code itself is wider than I would like to choose. It clrearly clustered HIV positive and negative patients for me, which is not a valid line of reserach in these patients, as they are cardiac device patients (angiography with stenting or new heart valve or for example a pacemaker).

**What useful clinical insight, if any, did this view provide?**

None

**Please list any concrete research question, hypothesis, or cohort-design application suggested by this view.**

How does HIV+ status affect outcome in patients with cardiac devices.

**Additional thoughts while reviewing this diagnosis.**

None to report.

#### Overall evaluation

| **Clinical orientation** | **Question generation** | **Study-design support** | **Information value** |
| --- | --- | --- | --- |
| 5/7 | 6/7 | 6/7 | 5/7 |

**What was the main way in which the atlas could support research planning?**

Guide me towards patterns which I have not yet recognised in my day-to-day clinical life or current research.

**What was the main limitation or risk of misinterpretation when using the atlas?**

Diagnostic codes are very wide, no option to choose more exact codes.

**Any additional comments about the atlas overall?**

No other comments.

### Respondent R02

*Three diagnosis-level reviews followed by an overall evaluation.*

#### Review 1: I46

| **Diagnosis** | **Within expertise** | **Time spent** | **Plausibility** | **Research usefulness** | **Interpretability** |
| --- | --- | --- | --- | --- | --- |
| I46 | Yes | 15-30 minutes | 4/7 | 4/7 | 5/7 |

**Could any concept, pattern, or visual output in this view be misleading?**

Prevalence bar chart does not demonstrate in/scale bars; Fine needle aspiration biopsy is probably a misnomer

**What useful clinical insight, if any, did this view provide?**

Medications, but unsure of context (is it prescriptions, written recs or pharmacy fillings?)

**Please list any concrete research question, hypothesis, or cohort-design application suggested by this view.**

The idea is worthwhile, should be developed further.

**Additional thoughts while reviewing this diagnosis.**

Make the domain Legend in Dashboard into selection buttons

#### Review 2: I21

| **Diagnosis** | **Within expertise** | **Time spent** | **Plausibility** | **Research usefulness** | **Interpretability** |
| --- | --- | --- | --- | --- | --- |
| I21 | Yes | 15-30 minutes | 5/7 | 4/7 | 5/7 |

**Could any concept, pattern, or visual output in this view be misleading?**

Cluster names

**What useful clinical insight, if any, did this view provide?**

Cohort size

**Please list any concrete research question, hypothesis, or cohort-design application suggested by this view.**

Should enable cohort splitting by presence of concomitant diseases

**Additional thoughts while reviewing this diagnosis.**

*No response*

#### Review 3: Z95

| **Diagnosis** | **Within expertise** | **Time spent** | **Plausibility** | **Research usefulness** | **Interpretability** |
| --- | --- | --- | --- | --- | --- |
| Z95 | Yes | 15-30 minutes | 5/7 | 5/7 | 5/7 |

**Could any concept, pattern, or visual output in this view be misleading?**

Some variables were clustered together into one?

**What useful clinical insight, if any, did this view provide?**

Demographics

**Please list any concrete research question, hypothesis, or cohort-design application suggested by this view.**

Subgroup definition

**Additional thoughts while reviewing this diagnosis.**

*No response*

#### Overall evaluation

| **Clinical orientation** | **Question generation** | **Study-design support** | **Information value** |
| --- | --- | --- | --- |
| 5/7 | 6/7 | 4/7 | 5/7 |

**What was the main way in which the atlas could support research planning?**

Cohort planning decision tree

**What was the main limitation or risk of misinterpretation when using the atlas?**

Some of the plots and axis labels were difficult to understan.

**Any additional comments about the atlas overall?**

Add a knowledge graph feature?

### Respondent R03

*Three diagnosis-level reviews followed by an overall evaluation.*

#### Review 1: G43

| **Diagnosis** | **Within expertise** | **Time spent** | **Plausibility** | **Research usefulness** | **Interpretability** |
| --- | --- | --- | --- | --- | --- |
| G43 | No | 15-30 minutes | 5/7 | 6/7 | 2/7 |

**Could any concept, pattern, or visual output in this view be misleading?**

*No response*

**What useful clinical insight, if any, did this view provide?**

For G43, I found the information on comorbid conditions on the Overlap page particularly useful.

**Please list any concrete research question, hypothesis, or cohort-design application suggested by this view.**

How common are anxiety disorders among patients with migraine?

**Additional thoughts while reviewing this diagnosis.**

*No response*

#### Review 2: S72

| **Diagnosis** | **Within expertise** | **Time spent** | **Plausibility** | **Research usefulness** | **Interpretability** |
| --- | --- | --- | --- | --- | --- |
| S72 | No | 15-30 minutes | 4/7 | 6/7 | 4/7 |

**Could any concept, pattern, or visual output in this view be misleading?**

*No response*

**What useful clinical insight, if any, did this view provide?**

*No response*

**Please list any concrete research question, hypothesis, or cohort-design application suggested by this view.**

What medications were used during the 90 days before the index event? Which comorbid conditions were present during the 90 days before the index event?

**Additional thoughts while reviewing this diagnosis.**

*No response*

#### Review 3: C61

| **Diagnosis** | **Within expertise** | **Time spent** | **Plausibility** | **Research usefulness** | **Interpretability** |
| --- | --- | --- | --- | --- | --- |
| C61 | No | 15-30 minutes | 4/7 | 6/7 | 3/7 |

**Could any concept, pattern, or visual output in this view be misleading?**

*No response*

**What useful clinical insight, if any, did this view provide?**

*No response*

**Please list any concrete research question, hypothesis, or cohort-design application suggested by this view.**

I wanted to know how many people in the target group who underwent surgery also received rehabilitation services (physiotherapy).

**Additional thoughts while reviewing this diagnosis.**

*No response*

#### Overall evaluation

| **Clinical orientation** | **Question generation** | **Study-design support** | **Information value** |
| --- | --- | --- | --- |
| 6/7 | 7/7 | 4/7 | 6/7 |

**What was the main way in which the atlas could support research planning?**

The atlas helps provide a more comprehensive epidemiological overview.

**What was the main limitation or risk of misinterpretation when using the atlas?**

Even after a couple of hours of practice, it was not yet easy to use, creating a risk that results could be misinterpreted or not found at all.

**Any additional comments about the atlas overall?**

*No response*

### Respondent R04

*Three diagnosis-level reviews followed by an overall evaluation.*

#### Review 1: I48

| **Diagnosis** | **Within expertise** | **Time spent** | **Plausibility** | **Research usefulness** | **Interpretability** |
| --- | --- | --- | --- | --- | --- |
| I48 | Yes | >30 minutes | 7/7 | 7/7 | 6/7 |

**Could any concept, pattern, or visual output in this view be misleading?**

*No response*

**What useful clinical insight, if any, did this view provide?**

There were several interesting observations. For example, the association between an emergency department visit and apixaban suggested which anticoagulant is preferentially prescribed from the emergency department. It was also very interesting to see which investigations and tests were performed, and in what proportion of patients, immediately (within 30 days) after an atrial fibrillation diagnosis.

**Please list any concrete research question, hypothesis, or cohort-design application suggested by this view.**

1. Early ablation or cardioversion and its association with mortality.

2. The effect of antiarrhythmic treatment on mortality.

3. The association between initiation of antiarrhythmic drugs and hospitalisation.

4. Comparison of different anticoagulants in relation to complications such as stroke.

**Additional thoughts while reviewing this diagnosis.**

A very interesting dataset on patients with atrial fibrillation. I particularly liked the median first-occurrence approach, which provides a good overview of which medications are prescribed and how quickly.

#### Review 2: I46

| **Diagnosis** | **Within expertise** | **Time spent** | **Plausibility** | **Research usefulness** | **Interpretability** |
| --- | --- | --- | --- | --- | --- |
| I46 | Yes | >30 minutes | 7/7 | 7/7 | 6/7 |

**Could any concept, pattern, or visual output in this view be misleading?**

It was somewhat misleading to view the 90 days before cardiac arrest and see a death certificate recorded in 37.87% before the cardiac arrest. This is understandably related to the timing of diagnosis-code entry and death-certificate recording, because both occurred within the same period.

**What useful clinical insight, if any, did this view provide?**

The demographic distribution of patients who had experienced cardiac arrest seemed interesting and useful. There was a clear twofold predominance of men, and the proportion of successful resuscitations was 57%.

**Please list any concrete research question, hypothesis, or cohort-design application suggested by this view.**

If the pre-diagnosis period for cardiac arrest were longer than 90 days, it might be possible to model which medications are most important for reducing cardiac-arrest risk and, ideally, examine differences within medication classes, such as between different beta-blockers.

**Additional thoughts while reviewing this diagnosis.**

*No response*

#### Review 3: I44

| **Diagnosis** | **Within expertise** | **Time spent** | **Plausibility** | **Research usefulness** | **Interpretability** |
| --- | --- | --- | --- | --- | --- |
| I44 | Yes | >30 minutes | 7/7 | 7/7 | 6/7 |

**Could any concept, pattern, or visual output in this view be misleading?**

Probably not misleading, but requiring further investigation. There were more than 200 very young patients (younger than 30 years) with an I44 diagnosis. However, this diagnosis code includes both milder conditions and more severe conditions requiring pacemaker implantation. Analysis at the subcode level would help clarify this.

**What useful clinical insight, if any, did this view provide?**

It was easy to see how quickly a pacemaker was implanted after atrioventricular block: a median of 3 days, in 25% of cases.

**Please list any concrete research question, hypothesis, or cohort-design application suggested by this view.**

1. The same analyses should be performed at the ICD subcode level because this code includes situations of very different severity, ranging from those requiring only monitoring to life-threatening conditions. This may also explain the bimodal distribution on the Demographics page.

2. Several research questions could be derived, including the association between pacemaker implantation and subsequent heart failure.

3. With a longer lookback, one could examine which comorbidities and medications correlate most strongly with the development of atrioventricular block. This would require a period longer than 90 days and analysis by ICD subcode.

**Additional thoughts while reviewing this diagnosis.**

*No response*

#### Overall evaluation

| **Clinical orientation** | **Question generation** | **Study-design support** | **Information value** |
| --- | --- | --- | --- |
| 7/7 | 7/7 | 7/7 | 6/7 |

**What was the main way in which the atlas could support research planning?**

The atlas provides a good overview of the basic characteristics of a disease group and enables more specific research hypotheses to be formulated. For some disease groups, good population-level statistics on basic features such as age distribution, sex distribution, and prevalence are currently lacking. Expanding from the EST-Health 30% sample to the full population would make the resource especially valuable for comparing real-world prescribing and investigation patterns with guideline recommendations, such as anticoagulant prescribing after atrial fibrillation diagnosis.

**What was the main limitation or risk of misinterpretation when using the atlas?**

Events that occur very close together may sometimes be difficult to interpret, although they are usually rationally explainable. One example is the death certificate recorded during the 90-day period before cardiac arrest.

**Any additional comments about the atlas overall?**

A very interesting dataset.

### Respondent R05

*Three diagnosis-level reviews followed by an overall evaluation.*

#### Review 1: I50

| **Diagnosis** | **Within expertise** | **Time spent** | **Plausibility** | **Research usefulness** | **Interpretability** |
| --- | --- | --- | --- | --- | --- |
| I50 | Partly | 15-30 minutes | 7/7 | 7/7 | 6/7 |

**Could any concept, pattern, or visual output in this view be misleading?**

"Emergency Room and Inpatient Visit" combines events that are not clinically equivalent, making interpretation more difficult. "Death" and "Death Certificate" are also displayed as separate concepts.

**What useful clinical insight, if any, did this view provide?**

The clearest and most useful finding was the concentration of acute-care concepts in C1 when viewing up to one year after I50 diagnosis. This provides a concrete basis for investigating a possible severe-decompensation or intensive-care subgroup.

**Please list any concrete research question, hypothesis, or cohort-design application suggested by this view.**

1. Does heart failure recorded during the 90 days before the index diagnosis, or use of torasemide, distinguish patients with previously known heart failure from patients who may be newly diagnosed, and do their one-year hospitalisation and mortality differ?

2. Does a pre-diagnosis acute-care pattern predict post-diagnosis membership in the more intensive C1 care-profile cluster, recurrent hospitalisation, or death?

3. Do the post-diagnosis differences between C1 and C2 persist after accounting for age and sex?

**Additional thoughts while reviewing this diagnosis.**

*No response*

#### Review 2: I10

| **Diagnosis** | **Within expertise** | **Time spent** | **Plausibility** | **Research usefulness** | **Interpretability** |
| --- | --- | --- | --- | --- | --- |
| I10 | Partly | 15-30 minutes | 6/7 | 6/7 | 6/7 |

**Could any concept, pattern, or visual output in this view be misleading?**

The pre-diagnosis view was somewhat confusing because 70% already had I10 recorded during the 90 days before the index diagnosis. For study planning, it would be useful to filter out people who had an I10 diagnosis before the index date, but this did not appear to be possible.

**What useful clinical insight, if any, did this view provide?**

Twenty-four-hour blood-pressure monitoring occurred in 15% of patients, at a median of 20 days after the index diagnosis; the median was 14 days in C1 and 36 days in C2. This could be used to investigate whether earlier ambulatory blood-pressure monitoring is associated with earlier treatment initiation or adjustment.

**Please list any concrete research question, hypothesis, or cohort-design application suggested by this view.**

1. Is early 24-hour blood-pressure monitoring after an I10 diagnosis associated with earlier initiation of or adjustment to antihypertensive treatment?

2. Does pre-diagnosis electrocardiographic monitoring identify an older subgroup or one with a greater burden of cardiovascular comorbidity, and do post-diagnosis treatment choice or monitoring intensity differ in these patients?

**Additional thoughts while reviewing this diagnosis.**

*No response*

#### Review 3: I48

| **Diagnosis** | **Within expertise** | **Time spent** | **Plausibility** | **Research usefulness** | **Interpretability** |
| --- | --- | --- | --- | --- | --- |
| I48 | Partly | 15-30 minutes | 7/7 | 7/7 | 6/7 |

**Could any concept, pattern, or visual output in this view be misleading?**

*No response*

**What useful clinical insight, if any, did this view provide?**

A strong acute-care subgroup could be identified before diagnosis. After diagnosis, a cluster with a similarly intensive care profile had more echocardiography, invasive monitoring, and death. The distribution of anticoagulants was also useful.

**Please list any concrete research question, hypothesis, or cohort-design application suggested by this view.**

1. Does early echocardiography alter the care pathway after an I48 diagnosis?

2. Which combinations of pre-diagnosis investigations can be distinguished, and are they associated with different post-diagnosis treatment patterns?

**Additional thoughts while reviewing this diagnosis.**

*No response*

#### Overall evaluation

| **Clinical orientation** | **Question generation** | **Study-design support** | **Information value** |
| --- | --- | --- | --- |
| 7/7 | 7/7 | 6/7 | 7/7 |

**What was the main way in which the atlas could support research planning?**

The atlas helps define subgroups suitable for further analysis, establish inclusion and exclusion criteria, and identify events that researchers might not otherwise think to examine.

**What was the main limitation or risk of misinterpretation when using the atlas?**

The main limitation is that diagnoses cannot be combined. For example, I48 cannot be analysed in the same view together with heart failure, hypertension, stroke, or kidney disease, although these comorbidities strongly influence treatment choice and prognosis.

**Any additional comments about the atlas overall?**

*No response*

### Respondent R06

*Three diagnosis-level reviews followed by an overall evaluation.*

#### Review 1: j18

| **Diagnosis** | **Within expertise** | **Time spent** | **Plausibility** | **Research usefulness** | **Interpretability** |
| --- | --- | --- | --- | --- | --- |
| j18 | Partly | 15-30 minutes | 3/7 | 4/7 | 3/7 |

**Could any concept, pattern, or visual output in this view be misleading?**

Radiology is incompletely represented. The diagnosis of pneumonia begins with radiography, not CT, yet radiographs are not represented at all. Ultrasound examinations of the lungs, pleura, abdomen, and pelvis are also absent, while radiotherapy, which is marginal in this setting, is shown. Important procedures such as pleural drainage and biopsy, including whether they were guided by ultrasound or CT, are not shown.

**What useful clinical insight, if any, did this view provide?**

The information would be useful if specialists from the relevant fields, such as radiology and laboratory medicine, were consulted.

**Please list any concrete research question, hypothesis, or cohort-design application suggested by this view.**

I did not understand the basis for forming the two groups. Radiology would be interesting, but procedures performed in patients - radiography, ultrasound, and other interventions - were not represented. Their use across patient ages, care stages, and clinical settings would be valuable for clinical practice, care-quality assessment, and resource use.

**Additional thoughts while reviewing this diagnosis.**

The analysis would be interesting, but the processing of the data is incomplete.

#### Review 2: I63

| **Diagnosis** | **Within expertise** | **Time spent** | **Plausibility** | **Research usefulness** | **Interpretability** |
| --- | --- | --- | --- | --- | --- |
| I63 | Partly | 15-30 minutes | 4/7 | 4/7 | 4/7 |

**Could any concept, pattern, or visual output in this view be misleading?**

Radiology is incompletely represented. An important diagnostic component, CT perfusion, is missing even though it has a separate code and informs major treatment decisions. CT angiography, a diagnostic procedure, has been combined with conventional angiography, which is a treatment procedure; these should not be combined, or at least should be viewable separately. It is important to know how many patients receive CT angiography and what smaller proportion proceed to conventional angiography. Head-only CT angiography should be distinguishable from combined neck and head angiography. Chest radiography, abdominal ultrasound, Doppler examination of the carotid arteries, CT examinations of other body regions, MR perfusion, and swallowing fluoroscopy are missing. MRI should distinguish native MRI, time-of-flight angiography, and contrast-enhanced MR angiography.

**What useful clinical insight, if any, did this view provide?**

The radiology data cannot be examined adequately and do not reflect clinical practice, even though radiology is central to stroke diagnosis and treatment.

**Please list any concrete research question, hypothesis, or cohort-design application suggested by this view.**

Stroke diagnostics and treatment procedures are important hospital-quality indicators. One research question is whether stroke diagnostics are performed similarly across different types of hospitals. Another is the diagnosis and treatment of childhood stroke, for which national data are lacking.

**Additional thoughts while reviewing this diagnosis.**

Good work, but clinical specialists should review which data are included.

#### Review 3: K35

| **Diagnosis** | **Within expertise** | **Time spent** | **Plausibility** | **Research usefulness** | **Interpretability** |
| --- | --- | --- | --- | --- | --- |
| K35 | Partly | 15-30 minutes | 5/7 | 4/7 | 4/7 |

**Could any concept, pattern, or visual output in this view be misleading?**

Radiology is again missing, including chest radiography, abdominal radiography, contrast abdominal radiography, and fluoroscopy. It was unclear how the concept combining CT examination and two body regions was constructed, because it may include other regions such as the lungs.

**What useful clinical insight, if any, did this view provide?**

The radiology component is interesting but somewhat incomplete.

**Please list any concrete research question, hypothesis, or cohort-design application suggested by this view.**

Diagnostic and treatment choices for children across different hospital types could be compared, as these may differ between county and regional hospitals. Investigations and treatment strategies in pregnant patients, which differ from usual practice, could also be studied.

**Additional thoughts while reviewing this diagnosis.**

Good work, but radiology is again incompletely represented and does not cover the full diagnostic process.

#### Overall evaluation

| **Clinical orientation** | **Question generation** | **Study-design support** | **Information value** |
| --- | --- | --- | --- |
| 5/7 | 5/7 | 5/7 | 5/7 |

**What was the main way in which the atlas could support research planning?**

The atlas is well suited to obtaining an initial overview of the situation.

**What was the main limitation or risk of misinterpretation when using the atlas?**

Some data are missing or have been combined too broadly. Radiology, laboratory medicine, and pathology are foundations of modern diagnostics. Radiology coding is designed for reimbursement rather than substantive data analysis, so specialists from the relevant fields need to be involved.

**Any additional comments about the atlas overall?**

A visually appealing resource, but its design requires further development.

### Respondent R07

*Three diagnosis-level reviews followed by an overall evaluation.*

#### Review 1: I48

| **Diagnosis** | **Within expertise** | **Time spent** | **Plausibility** | **Research usefulness** | **Interpretability** |
| --- | --- | --- | --- | --- | --- |
| I48 | Partly | <15 minutes | 4/7 | 5/7 | 5/7 |

**Could any concept, pattern, or visual output in this view be misleading?**

Some visit types, such as follow-up visits and nurse appointments, are classified under observations.

**What useful clinical insight, if any, did this view provide?**

The clusters are promising. For this diagnosis, a likely COVID-related subgroup was distinguished, with higher mortality and more respiratory problems.

**Please list any concrete research question, hypothesis, or cohort-design application suggested by this view.**

For I48, and many other chronic conditions, a post-diagnosis period longer than one year would be appropriate. These data could be used to describe differences in disease management during the COVID period and, if longer follow-up were available, their effect on subsequent disease course and management.

**Additional thoughts while reviewing this diagnosis.**

It would be useful to view medications by group, such as anticoagulants or rate-control medications. Health care changed substantially during 2015-2025, so calendar-year filters would help distinguish recent care, for example 2023-2025. In some analyses, it would also be useful to exclude or separately examine the COVID period.

#### Review 2: M16

| **Diagnosis** | **Within expertise** | **Time spent** | **Plausibility** | **Research usefulness** | **Interpretability** |
| --- | --- | --- | --- | --- | --- |
| M16 | No | <15 minutes | 6/7 | 6/7 | 6/7 |

**Could any concept, pattern, or visual output in this view be misleading?**

For chronic diseases and conditions affecting older people, the enrichment threshold must be lowered to display expected procedures and medications. Because these patients are often multimorbid, however, this also introduces considerable noise. The view may therefore show either too little or too much.

**What useful clinical insight, if any, did this view provide?**

The post-diagnosis care overview would be more useful for this diagnosis with approximately three years of follow-up, because joint replacement does not necessarily occur within one year. It was surprising how few diagnosed patients received lifestyle interventions and physiotherapy, which are standard approaches elsewhere.

**Please list any concrete research question, hypothesis, or cohort-design application suggested by this view.**

It would be useful to examine several codes together, because an endoprosthesis pathway includes both M16 and M17. With a longer period covering joint replacement, changes in service use could be compared between patients who did and did not receive a prosthesis. Service-use data could also support cost-effectiveness analysis.

**Additional thoughts while reviewing this diagnosis.**

*No response*

#### Review 3: E11

| **Diagnosis** | **Within expertise** | **Time spent** | **Plausibility** | **Research usefulness** | **Interpretability** |
| --- | --- | --- | --- | --- | --- |
| E11 | No | 15-30 minutes | 6/7 | 5/7 | 5/7 |

**Could any concept, pattern, or visual output in this view be misleading?**

Visits are distributed across three categories but do not include information on non-pharmacological interventions such as assessment, lifestyle counselling, and foot or eye examinations. In the three-month pre-diagnosis view, is the day axis reversed? Ninety days should appear to the left of the event.

**What useful clinical insight, if any, did this view provide?**

This information would be more useful with a longer period for this chronic condition, allowing the dynamics of complications to be examined if the content of visits and observations were also interpretable.

**Please list any concrete research question, hypothesis, or cohort-design application suggested by this view.**

With longer post-diagnosis follow-up, one could model associations between complications, such as diabetic foot ulcer and subsequent amputation, and patterns of patient monitoring and medication use.

**Additional thoughts while reviewing this diagnosis.**

For acute conditions such as stroke, a longer pre-diagnosis period would be useful. For chronic conditions such as diabetes and arrhythmias, a longer post-diagnosis period of at least three years would be more useful. If users could also compare medication users with non-users or compare other interventions, the resource could support a very large number of studies.

#### Overall evaluation

| **Clinical orientation** | **Question generation** | **Study-design support** | **Information value** |
| --- | --- | --- | --- |
| 6/7 | 6/7 | 6/7 | 6/7 |

**What was the main way in which the atlas could support research planning?**

For example, the atlas could support planning a study of which interventions, including medications, in the I48 atrial-fibrillation treatment regimen most effectively prevent stroke.

**What was the main limitation or risk of misinterpretation when using the atlas?**

The main limitation is source-data quality. Current information systems do not allow clinicians to describe visits or interventions in sufficient detail, leaving substantial information missing. Visit-related data are also divided among observations, visits, and visit details, creating unnecessary noise. Lowering enrichment thresholds to expose relevant information in multimorbid groups may add concepts related to other conditions or generally poorer health, creating a further risk of misinterpretation.

**Any additional comments about the atlas overall?**

*No response*

### Respondent R08

*Three diagnosis-level reviews followed by an overall evaluation.*

#### Review 1: I21

| **Diagnosis** | **Within expertise** | **Time spent** | **Plausibility** | **Research usefulness** | **Interpretability** |
| --- | --- | --- | --- | --- | --- |
| I21 | Partly | 15-30 minutes | 7/7 | 6/7 | 5/7 |

**Could any concept, pattern, or visual output in this view be misleading?**

Not directly, although the low frequency of some prescriptions was surprising: atorvastatin 40 mg in only 39.32%, clopidogrel in 30.04%, and ticagrelor in 38.1%. This raises the question of whether the rates are truly this low or whether the program is missing something. Filtering for "physical therapy" also produced "physical therapy procedure" and ordinal variants from "physical therapy procedure 1st" through "10th". The median first occurrence differed between "physical therapy procedure" (63 days) and "physical therapy procedure 1st" (5 days), although both appear to refer to a first procedure. Drugs, conditions, observations, and other concepts on the Dashboard should be sortable by prevalence.

**What useful clinical insight, if any, did this view provide?**

The medication information is useful because it shows which medications are prescribed most often and when. The unexpectedly low prescribing frequencies immediately motivate further investigation.

**Please list any concrete research question, hypothesis, or cohort-design application suggested by this view.**

Which patient groups receive physiotherapy earlier during hospital care? Which patients do not receive statins after myocardial infarction? How does management differ between patients experiencing their first infarction and those with recurrent infarction?

**Additional thoughts while reviewing this diagnosis.**

I could not obtain the desired I21 view after selecting only "drug exposure" and then "ordinal rows". It would also be useful if manually selected medications appeared on the graph. Selecting a drug in the concept list currently places a check mark beside it but does not appear to change the visualisation.

#### Review 2: I10

| **Diagnosis** | **Within expertise** | **Time spent** | **Plausibility** | **Research usefulness** | **Interpretability** |
| --- | --- | --- | --- | --- | --- |
| I10 | Yes | <15 minutes | 3/7 | 2/7 | 3/7 |

**Could any concept, pattern, or visual output in this view be misleading?**

The trajectory view suggests that the first antihypertensive prescription occurs about 100 days after diagnosis, which seems implausible. If correct, it would be an important care-quality topic. The principal procedures appeared to be ultrasound and contrast-enhanced CT, seemingly more common among patients with hypertension than in the general population, although these procedures are not required to diagnose hypertension. This raises the possibility that hypertension is identified more often among patients with greater contact with the health-care system.

**What useful clinical insight, if any, did this view provide?**

It is useful to see which investigations and tests occurred before diagnosis and which medications were first prescribed, and when. It would also be useful to see comorbidities, but these did not appear even when the target-prevalence and prevalence-difference-ratio filters were set from 0% to 100%.

**Please list any concrete research question, hypothesis, or cohort-design application suggested by this view.**

What proportion of patients receive blood tests after a hypertension diagnosis, and which tests are performed?

**Additional thoughts while reviewing this diagnosis.**

*No response*

#### Review 3: H65

| **Diagnosis** | **Within expertise** | **Time spent** | **Plausibility** | **Research usefulness** | **Interpretability** |
| --- | --- | --- | --- | --- | --- |
| H65 | Yes | 15-30 minutes | 4/7 | 4/7 | 3/7 |

**Could any concept, pattern, or visual output in this view be misleading?**

For otitis media, approximately 10% of patients received ciprofloxacin and hydrocortisone drops during the preceding 90 days, although these are more typical treatments for otitis externa. The median day value of 90 was unclear: did this mean 90 days before the diagnosis or the day on which H65 was recorded? When filtering to measurements, blood tests were absent and only hearing tests appeared, although C-reactive protein and complete blood count measurements might be expected. The initial view also suggested an older population than expected (median age 26), whereas the detailed demographics view showed that the condition was most frequent among children aged 0-10 years.

**What useful clinical insight, if any, did this view provide?**

Overall, the information is useful and provides an overview of which antibiotics are preferred.

**Please list any concrete research question, hypothesis, or cohort-design application suggested by this view.**

How does patient age affect antibiotic choice? Does antibiotic selection follow current clinical guidelines? Which comorbidities occur among adults with otitis media?

**Additional thoughts while reviewing this diagnosis.**

As noted above, it should be possible to select concepts on the Dashboard and display them on the graph alongside the automatically selected concepts.

#### Overall evaluation

| **Clinical orientation** | **Question generation** | **Study-design support** | **Information value** |
| --- | --- | --- | --- |
| 5/7 | 7/7 | 5/7 | 7/7 |

**What was the main way in which the atlas could support research planning?**

The atlas provides an initial overview of the most common procedures, diagnoses, medications, and other events associated with the diagnosis under study. This is particularly useful when the researcher has limited clinical familiarity with the disease. It could also help identify which aspects of guideline adherence warrant detailed analysis before a full study is undertaken.

**What was the main limitation or risk of misinterpretation when using the atlas?**

The formation of clusters was unclear, making them difficult to use. Many terms may also be unfamiliar to ordinary clinicians, so more tooltips and explanations are needed. Because the atlas is powerful and has many settings, users must spend time learning it; otherwise, obtaining an overview may take a long time or the results may be misinterpreted. This is particularly important for occasional users, who may need to relearn the interface each time they use it.

**Any additional comments about the atlas overall?**

*No response*

### Respondent R09

*Three diagnosis-level reviews followed by an overall evaluation.*

#### Review 1: C53, L40, A53/A51

| **Diagnosis** | **Within expertise** | **Time spent** | **Plausibility** | **Research usefulness** | **Interpretability** |
| --- | --- | --- | --- | --- | --- |
| C53 | Yes | >30 minutes | 7/7 | 7/7 | 5/7 |

**Could any concept, pattern, or visual output in this view be misleading?**

Several aspects were unclear. The timeline should show more clearly where day counting begins; in the 90-day pre-diagnosis view, it was not immediately clear whether 71 meant 71 days before diagnosis or day 71 of the observation window. "Trajectories/Progression" suggests longitudinal progression in the same patients, although the view appears to show aggregate statistics. For automatic clustering, the basis for selecting the number of clusters and the features used to assign patients should be visible. A useful test case was C53 with three clusters: in C1, the median for "Evaluation of biopsy specimen + H&E" was approximately 70, while "Cervical biopsy" was approximately 83. If interpreted as event times, this suggests that specimen evaluation occurred before biopsy. This may not be an error because the medians may concern different patients, mapping, and aggregate values, but it illustrates why the Trajectories view must explain what can and cannot be inferred from these medians.

**What useful clinical insight, if any, did this view provide?**

A brief exploration identified three interesting C53 clusters. C1 was a younger diagnostic group (n=74; mean age 49.6) with extensive cervical-specific diagnostics, including biopsy, colposcopy, PCR, and dysplasia-related concepts, suggesting a clear pre-diagnosis pathway. C2 showed little pre-diagnosis activity (n=270; age 62.8), with relatively few investigations and procedures. C3 was older and intensively investigated (n=127; age 65.5), with substantial CT and laboratory testing, potentially indicating more severe or advanced disease. Men comprised 1.06% of patients with C53, raising a possible data-quality question.

**Please list any concrete research question, hypothesis, or cohort-design application suggested by this view.**

*No response*

**Additional thoughts while reviewing this diagnosis.**

*No response*

#### Review 2: L40

| **Diagnosis** | **Within expertise** | **Time spent** | **Plausibility** | **Research usefulness** | **Interpretability** |
| --- | --- | --- | --- | --- | --- |
| L40 | Yes | 15-30 minutes | 7/7 | 6/7 | 4/7 |

**Could any concept, pattern, or visual output in this view be misleading?**

Can semantically related concepts, such as "Psoriasis" and "Psoriasis vulgaris", be combined and treated as one concept? They currently remain separate, which may fragment results when studying psoriasis more broadly. In Mappings, some more specific forms of psoriasis appear to be mapped to the general concept "Disorder of integument". It was unclear how users can inspect or control this aggregation.

**What useful clinical insight, if any, did this view provide?**

*No response*

**Please list any concrete research question, hypothesis, or cohort-design application suggested by this view.**

*No response*

**Additional thoughts while reviewing this diagnosis.**

*No response*

#### Review 3: A51

| **Diagnosis** | **Within expertise** | **Time spent** | **Plausibility** | **Research usefulness** | **Interpretability** |
| --- | --- | --- | --- | --- | --- |
| A51 | Yes | 15-30 minutes | 7/7 | 7/7 | 5/7 |

**Could any concept, pattern, or visual output in this view be misleading?**

*No response*

**What useful clinical insight, if any, did this view provide?**

Approximately 70% of patients were men, which may indicate substantial transmission among men who have sex with men. It would be interesting to examine how the male-to-female ratio has changed over time, but the tool did not appear to support this.

**Please list any concrete research question, hypothesis, or cohort-design application suggested by this view.**

*No response*

**Additional thoughts while reviewing this diagnosis.**

*No response*

#### Overall evaluation

| **Clinical orientation** | **Question generation** | **Study-design support** | **Information value** |
| --- | --- | --- | --- |
| 7/7 | 7/7 | 6/7 | 7/7 |

**What was the main way in which the atlas could support research planning?**

*No response*

**What was the main limitation or risk of misinterpretation when using the atlas?**

The main limitation is that aggregate data can easily be interpreted as individual patient trajectories or associations that the data may not actually demonstrate. Clinical expertise is therefore required to interpret the results.

**Any additional comments about the atlas overall?**

*No response*

### Respondent R10

### Three diagnosis-level reviews followed by an overall evaluation.

### Review 1: D50

| Diagnosis | Within expertise | Time spent | Plausibility | Research usefulness | Interpretability |
| --- | --- | --- | --- | --- | --- |
| D50 | Partly | 15-30 minutes | 7/7 | 3/7 | 6/7 |

### Could any concept, pattern, or visual output in this view be misleading?

### No response

### What useful clinical insight, if any, did this view provide?

### I was able to identify quite easily the demographic characteristics of patients who received iron injections for treatment of iron deficiency anaemia. I also saw that men were overrepresented, compared with the average iron deficiency anaemia patient, among patients with likely more severe anaemia. It was fairly easy to infer that the two clusters were distinguished mainly by age.

### Please list any concrete research question, hypothesis, or cohort-design application suggested by this view.

### If we wanted to compare oral iron replacement therapy versus intravenous therapy using registry-based data, sex and age would need to be treated as confounders.

### Additional thoughts while reviewing this diagnosis.

### For iron deficiency anaemia, I had to adjust the filters to find interesting information. Compared with, for example, myocardial infarction.

### Review 2: E03

| Diagnosis | Within expertise | Time spent | Plausibility | Research usefulness | Interpretability |
| --- | --- | --- | --- | --- | --- |
| E03 | Partly | 15-30 minutes | 4/7 | 3/7 | 2/7 |

### Could any concept, pattern, or visual output in this view be misleading?

### No response

### What useful clinical insight, if any, did this view provide?

### I would not say it could not be useful, but in this example it was difficult for me to formulate any hypothesis. The automatically generated clusters differed more in background characteristics than in thyroid-related tests or procedures.

### Please list any concrete research question, hypothesis, or cohort-design application suggested by this view.

### No response

### Additional thoughts while reviewing this diagnosis.

### I expected to see what distinguishes people who had TPO or TRAb antibodies tested from those who did not, but I did not see that information because other blood tests dominated. So if one comes with a more specific question, it may be difficult to find this information. It is probably more useful precisely when hypotheses or research questions are lacking.

### Review 3: F33

| Diagnosis | Within expertise | Time spent | Plausibility | Research usefulness | Interpretability |
| --- | --- | --- | --- | --- | --- |
| F33 | No | 15-30 minutes | 7/7 | 4/7 | 3/7 |

### Could any concept, pattern, or visual output in this view be misleading?

### No response

### What useful clinical insight, if any, did this view provide?

### No response

### Please list any concrete research question, hypothesis, or cohort-design application suggested by this view.

### No response

### Additional thoughts while reviewing this diagnosis.

### I was interested in deaths related to major depressive disorder and the disease trajectories preceding them. This platform is not well suited to answering such a question. Otherwise, there is a great deal of information on procedures and visits related to major depressive disorder, and some specialist could certainly make more useful use of it.

### Overall evaluation

| Clinical orientation | Question generation | Study-design support | Information value |
| --- | --- | --- | --- |
| 5/7 | 5/7 | 5/7 | 6/7 |

### What was the main way in which the atlas could support research planning?

### The best use seems to be generating research hypotheses within one’s own field. For example, one could select the top 1 to 3 diagnoses one works with and, without a prior hypothesis or restriction, explore which procedures, measurements, or other data tend to accompany the patient before and after diagnosis.

### What was the main limitation or risk of misinterpretation when using the atlas?

### For me, there were several main limitations: 1. Two diagnoses cannot be compared directly. 2. The data are strictly diagnosis-centred, whereas the questions that often interest us are centred on procedures, medications, or outcomes such as death. 3. At times one would want to narrow the population, for example to patients with headache who attended the emergency department, then ask who underwent CT and who did not, and what distinguished them.

### Any additional comments about the atlas overall?

### I also tried to get feedback from my spouse, but her feedback and that of her endocrinologist colleagues was that this is too complex a tool for an ordinary physician. The demo was not very helpful for them because it seemed too long and the terminology was unclear. To be fair, they tried it in the middle of a workday, so a physician with a major and important research question would probably still work out how to use it.
