## Appendix 3 for "A national-scale digital atlas of recorded care patterns across more than 1000 diagnostic categories using real-world health data: a retrospective observational study"

**Appendix 3: Follow-up window centric metrics**

**Table S1.** Observation window specific summarised workflow results

| **Window** | **-90 to 0 days** | **0 to +30 days** | **0 to +365 days** |
| --- | --- | --- | --- |
| **Mean patients** | 7204* | 7462 | 7462 |
| **Median patients** | 1477 (IQR 396–6863) | 1518 (IQR 411–7093) | 1518 (IQR 411–7093) |
| **Median age** | 49 | 49 | 49 |
| **Median male percentage** | 43.4% | 43.3% | 43.3% |
| **Total initial concepts (average)** | 2770 | 2188 | 4120 |
| **Total initial concepts (median)** | 2379 | 1919 | 3418 |
| **Total passed concepts (average)** | 76.4 | 94.7 | 103.3 |
| **Total passed concepts (median)** | 59 | 71.5 | 75 |
| **Total reduction (weighted)** | 97.8% | 97.0% | 97.8% |
| **Phase II reduction rate (weighted)** | 90.5% | 89.8% | 88.5% |
| **Phase III reduction rate (weighted)** | 68.3% | 59.1% | 73.2% |
| **Phase IV reduction rate (weighted)** | 26.3% | 28.5% | 30% |
| **Best clustering k=2 percentage** | 76.8% | 67.5% | 80.6% |
| **Average silhouette score k=2** | 0.213 | 0.250 | 0.221 |
| **Best clustering k=3 percentage** | 9.6% | 12.8% | 7.1% |
| **Average silhouette score k=3** | 0.126 | 0.160 | 0.132 |
| **Best clustering k=4 percentage** | 4.5% | 7.9% | 5.6% |
| **Average silhouette score k=4** | 0.110 | 0.134 | 0.112 |
| **Best clustering k=5 percentage** | 9.1% | 11.9% | 6.7% |
| **Average silhouette score k=5** | 0.107 | 0.127 | 0.107 |
| **Average best k silhouette score** | 0.224 | 0.234 | 0.229 |
| **Median conditions** | 6 (IQR 2–16) | 9 (IQR 3–25) | 9 (IQR 3–25) |
| **Median drugs** | 4 (IQR 0–15) | 6 (IQR 1–18) | 6 (IQR 1–22) |
| **Median procedures** | 12 (IQR 5–22) | 15 (IQR 7–27) | 17 (IQR 7–33) |
| **Median measurements** | 17 (IQR 9–28) | 20 (IQR 9–33) | 21 (IQR 8–35) |
| **Median observations** | 9 (IQR 4–15) | 10 (IQR 5–18) | 12 (IQR 5–22) |
| **Median visits** | 3 (IQR 2–4) | 3 (IQR 2–4) | 3 (IQR 2–4) |
| **Median visit details** | 3 (IQR 2–4) | 3 (IQR 2–4) | 3 (IQR 2–4) |
| **Median deaths** | NA | 0 (IQR 0–0) | 0 (IQR 0–1) |

*PRE90 counts were lower where the historical comparator window lacked sufficient observability
