## Appendix 4 for "A national-scale digital atlas of recorded care patterns across more than 1000 diagnostic categories using real-world health data: a retrospective observational study"

**Appendix 4. Mapping overview**

**Table S2.** Top 10 mappings over all views for hierarchy-mapping and correlation-mapping types.

| **Top 10 mappings via hierarchy** | | | | |
| --- | --- | --- | --- | --- |
| **Descendant** | **Parent** | **Views (n)** | | **Conditions (n)** |
| CBC panel - Blood by Automated count | CBC W Auto Differential panel - Blood | 1667 | | 677 |
| Fibrinogen screening | Coagulation pathway screening | 1259 | | 514 |
| Intensive Care | Inpatient Visit | 1159 | | 503 |
| CT angiography with contrast | CT with contrast | 1149 | | 471 |
| Prostate specific antigen measurement | Enzyme measurement | 837 | | 367 |
| CT of brain without contrast | CT without contrast | 823 | | 356 |
| Procedure to identify antibody | Immunology laboratory test | 736 | | 346 |
| ABO typing | Immunology laboratory test | 717 | | 391 |
| MRI with contrast | Magnetic resonance imaging | 701 | | 340 |
| Immunochromatographic test | Immunology laboratory test | 622 | | 343 |
| **Top 10 mappings via correlation and co-occurrence** | | | | |
| **Merged concepts** | | **Views (n)** | | **Conditions (n)** |
| Cholesterol measurement + Measurement of total cholesterol and triglycerides | | 1009 | | 580 |
| Insertion of catheter into urinary bladder + Insertion of indwelling catheter into urinary bladder | | 979 | | 431 |
| Analysis using real time PCR + Chromosome analysis, cytogenetic procedure AND/OR molecular biology method | | 867 | | 416 |
| Anesthesia duration + Recovery room monitoring, anesthesia | | 814 | | 394 |
| Evaluation of biopsy specimen + Hematoxylin and eosin stain method | | 735 | | 388 |
| CT with contrast + Radiology of two body areas | | 538 | | 298 |
| Disk diffusion susceptibility test + Microbial identification test | | 501 | | 292 |
| CT with contrast + CT without contrast + Radiology of two body areas | | 417 | | 237 |
| Developing a treatment plan + Multidisciplinary cancer case management | | 412 | | 216 |
| CT without contrast + Radiology of two body areas | | 409 | | 218 |
