## Appendix 5 for "A national-scale digital atlas of recorded care patterns across more than 1000 diagnostic categories using real-world health data: a retrospective observational study"

**Appendix 5: Validation and clustering results per ICD-10 code**

**Table S3.** Validation and clustering results per ICD-10 code

| **icd10** | **target_patients** | **best_k_POST365** | **concepts_PRE90** | **direct_PRE90** | **indirect_PRE90** | **concepts_POST30** | **direct_POST30** | **indirect_POST30** | **concepts_POST365** | **direct_POST365** | **indirect_POST365** |
| --- | --- | --- | --- | --- | --- | --- | --- | --- | --- | --- | --- |
| J06 | 179,982 | 2 | 127 | 44.9 | 28.4 | 213 | 15.0 | 58.7 | 354 | 9.3 | 47.5 |
| B34 | 142,755 | 2 | 121 | 18.0 | 57.4 | 196 | 9.7 | 76.5 | 336 | 5.4 | 59.5 |
| M54 | 130,354 | 2 | 201 | 35.8 | 38.8 | 291 | 31.6 | 39.5 | 436 | 24.3 | 40.4 |
| K04 | 105,961 | 2 | 89 | 51.7 | 16.9 | 61 | 36.1 | 54.1 | 132 | 18.2 | 34.9 |
| H10 | 96,233 | 2 | 235 | 23.8 | 33.6 | 223 | 15.2 | 48.0 | 258 | 14.3 | 43.8 |
| M25 | 94,298 | 2 | 201 | 46.8 | 29.9 | 239 | 32.2 | 38.1 | 345 | 17.1 | 53.9 |
| J20 | 90,804 | 2 | 216 | 37.0 | 32.4 | 293 | 4.4 | 57.7 | 400 | 7.5 | 55.5 |
| H52 | 90,674 | 2 | 261 | 11.1 | 39.5 | 188 | 17.6 | 55.3 | 354 | 6.5 | 47.5 |
| R10 | 88,653 | 2 | 238 | 51.7 | 32.4 | 376 | 34.0 | 51.9 | 458 | 32.3 | 42.8 |
| J02 | 77,224 | 2 | 137 | 40.1 | 32.1 | 167 | 18.0 | 43.1 | 268 | 14.9 | 38.8 |
| E78 | 76,815 | 2 | 269 | 36.4 | 35.7 | 416 | 13.7 | 47.8 | 487 | 9.4 | 49.9 |
| J00 | 76,564 | 2 | 145 | 47.6 | 32.4 | 152 | 5.3 | 74.3 | 246 | 4.5 | 59.8 |
| K29 | 74,129 | 2 | 538 | 21.0 | 28.6 | 506 | 8.5 | 40.9 | 656 | 7.5 | 38.8 |
| J30 | 72,854 | 2 | 265 | 30.6 | 37.4 | 264 | 14.8 | 53.8 | 352 | 12.5 | 46.3 |
| M79 | 72,474 | 2 | 206 | 45.1 | 26.2 | 268 | 28.7 | 48.9 | 356 | 8.7 | 54.5 |
| E55 | 71,846 | 2 | 213 | 26.3 | 39.0 | 479 | 4.8 | 55.1 | 490 | 3.7 | 66.7 |
| N39 | 64,590 | 2 | 468 | 31.4 | 34.8 | 532 | 18.0 | 40.0 | 568 | 20.4 | 38.7 |
| N30 | 64,489 | 4 | 261 | 24.9 | 31.8 | 277 | 14.1 | 48.7 | 384 | 7.8 | 46.1 |
| J01 | 64,431 | 2 | 162 | 36.4 | 33.3 | 185 | 21.6 | 53.0 | 265 | 19.2 | 46.0 |
| I10 | 63,482 | 2 | 311 | 25.1 | 26.4 | 492 | 15.0 | 42.7 | 528 | 12.9 | 43.9 |
| K05 | 62,874 | 2 | 78 | 35.9 | 46.1 | 62 | 45.2 | 48.4 | 129 | 22.5 | 41.9 |
| L30 | 59,102 | 2 | 211 | 32.2 | 29.4 | 209 | 31.1 | 47.9 | 287 | 23.3 | 44.6 |
| K02 | 58,798 | 2 | 32 | 25.0 | 59.4 | 28 | 25.0 | 60.7 | 65 | 9.2 | 40.0 |
| R05 | 57,437 | 2 | 175 | 58.9 | 24.0 | 212 | 44.3 | 34.9 | 276 | 9.8 | 62.7 |
| H61 | 56,744 | 2 | 128 | 13.3 | 52.3 | 155 | 12.3 | 44.5 | 223 | 9.0 | 43.0 |
| H65 | 53,944 | 2 | 161 | 29.2 | 50.9 | 173 | 12.1 | 69.9 | 213 | 8.4 | 63.4 |
| L23 | 53,407 | 2 | 180 | 27.8 | 38.3 | 157 | 13.4 | 52.9 | 248 | 14.5 | 41.5 |
| G47 | 52,795 | 2 | 489 | 28.0 | 37.0 | 444 | 8.6 | 62.4 | 551 | 8.0 | 59.2 |
| I11 | 52,574 | 2 | 419 | 29.6 | 25.3 | 604 | 13.1 | 47.5 | 660 | 12.1 | 50.8 |
| K21 | 52,387 | 2 | 384 | 15.1 | 41.1 | 359 | 8.9 | 41.0 | 460 | 6.7 | 43.3 |
| J03 | 50,714 | 2 | 125 | 35.2 | 40.8 | 161 | 31.1 | 52.2 | 240 | 16.7 | 48.3 |
| J04 | 49,838 | 2 | 118 | 48.3 | 39.8 | 168 | 10.1 | 72.6 | 237 | 6.8 | 63.3 |
| H25 | 49,004 | 2 | 225 | 19.6 | 37.3 | 175 | 16.6 | 44.0 | 242 | 13.6 | 50.8 |
| J35 | 47,328 | 2 | 241 | 32.0 | 39.0 | 261 | 21.1 | 57.9 | 314 | 11.2 | 63.1 |
| M17 | 47,237 | 2 | 170 | 42.4 | 32.4 | 231 | 18.2 | 56.7 | 283 | 21.9 | 49.1 |
| F41 | 45,415 | 2 | 353 | 31.4 | 30.9 | 316 | 23.4 | 46.2 | 393 | 16.8 | 40.5 |
| R07 | 45,338 | 2 | 190 | 56.8 | 32.6 | 268 | 25.4 | 56.7 | 353 | 16.4 | 63.7 |
| D50 | 45,299 | 2 | 329 | 37.4 | 31.3 | 472 | 11.9 | 48.9 | 509 | 11.6 | 51.9 |
| B35 | 44,925 | 2 | 152 | 22.4 | 44.1 | 155 | 16.8 | 56.1 | 225 | 11.6 | 43.1 |
| R51 | 43,590 | 2 | 168 | 47.0 | 38.7 | 236 | 17.8 | 64.0 | 308 | 18.5 | 54.2 |
| K03 | 42,699 | 3 | 41 | 14.6 | 26.8 | 21 | 38.1 | 52.4 | 69 | 4.3 | 47.8 |
| M75 | 42,682 | 2 | 155 | 20.0 | 58.1 | 179 | 15.6 | 60.3 | 223 | 13.0 | 63.7 |
| B37 | 42,453 | 2 | 318 | 47.5 | 30.5 | 310 | 11.6 | 58.4 | 385 | 17.4 | 50.4 |
| E66 | 42,384 | 2 | 274 | 40.1 | 34.7 | 459 | 8.9 | 76.0 | 463 | 6.7 | 71.3 |
| D22 | 40,542 | 2 | 111 | 21.6 | 28.8 | 97 | 9.3 | 41.2 | 157 | 7.6 | 35.0 |
| J18 | 40,531 | 2 | 339 | 33.9 | 36.3 | 489 | 11.9 | 58.7 | 491 | 12.4 | 53.4 |
| M77 | 38,506 | 2 | 109 | 52.3 | 36.7 | 144 | 32.6 | 49.3 | 194 | 24.2 | 50.0 |
| J98 | 38,107 | 2 | 63 | 38.1 | 44.4 | 119 | 37.0 | 52.1 | 196 | 18.9 | 59.2 |
| D23 | 37,631 | 2 | 137 | 21.9 | 44.5 | 125 | 19.2 | 57.6 | 179 | 10.1 | 54.8 |
| S60 | 35,712 | 2 | 67 | 28.4 | 41.8 | 73 | 20.6 | 45.2 | 183 | 19.1 | 41.0 |
| M21 | 35,566 | 2 | 122 | 27.9 | 50.8 | 126 | 11.9 | 69.8 | 174 | 10.9 | 63.8 |
| I50 | 35,024 | 2 | 472 | 33.5 | 35.6 | 511 | 15.3 | 55.8 | 534 | 13.5 | 54.5 |
| I49 | 34,814 | 2 | 263 | 42.2 | 32.7 | 364 | 12.1 | 59.3 | 407 | 11.8 | 51.4 |
| S93 | 34,297 | 2 | 51 | 33.3 | 45.1 | 67 | 17.9 | 64.2 | 106 | 10.4 | 59.4 |
| M51 | 34,192 | 2 | 199 | 36.7 | 37.7 | 215 | 36.5 | 32.5 | 295 | 31.9 | 38.3 |
| K59 | 34,153 | 2 | 226 | 29.2 | 45.1 | 275 | 3.6 | 54.5 | 339 | 6.2 | 53.7 |
| K30 | 33,382 | 2 | 247 | 24.7 | 34.8 | 227 | 10.6 | 50.2 | 286 | 9.4 | 47.9 |
| N76 | 33,237 | 2 | 203 | 32.0 | 42.4 | 181 | 26.5 | 56.4 | 224 | 21.9 | 47.3 |
| J34 | 32,702 | 2 | 235 | 34.9 | 32.3 | 210 | 28.1 | 51.0 | 273 | 20.5 | 49.8 |
| H35 | 32,270 | 2 | 191 | 39.3 | 41.4 | 165 | 12.7 | 58.2 | 233 | 17.6 | 52.8 |
| M70 | 31,714 | 2 | 163 | 40.5 | 38.0 | 213 | 24.4 | 43.7 | 225 | 21.3 | 40.4 |
| M19 | 31,682 | 2 | 154 | 44.8 | 41.6 | 174 | 28.2 | 50.0 | 230 | 25.2 | 45.6 |
| L02 | 31,601 | 2 | 172 | 39.0 | 33.7 | 177 | 19.8 | 58.8 | 210 | 22.9 | 46.2 |
| S61 | 31,587 | 2 | 59 | 55.9 | 35.6 | 83 | 26.5 | 62.6 | 123 | 18.9 | 64.8 |
| S01 | 31,245 | 2 | 103 | 39.8 | 50.5 | 182 | 23.2 | 53.0 | 196 | 21.9 | 61.7 |
| H81 | 30,636 | 2 | 184 | 37.5 | 46.2 | 231 | 15.2 | 52.8 | 267 | 10.5 | 69.3 |
| H60 | 30,276 | 2 | 122 | 25.4 | 49.2 | 148 | 10.1 | 60.1 | 167 | 17.4 | 55.1 |
| M16 | 30,255 | 2 | 173 | 36.4 | 42.2 | 228 | 18.5 | 61.2 | 261 | 16.5 | 52.5 |
| L03 | 30,177 | 2 | 228 | 46.9 | 35.1 | 240 | 21.7 | 59.2 | 267 | 13.1 | 73.4 |
| B80 | 30,045 | 2 | 54 | 11.1 | 50.0 | 44 | 18.2 | 47.7 | 88 | 9.1 | 52.3 |
| A09 | 29,930 | 2 | 99 | 30.3 | 44.4 | 139 | 22.3 | 52.5 | 172 | 14.5 | 51.7 |
| N95 | 29,393 | 2 | 163 | 23.3 | 42.3 | 190 | 13.7 | 74.7 | 241 | 9.5 | 68.0 |
| M15 | 28,481 | 2 | 179 | 38.8 | 40.5 | 228 | 23.7 | 55.7 | 252 | 16.7 | 52.8 |
| G44 | 28,280 | 2 | 207 | 49.8 | 33.3 | 214 | 14.5 | 66.8 | 275 | 13.1 | 66.9 |
| S90 | 27,568 | 2 | 47 | 34.0 | 53.2 | 51 | 21.6 | 62.8 | 88 | 13.6 | 63.6 |
| F32 | 27,303 | 2 | 291 | 39.9 | 33.0 | 299 | 18.1 | 53.9 | 338 | 19.2 | 51.2 |
| K00 | 27,171 | 3 | 41 | 24.4 | 63.4 | 20 | 25.0 | 75.0 | 48 | 8.3 | 54.2 |
| I48 | 26,870 | 2 | 345 | 44.6 | 27.5 | 460 | 11.1 | 55.6 | 516 | 10.7 | 56.8 |
| S00 | 26,695 | 2 | 108 | 35.2 | 60.2 | 143 | 4.9 | 62.9 | 153 | 4.6 | 58.8 |
| L70 | 26,415 | 2 | 85 | 31.8 | 42.4 | 86 | 33.7 | 30.2 | 169 | 18.3 | 30.8 |
| L20 | 25,923 | 2 | 121 | 43.8 | 37.2 | 116 | 34.5 | 49.1 | 164 | 14.0 | 61.6 |
| B01 | 25,860 | 2 | 41 | 17.1 | 58.5 | 19 | 31.6 | 68.4 | 64 | 7.8 | 31.2 |
| J11 | 25,510 | 2 | 67 | 20.9 | 67.2 | 96 | 9.4 | 76.0 | 113 | 7.1 | 71.7 |
| A08 | 25,503 | 2 | 75 | 42.7 | 44.0 | 114 | 13.2 | 60.5 | 148 | 14.2 | 54.0 |
| J45 | 25,435 | 2 | 259 | 37.8 | 37.8 | 259 | 21.6 | 47.1 | 306 | 17.3 | 43.5 |
| R42 | 25,206 | 2 | 149 | 42.3 | 35.6 | 132 | 18.9 | 62.9 | 176 | 16.5 | 57.4 |
| H90 | 25,114 | 2 | 184 | 34.2 | 37.5 | 183 | 10.9 | 61.2 | 207 | 16.4 | 66.7 |
| N92 | 24,789 | 2 | 124 | 40.3 | 45.2 | 165 | 32.1 | 46.1 | 205 | 28.3 | 49.8 |
| M10 | 24,712 | 2 | 291 | 37.8 | 35.7 | 306 | 10.5 | 54.9 | 334 | 9.3 | 52.7 |
| L82 | 24,618 | 2 | 105 | 17.1 | 45.7 | 91 | 14.3 | 49.5 | 121 | 9.9 | 54.5 |
| G56 | 24,567 | 2 | 149 | 36.9 | 49.7 | 167 | 21.0 | 53.9 | 204 | 8.3 | 62.8 |
| B07 | 24,175 | 2 | 52 | 15.4 | 40.4 | 55 | 20.0 | 32.7 | 104 | 9.6 | 56.7 |
| J31 | 24,096 | 2 | 208 | 51.0 | 28.4 | 202 | 19.8 | 49.0 | 242 | 21.5 | 47.5 |
| M42 | 23,725 | 2 | 160 | 37.5 | 37.5 | 207 | 11.6 | 60.4 | 241 | 6.6 | 55.2 |
| H92 | 23,682 | 2 | 112 | 48.2 | 44.6 | 133 | 21.0 | 66.2 | 144 | 15.3 | 66.7 |
| M23 | 23,272 | 2 | 113 | 19.5 | 58.4 | 115 | 13.0 | 68.7 | 155 | 11.0 | 57.4 |
| S80 | 23,173 | 2 | 102 | 57.8 | 11.8 | 135 | 12.6 | 53.3 | 147 | 11.6 | 63.3 |
| I83 | 23,073 | 2 | 120 | 26.7 | 51.7 | 162 | 4.3 | 56.8 | 211 | 9.0 | 60.7 |
| M47 | 22,915 | 2 | 198 | 36.9 | 37.9 | 228 | 30.7 | 34.6 | 274 | 19.0 | 36.5 |
| N40 | 22,390 | 2 | 196 | 20.9 | 35.2 | 269 | 11.9 | 56.9 | 331 | 13.3 | 55.9 |
| K80 | 22,248 | 2 | 190 | 15.8 | 55.3 | 320 | 9.7 | 48.1 | 360 | 8.3 | 45.8 |
| S62 | 22,036 | 2 | 67 | 25.4 | 50.8 | 92 | 18.5 | 55.4 | 107 | 27.1 | 55.1 |
| Z32 | 21,951 | 2 | 108 | 58.3 | 26.9 | 134 | 41.8 | 53.7 | 222 | 46.0 | 48.6 |
| I25 | 21,708 | 2 | 308 | 41.2 | 36.4 | 416 | 25.7 | 45.0 | 435 | 26.9 | 48.5 |
| R52 | 21,705 | 2 | 285 | 54.0 | 29.5 | 283 | 40.6 | 48.8 | 283 | 36.0 | 51.2 |
| A69 | 21,491 | 2 | 110 | 25.4 | 59.1 | 115 | 13.0 | 71.3 | 145 | 11.7 | 60.7 |
| B00 | 21,156 | 2 | 92 | 18.5 | 65.2 | 105 | 14.3 | 74.3 | 116 | 18.1 | 56.9 |
| E11 | 20,983 | 2 | 223 | 40.8 | 26.5 | 297 | 27.3 | 50.8 | 308 | 17.2 | 57.5 |
| I20 | 20,917 | 2 | 225 | 56.0 | 31.2 | 309 | 25.6 | 54.0 | 372 | 23.9 | 52.4 |
| H40 | 20,565 | 4 | 164 | 31.7 | 37.8 | 145 | 24.8 | 55.2 | 183 | 19.7 | 61.8 |
| H11 | 20,534 | 3 | 126 | 23.0 | 43.6 | 101 | 4.0 | 62.4 | 131 | 16.0 | 48.9 |
| M41 | 20,432 | 2 | 124 | 17.7 | 63.7 | 110 | 4.6 | 71.8 | 142 | 5.6 | 78.9 |
| L60 | 20,253 | 2 | 94 | 35.1 | 53.2 | 95 | 34.7 | 62.1 | 110 | 19.1 | 63.6 |
| K01 | 20,247 | 2 | 50 | 58.0 | 24.0 | 43 | 16.3 | 79.1 | 67 | 11.9 | 62.7 |
| L24 | 20,244 | 2 | 122 | 18.9 | 53.3 | 110 | 20.0 | 51.8 | 146 | 18.5 | 44.5 |
| L01 | 20,145 | 2 | 109 | 33.0 | 48.6 | 124 | 24.2 | 47.6 | 140 | 23.6 | 35.0 |
| R11 | 20,143 | 3 | 214 | 45.3 | 39.7 | 254 | 17.7 | 67.3 | 274 | 10.6 | 69.3 |
| S20 | 19,972 | 2 | 77 | 26.0 | 66.2 | 111 | 24.3 | 55.9 | 119 | 3.4 | 77.3 |
| F51 | 19,619 | 2 | 274 | 17.5 | 31.8 | 210 | 19.5 | 37.6 | 271 | 9.6 | 41.7 |
| F43 | 19,559 | 2 | 185 | 42.2 | 37.8 | 184 | 8.2 | 68.5 | 220 | 8.6 | 70.9 |
| L50 | 19,427 | 2 | 127 | 36.2 | 41.7 | 131 | 21.4 | 51.9 | 148 | 29.0 | 50.7 |
| S52 | 19,409 | 2 | 88 | 39.8 | 40.9 | 146 | 20.6 | 55.5 | 141 | 15.6 | 66.0 |
| H01 | 19,400 | 2 | 112 | 32.1 | 35.7 | 99 | 32.3 | 32.3 | 114 | 21.9 | 46.5 |
| O80 | 19,194 | 2 | 124 | 16.1 | 41.9 | 123 | 6.5 | 69.9 | 145 | 16.6 | 68.3 |
| S92 | 18,903 | 2 | 67 | 22.4 | 61.2 | 84 | 17.9 | 69.0 | 93 | 19.4 | 52.7 |
| H00 | 18,763 | 2 | 64 | 34.4 | 35.9 | 56 | 37.5 | 32.1 | 66 | 30.3 | 43.9 |
| D25 | 18,612 | 2 | 151 | 23.8 | 47.7 | 195 | 11.6 | 55.8 | 244 | 14.8 | 53.7 |
| K58 | 18,595 | 2 | 143 | 30.1 | 51.0 | 161 | 6.2 | 72.7 | 201 | 5.5 | 54.2 |
| T15 | 18,041 | 2 | 34 | 61.8 | 38.2 | 45 | 15.6 | 73.3 | 57 | 42.1 | 40.4 |
| L21 | 17,817 | 2 | 93 | 26.9 | 48.4 | 96 | 20.8 | 60.4 | 143 | 11.9 | 55.2 |
| H66 | 17,660 | 3 | 118 | 45.8 | 39.8 | 151 | 32.5 | 43.7 | 140 | 29.3 | 49.3 |
| N18 | 17,522 | 2 | 291 | 35.6 | 41.9 | 460 | 12.0 | 65.4 | 411 | 15.8 | 61.8 |
| F45 | 17,516 | 2 | 185 | 35.1 | 51.4 | 200 | 7.5 | 68.5 | 223 | 6.7 | 78.0 |
| R04 | 17,469 | 2 | 164 | 37.8 | 42.7 | 221 | 21.3 | 55.2 | 250 | 18.4 | 54.0 |
| H91 | 17,209 | 2 | 108 | 36.1 | 41.7 | 153 | 17.6 | 55.6 | 163 | 21.5 | 49.7 |
| N64 | 17,157 | 2 | 69 | 24.6 | 40.6 | 94 | 24.5 | 55.3 | 125 | 30.4 | 51.2 |
| R06 | 17,014 | 2 | 180 | 42.8 | 35.6 | 247 | 34.0 | 49.0 | 279 | 30.1 | 49.8 |
| K08 | 16,962 | 2 | 38 | 81.6 | 10.5 | 33 | 12.1 | 87.9 | 45 | 6.7 | 68.9 |
| M13 | 16,506 | 2 | 128 | 52.3 | 32.8 | 175 | 34.9 | 53.7 | 198 | 34.9 | 54.5 |
| O99 | 16,466 | 2 | 197 | 61.4 | 26.4 | 210 | 31.0 | 61.4 | 209 | 29.2 | 63.2 |
| E04 | 16,254 | 2 | 160 | 11.9 | 39.4 | 190 | 6.8 | 57.9 | 221 | 12.2 | 39.8 |
| S83 | 16,177 | 2 | 75 | 26.7 | 57.3 | 83 | 41.0 | 49.4 | 126 | 17.5 | 57.1 |
| J10 | 15,610 | 2 | 127 | 20.5 | 44.1 | 188 | 8.0 | 65.4 | 178 | 14.0 | 60.1 |
| R73 | 15,328 | 2 | 74 | 25.7 | 50.0 | 182 | 17.6 | 57.1 | 183 | 7.7 | 63.4 |
| M20 | 15,288 | 2 | 82 | 32.9 | 45.1 | 104 | 3.9 | 76.9 | 149 | 8.1 | 65.8 |
| J44 | 15,281 | 2 | 252 | 26.2 | 53.2 | 331 | 20.2 | 61.6 | 387 | 18.9 | 58.7 |
| I70 | 15,148 | 2 | 221 | 33.9 | 26.2 | 305 | 28.5 | 35.7 | 324 | 26.5 | 43.2 |
| N41 | 15,115 | 2 | 135 | 33.3 | 43.7 | 144 | 17.4 | 55.6 | 235 | 18.7 | 46.0 |
| R00 | 15,042 | 2 | 123 | 30.9 | 48.8 | 209 | 11.0 | 72.7 | 263 | 11.8 | 52.9 |
| K76 | 14,874 | 2 | 190 | 43.2 | 43.2 | 277 | 22.4 | 64.3 | 302 | 22.9 | 64.6 |
| L72 | 14,856 | 2 | 74 | 25.7 | 37.8 | 70 | 10.0 | 65.7 | 87 | 10.3 | 64.4 |
| E03 | 14,792 | 2 | 158 | 24.0 | 52.5 | 224 | 11.2 | 66.5 | 259 | 12.0 | 58.1 |
| N83 | 14,608 | 2 | 120 | 30.8 | 40.8 | 160 | 17.5 | 48.8 | 195 | 19.0 | 63.1 |
| G43 | 14,463 | 2 | 109 | 36.7 | 48.6 | 128 | 18.0 | 53.9 | 167 | 12.6 | 54.5 |
| M50 | 14,422 | 5 | 130 | 45.4 | 29.2 | 152 | 30.3 | 39.5 | 169 | 13.0 | 50.3 |
| N84 | 14,252 | 2 | 152 | 23.7 | 48.7 | 153 | 13.7 | 41.2 | 212 | 16.0 | 65.1 |
| F10 | 13,962 | 2 | 159 | 22.6 | 66.7 | 262 | 16.0 | 78.6 | 311 | 21.2 | 70.1 |
| O70 | 13,878 | 2 | 111 | 20.7 | 57.7 | 129 | 9.3 | 59.7 | 125 | 9.6 | 50.4 |
| H04 | 13,833 | 2 | 92 | 35.9 | 35.9 | 81 | 28.4 | 37.0 | 80 | 5.0 | 62.5 |
| B97 | 13,691 | 2 | 104 | 23.1 | 48.1 | 111 | 26.1 | 48.6 | 129 | 24.0 | 48.8 |
| H43 | 13,676 | 2 | 108 | 28.7 | 60.2 | 100 | 20.0 | 60.0 | 98 | 6.1 | 77.6 |
| M48 | 13,621 | 2 | 177 | 37.3 | 40.7 | 199 | 18.6 | 57.3 | 238 | 27.3 | 43.3 |
| S82 | 13,613 | 2 | 87 | 40.2 | 34.5 | 136 | 27.2 | 59.6 | 162 | 35.2 | 54.9 |
| H26 | 13,586 | 2 | 107 | 30.8 | 50.5 | 112 | 18.8 | 56.2 | 113 | 13.3 | 66.4 |
| E06 | 13,543 | 2 | 141 | 27.0 | 44.7 | 162 | 14.8 | 53.7 | 197 | 12.7 | 57.9 |
| L40 | 13,523 | 2 | 77 | 32.5 | 61.0 | 85 | 30.6 | 56.5 | 118 | 29.7 | 46.6 |
| D17 | 13,431 | 2 | 70 | 18.6 | 35.7 | 97 | 20.6 | 48.5 | 120 | 19.2 | 51.7 |
| I80 | 13,411 | 2 | 204 | 36.3 | 39.2 | 232 | 15.5 | 58.2 | 266 | 14.3 | 64.3 |
| E79 | 13,311 | 2 | 184 | 29.9 | 45.6 | 259 | 4.2 | 80.3 | 250 | 8.0 | 73.2 |
| N20 | 13,231 | 2 | 129 | 41.9 | 31.8 | 216 | 21.3 | 49.5 | 232 | 13.8 | 48.3 |
| R50 | 13,209 | 2 | 107 | 37.4 | 50.5 | 158 | 28.5 | 62.0 | 174 | 28.2 | 54.6 |
| S63 | 13,012 | 2 | 38 | 36.8 | 55.3 | 52 | 34.6 | 51.9 | 79 | 27.9 | 51.9 |
| M65 | 12,994 | 2 | 100 | 55.0 | 32.0 | 110 | 33.6 | 50.0 | 115 | 31.3 | 51.3 |
| R55 | 12,910 | 2 | 130 | 40.8 | 43.9 | 206 | 15.5 | 61.6 | 229 | 10.9 | 66.8 |
| H53 | 12,869 | 2 | 143 | 53.9 | 37.8 | 151 | 47.0 | 38.4 | 178 | 42.7 | 43.3 |
| N94 | 12,837 | 2 | 76 | 35.5 | 46.0 | 110 | 48.2 | 40.0 | 136 | 33.8 | 45.6 |
| M24 | 12,764 | 2 | 157 | 37.6 | 48.4 | 129 | 21.7 | 72.9 | 143 | 18.2 | 64.3 |
| M67 | 12,734 | 2 | 74 | 27.0 | 56.8 | 78 | 10.3 | 65.4 | 99 | 12.1 | 60.6 |
| B02 | 12,669 | 2 | 96 | 22.9 | 59.4 | 114 | 21.0 | 60.5 | 107 | 22.4 | 62.6 |
| J96 | 12,479 | 2 | 292 | 50.7 | 33.6 | 302 | 8.3 | 62.2 | 286 | 12.2 | 65.7 |
| L08 | 12,466 | 2 | 135 | 48.1 | 35.6 | 159 | 36.5 | 44.0 | 155 | 31.6 | 51.0 |
| A63 | 12,456 | 2 | 96 | 41.7 | 45.8 | 107 | 40.2 | 43.9 | 151 | 24.5 | 49.7 |
| S50 | 12,163 | 2 | 69 | 34.8 | 56.5 | 89 | 13.5 | 57.3 | 99 | 13.1 | 53.5 |
| R03 | 11,933 | 2 | 72 | 29.6 | 52.1 | 130 | 29.2 | 51.5 | 152 | 24.3 | 44.1 |
| F33 | 11,914 | 2 | 154 | 43.5 | 36.4 | 159 | 26.4 | 57.2 | 171 | 31.0 | 53.2 |
| B36 | 11,852 | 2 | 67 | 26.9 | 43.3 | 81 | 24.7 | 37.0 | 87 | 31.0 | 33.3 |
| I63 | 11,601 | 4 | 166 | 38.0 | 33.1 | 308 | 29.2 | 52.9 | 331 | 20.5 | 61.3 |
| I69 | 11,336 | 2 | 269 | 45.4 | 38.3 | 320 | 23.1 | 61.9 | 327 | 27.8 | 62.4 |
| T63 | 11,208 | 2 | 30 | 26.7 | 63.3 | 52 | 32.7 | 55.8 | 53 | 5.7 | 67.9 |
| L71 | 11,152 | 2 | 70 | 24.3 | 48.6 | 85 | 20.0 | 42.4 | 87 | 21.8 | 43.7 |
| J15 | 11,141 | 2 | 255 | 37.6 | 33.3 | 351 | 13.4 | 59.0 | 344 | 12.8 | 62.8 |
| E53 | 11,102 | 2 | 193 | 21.2 | 54.9 | 331 | 9.7 | 51.7 | 296 | 8.1 | 58.1 |
| S30 | 11,068 | 2 | 81 | 23.5 | 43.2 | 125 | 8.0 | 54.4 | 129 | 6.2 | 66.7 |
| S22 | 11,064 | 2 | 112 | 41.1 | 50.0 | 208 | 28.9 | 54.8 | 237 | 23.2 | 60.8 |
| K25 | 10,943 | 2 | 237 | 21.5 | 37.1 | 291 | 18.6 | 46.0 | 348 | 13.8 | 43.4 |
| S42 | 10,871 | 2 | 87 | 64.4 | 24.1 | 156 | 25.6 | 62.2 | 169 | 23.1 | 62.7 |
| N31 | 10,584 | 2 | 145 | 29.7 | 47.6 | 147 | 20.4 | 56.5 | 154 | 17.5 | 64.3 |
| D63 | 10,452 | 2 | 367 | 49.3 | 38.7 | 441 | 21.1 | 64.6 | 460 | 23.7 | 64.1 |
| L84 | 10,381 | 2 | 50 | 12.0 | 56.0 | 49 | 6.1 | 67.3 | 62 | 4.8 | 64.5 |
| S05 | 10,378 | 2 | 62 | 35.5 | 51.6 | 83 | 33.7 | 56.6 | 88 | 56.8 | 29.6 |
| A49 | 10,259 | 2 | 269 | 44.2 | 29.0 | 324 | 15.4 | 64.2 | 305 | 24.3 | 57.4 |
| H68 | 10,208 | 2 | 113 | 38.0 | 51.3 | 117 | 17.1 | 67.5 | 112 | 21.4 | 67.9 |
| S02 | 10,131 | 2 | 106 | 53.8 | 34.0 | 175 | 16.6 | 71.4 | 177 | 26.0 | 63.8 |
| R22 | 10,077 | 2 | 35 | 28.6 | 42.9 | 95 | 31.6 | 47.4 | 109 | 32.1 | 49.5 |
| I67 | 10,044 | 2 | 140 | 37.1 | 29.3 | 211 | 12.8 | 64.0 | 220 | 12.7 | 72.3 |
| S40 | 10,034 | 2 | 66 | 19.7 | 69.7 | 90 | 6.7 | 53.3 | 111 | 8.1 | 63.1 |
| K57 | 10,010 | 2 | 169 | 22.5 | 47.3 | 210 | 9.1 | 59.5 | 242 | 10.7 | 50.8 |
| F06 | 9,971 | 2 | 238 | 40.3 | 48.3 | 260 | 28.9 | 65.0 | 267 | 38.6 | 50.2 |
| G58 | 9,962 | 2 | 83 | 34.9 | 56.6 | 124 | 11.3 | 62.9 | 118 | 9.3 | 72.0 |
| D64 | 9,925 | 2 | 227 | 32.6 | 42.7 | 370 | 12.4 | 64.0 | 400 | 7.2 | 67.8 |
| J32 | 9,818 | 2 | 157 | 44.0 | 40.8 | 165 | 25.4 | 52.1 | 169 | 14.2 | 58.0 |
| D48 | 9,710 | 2 | 102 | 33.3 | 52.9 | 148 | 21.6 | 74.3 | 200 | 21.0 | 66.0 |
| D51 | 9,707 | 3 | 155 | 21.9 | 54.2 | 214 | 9.3 | 50.5 | 203 | 11.8 | 60.6 |
| L98 | 9,690 | 2 | 145 | 46.2 | 42.1 | 172 | 25.0 | 55.2 | 193 | 21.2 | 61.7 |
| N87 | 9,674 | 3 | 93 | 15.1 | 54.8 | 104 | 19.2 | 56.7 | 166 | 12.7 | 50.0 |
| S81 | 9,576 | 2 | 63 | 39.7 | 50.8 | 93 | 35.5 | 50.5 | 89 | 32.6 | 52.8 |
| N50 | 9,500 | 4 | 83 | 51.8 | 31.3 | 85 | 27.1 | 65.9 | 129 | 29.5 | 58.9 |
| I13 | 9,469 | 2 | 244 | 47.9 | 31.9 | 344 | 25.3 | 45.6 | 316 | 21.2 | 60.8 |
| N28 | 9,380 | 2 | 127 | 35.4 | 43.3 | 208 | 20.2 | 52.9 | 215 | 12.1 | 68.8 |
| D18 | 9,379 | 2 | 78 | 30.8 | 50.0 | 97 | 15.5 | 57.7 | 92 | 22.8 | 60.9 |
| K12 | 9,371 | 2 | 75 | 17.3 | 66.7 | 84 | 9.5 | 76.2 | 91 | 16.5 | 56.0 |
| S06 | 9,353 | 2 | 100 | 29.0 | 52.0 | 182 | 17.6 | 65.4 | 198 | 14.1 | 67.7 |
| B33 | 9,251 | 2 | 39 | 25.6 | 69.2 | 58 | 19.0 | 72.4 | 71 | 7.0 | 83.1 |
| R47 | 9,204 | 2 | 157 | 27.4 | 53.5 | 213 | 22.5 | 51.2 | 244 | 17.6 | 57.4 |
| N48 | 9,169 | 2 | 95 | 27.4 | 54.7 | 92 | 31.5 | 54.4 | 122 | 26.2 | 64.8 |
| N60 | 9,048 | 2 | 57 | 31.6 | 45.6 | 63 | 25.4 | 42.9 | 102 | 18.6 | 48.0 |
| M72 | 8,998 | 2 | 66 | 18.2 | 63.6 | 82 | 14.6 | 62.2 | 88 | 15.9 | 62.5 |
| E28 | 8,991 | 2 | 79 | 38.0 | 55.7 | 103 | 29.1 | 45.6 | 141 | 29.8 | 45.4 |
| E87 | 8,825 | 2 | 277 | 40.4 | 28.5 | 251 | 13.9 | 43.0 | 349 | 21.8 | 34.7 |
| O26 | 8,811 | 2 | 123 | 47.1 | 52.9 | 145 | 38.6 | 53.1 | 166 | 44.6 | 51.2 |
| K40 | 8,786 | 2 | 75 | 29.3 | 45.3 | 123 | 5.7 | 69.1 | 146 | 10.3 | 64.4 |
| O23 | 8,770 | 3 | 115 | 24.4 | 39.1 | 150 | 17.3 | 42.0 | 177 | 13.0 | 44.6 |
| N72 | 8,707 | 2 | 114 | 28.1 | 43.0 | 116 | 25.0 | 50.0 | 142 | 28.9 | 45.1 |
| M43 | 8,701 | 3 | 104 | 26.0 | 71.2 | 130 | 16.9 | 55.4 | 133 | 13.5 | 72.2 |
| S91 | 8,664 | 2 | 40 | 55.0 | 42.5 | 54 | 48.1 | 50.0 | 60 | 36.7 | 55.0 |
| M76 | 8,563 | 2 | 82 | 46.3 | 41.5 | 88 | 36.4 | 40.9 | 76 | 11.8 | 68.4 |
| G50 | 8,505 | 2 | 108 | 19.4 | 53.7 | 127 | 11.8 | 61.4 | 124 | 10.5 | 39.5 |
| N91 | 8,452 | 2 | 44 | 22.7 | 65.9 | 68 | 26.5 | 38.2 | 90 | 30.0 | 40.0 |
| R21 | 8,430 | 2 | 66 | 36.4 | 45.5 | 86 | 37.2 | 50.0 | 79 | 41.8 | 41.8 |
| D21 | 8,427 | 2 | 65 | 21.5 | 43.1 | 66 | 21.2 | 53.0 | 92 | 13.0 | 62.0 |
| D12 | 8,387 | 2 | 153 | 28.1 | 45.8 | 179 | 14.0 | 52.0 | 222 | 14.0 | 58.6 |
| K26 | 8,221 | 2 | 153 | 38.6 | 41.8 | 196 | 17.9 | 51.5 | 231 | 19.9 | 50.6 |
| L97 | 8,220 | 2 | 180 | 26.7 | 56.7 | 199 | 20.1 | 62.3 | 228 | 24.6 | 60.1 |
| F48 | 8,184 | 2 | 123 | 21.1 | 49.6 | 120 | 23.3 | 69.2 | 142 | 23.2 | 54.9 |
| I87 | 8,166 | 2 | 101 | 31.7 | 51.5 | 138 | 10.9 | 63.8 | 144 | 16.0 | 69.4 |
| G45 | 8,125 | 2 | 115 | 33.9 | 54.8 | 212 | 19.3 | 55.2 | 207 | 26.6 | 50.7 |
| J12 | 8,097 | 2 | 119 | 23.5 | 56.3 | 202 | 9.9 | 69.8 | 210 | 15.2 | 66.2 |
| L85 | 8,045 | 2 | 60 | 40.0 | 48.3 | 74 | 12.2 | 62.2 | 74 | 29.7 | 47.3 |
| A41 | 8,041 | 2 | 319 | 53.3 | 36.7 | 243 | 16.5 | 64.2 | 257 | 14.0 | 72.0 |
| B08 | 7,997 | 2 | 46 | 21.7 | 71.7 | 58 | 19.0 | 72.4 | 42 | 19.0 | 57.1 |
| H02 | 7,958 | 2 | 87 | 17.2 | 48.3 | 84 | 23.8 | 48.8 | 88 | 22.7 | 52.3 |
| N10 | 7,930 | 2 | 140 | 33.6 | 47.9 | 179 | 14.5 | 60.3 | 187 | 9.1 | 57.2 |
| F90 | 7,850 | 2 | 91 | 45.0 | 50.5 | 88 | 20.4 | 52.3 | 93 | 16.1 | 68.8 |
| N81 | 7,800 | 2 | 87 | 24.1 | 46.0 | 128 | 14.1 | 52.3 | 140 | 11.4 | 58.6 |
| I21 | 7,733 | 5 | 172 | 40.1 | 38.4 | 304 | 16.1 | 54.0 | 334 | 27.2 | 42.8 |
| K35 | 7,711 | 2 | 83 | 12.1 | 55.4 | 144 | 9.0 | 64.6 | 136 | 27.9 | 60.3 |
| F80 | 7,703 | 3 | 64 | 40.6 | 34.4 | 68 | 14.7 | 60.3 | 106 | 16.0 | 60.4 |
| M22 | 7,586 | 2 | 73 | 32.9 | 57.5 | 63 | 14.3 | 69.8 | 74 | 14.9 | 63.5 |
| A46 | 7,499 | 2 | 122 | 41.8 | 41.0 | 170 | 15.3 | 64.1 | 176 | 18.2 | 61.4 |
| L25 | 7,473 | 2 | 71 | 26.8 | 54.9 | 81 | 21.0 | 54.3 | 82 | 25.6 | 57.3 |
| L89 | 7,462 | 2 | 270 | 64.4 | 29.3 | 250 | 7.6 | 74.4 | 192 | 12.5 | 72.4 |
| L27 | 7,449 | 2 | 116 | 37.1 | 48.3 | 107 | 30.8 | 38.3 | 110 | 28.2 | 58.2 |
| O62 | 7,435 | 2 | 109 | 21.1 | 55.0 | 122 | 13.1 | 66.4 | 142 | 9.9 | 45.1 |
| O36 | 7,417 | 2 | 123 | 41.5 | 48.0 | 135 | 22.2 | 68.2 | 151 | 14.6 | 71.5 |
| S43 | 7,397 | 2 | 48 | 41.7 | 43.8 | 87 | 40.2 | 48.3 | 109 | 12.8 | 60.5 |
| R30 | 7,316 | 2 | 79 | 41.8 | 43.0 | 98 | 30.6 | 51.0 | 108 | 13.0 | 67.6 |
| K60 | 7,294 | 2 | 128 | 35.2 | 39.8 | 97 | 15.5 | 57.7 | 121 | 15.7 | 63.6 |
| L29 | 7,193 | 2 | 97 | 48.5 | 39.2 | 104 | 53.9 | 39.4 | 100 | 46.0 | 48.0 |
| F01 | 7,143 | 2 | 188 | 31.9 | 47.9 | 219 | 14.2 | 75.3 | 214 | 13.1 | 72.9 |
| R32 | 7,127 | 2 | 82 | 31.7 | 47.6 | 113 | 26.6 | 52.2 | 120 | 19.2 | 63.3 |
| M71 | 7,081 | 2 | 84 | 38.1 | 41.7 | 101 | 28.7 | 48.5 | 103 | 20.4 | 61.2 |
| G40 | 7,072 | 2 | 179 | 43.6 | 30.7 | 241 | 27.0 | 58.9 | 256 | 27.0 | 64.1 |
| M53 | 7,065 | 2 | 94 | 44.7 | 36.2 | 91 | 36.3 | 47.2 | 103 | 12.6 | 52.4 |
| M80 | 7,017 | 2 | 157 | 21.7 | 58.6 | 177 | 15.2 | 72.3 | 162 | 25.9 | 62.4 |
| K44 | 6,995 | 2 | 136 | 27.9 | 50.0 | 203 | 15.3 | 47.3 | 216 | 13.4 | 47.7 |
| S70 | 6,948 | 2 | 69 | 36.2 | 47.8 | 118 | 11.0 | 67.0 | 115 | 5.2 | 68.7 |
| J39 | 6,941 | 2 | 98 | 39.8 | 51.0 | 129 | 23.3 | 58.9 | 134 | 23.9 | 56.0 |
| N97 | 6,827 | 2 | 75 | 54.7 | 26.7 | 103 | 49.5 | 37.9 | 213 | 30.5 | 40.4 |
| G62 | 6,800 | 2 | 149 | 26.2 | 50.3 | 183 | 24.6 | 56.8 | 206 | 22.3 | 62.6 |
| J40 | 6,732 | 2 | 122 | 41.8 | 41.0 | 169 | 23.1 | 43.2 | 174 | 16.7 | 59.2 |
| N85 | 6,685 | 2 | 134 | 32.1 | 35.1 | 143 | 27.3 | 51.0 | 187 | 26.7 | 54.5 |
| I47 | 6,623 | 2 | 127 | 39.4 | 40.2 | 199 | 15.1 | 62.3 | 215 | 17.2 | 53.0 |
| L04 | 6,576 | 2 | 86 | 60.5 | 33.7 | 134 | 27.6 | 59.0 | 148 | 23.0 | 60.1 |
| D27 | 6,466 | 2 | 122 | 23.8 | 47.5 | 130 | 9.2 | 72.3 | 136 | 11.8 | 69.8 |
| L81 | 6,402 | 2 | 56 | 25.0 | 57.1 | 62 | 27.4 | 43.5 | 70 | 25.7 | 47.1 |
| K86 | 6,307 | 2 | 133 | 24.1 | 63.2 | 180 | 16.1 | 63.3 | 210 | 14.8 | 73.3 |
| S72 | 6,296 | 2 | 119 | 57.1 | 32.8 | 200 | 21.0 | 64.5 | 204 | 31.4 | 55.4 |
| E05 | 6,286 | 2 | 111 | 19.8 | 57.7 | 151 | 15.1 | 56.6 | 162 | 19.1 | 61.1 |
| E73 | 6,285 | 2 | 99 | 6.1 | 75.8 | 119 | 5.0 | 51.3 | 108 | 3.7 | 52.8 |
| R60 | 6,183 | 2 | 95 | 36.8 | 54.7 | 121 | 24.8 | 53.7 | 127 | 29.1 | 60.6 |
| G54 | 6,166 | 2 | 91 | 19.8 | 67.0 | 113 | 14.2 | 62.0 | 110 | 11.8 | 64.6 |
| L73 | 6,088 | 2 | 75 | 61.3 | 28.0 | 76 | 32.9 | 55.3 | 77 | 48.0 | 36.4 |
| N63 | 6,088 | 2 | 40 | 32.5 | 40.0 | 91 | 42.9 | 48.4 | 117 | 44.4 | 49.6 |
| N80 | 6,029 | 3 | 123 | 27.6 | 52.0 | 131 | 19.1 | 38.2 | 161 | 23.6 | 52.8 |
| O20 | 6,020 | 2 | 103 | 49.5 | 34.0 | 108 | 28.7 | 49.1 | 197 | 21.8 | 34.5 |
| K52 | 5,966 | 2 | 127 | 30.7 | 55.9 | 154 | 18.2 | 63.6 | 159 | 20.8 | 66.0 |
| B86 | 5,939 | 2 | 88 | 33.0 | 39.8 | 90 | 8.9 | 63.3 | 87 | 11.5 | 67.8 |
| G25 | 5,933 | 2 | 87 | 31.0 | 59.8 | 114 | 10.5 | 47.4 | 123 | 17.9 | 46.3 |
| D52 | 5,927 | 2 | 194 | 11.9 | 42.3 | 260 | 6.2 | 70.0 | 233 | 8.6 | 64.4 |
| O42 | 5,914 | 2 | 120 | 20.0 | 49.2 | 126 | 15.9 | 58.7 | 136 | 16.2 | 42.6 |
| N17 | 5,902 | 2 | 246 | 49.6 | 41.9 | 237 | 6.3 | 69.6 | 235 | 9.8 | 55.3 |
| T81 | 5,864 | 2 | 306 | 68.3 | 30.1 | 254 | 35.0 | 40.2 | 281 | 35.2 | 44.5 |
| N11 | 5,835 | 2 | 145 | 44.8 | 42.8 | 182 | 23.6 | 62.1 | 165 | 12.1 | 75.2 |
| L57 | 5,796 | 2 | 70 | 28.6 | 45.7 | 78 | 30.8 | 52.6 | 66 | 24.2 | 57.6 |
| S51 | 5,794 | 2 | 58 | 72.4 | 20.7 | 87 | 28.7 | 51.7 | 85 | 18.8 | 61.2 |
| R31 | 5,761 | 2 | 119 | 37.8 | 47.1 | 179 | 32.4 | 49.7 | 214 | 39.2 | 42.5 |
| M81 | 5,705 | 2 | 101 | 14.8 | 68.3 | 128 | 7.8 | 51.6 | 117 | 12.0 | 54.7 |
| M06 | 5,695 | 2 | 137 | 47.5 | 45.3 | 140 | 17.9 | 60.0 | 160 | 18.1 | 67.5 |
| N90 | 5,666 | 2 | 107 | 27.1 | 60.8 | 105 | 31.4 | 56.2 | 101 | 33.7 | 49.5 |
| C44 | 5,649 | 2 | 68 | 33.8 | 54.4 | 86 | 18.6 | 62.8 | 92 | 32.6 | 63.0 |
| O47 | 5,557 | 3 | 104 | 17.3 | 64.4 | 112 | 7.1 | 52.7 | 136 | 5.9 | 55.9 |
| K62 | 5,466 | 2 | 110 | 22.7 | 53.6 | 141 | 19.1 | 59.6 | 155 | 21.9 | 56.8 |
| R87 | 5,384 | 2 | 50 | 20.0 | 64.0 | 54 | 22.2 | 64.8 | 74 | 18.9 | 66.2 |
| O71 | 5,379 | 3 | 107 | 22.4 | 46.7 | 114 | 14.9 | 56.1 | 105 | 16.2 | 43.8 |
| M60 | 5,318 | 2 | 75 | 16.0 | 69.3 | 95 | 9.5 | 69.5 | 88 | 11.4 | 75.0 |
| M94 | 5,318 | 2 | 87 | 27.6 | 49.4 | 57 | 24.6 | 66.7 | 63 | 17.5 | 69.8 |
| N47 | 5,281 | 2 | 79 | 20.2 | 60.8 | 85 | 16.5 | 61.2 | 86 | 4.7 | 70.9 |
| K92 | 5,246 | 2 | 152 | 42.1 | 38.8 | 217 | 26.3 | 55.3 | 206 | 48.1 | 35.4 |
| O24 | 5,181 | 2 | 98 | 17.4 | 81.6 | 92 | 17.4 | 70.7 | 166 | 10.8 | 70.5 |
| I35 | 5,127 | 2 | 137 | 12.4 | 59.9 | 220 | 9.1 | 75.0 | 250 | 8.0 | 72.0 |
| K43 | 5,095 | 2 | 77 | 16.9 | 49.4 | 109 | 9.2 | 82.6 | 91 | 17.6 | 61.5 |
| R59 | 5,087 | 2 | 75 | 37.3 | 49.3 | 114 | 21.9 | 50.0 | 146 | 28.1 | 58.2 |
| I26 | 5,081 | 2 | 230 | 35.2 | 45.6 | 281 | 10.0 | 54.8 | 258 | 11.6 | 67.4 |
| K42 | 5,067 | 2 | 69 | 18.8 | 63.8 | 119 | 6.7 | 74.0 | 126 | 7.9 | 63.5 |
| N86 | 5,054 | 2 | 31 | 29.0 | 64.5 | 79 | 17.7 | 51.9 | 89 | 13.5 | 57.3 |
| A56 | 5,040 | 2 | 70 | 34.3 | 45.7 | 69 | 21.7 | 68.1 | 120 | 16.7 | 50.0 |
| R33 | 5,017 | 2 | 164 | 31.1 | 43.3 | 218 | 14.2 | 56.4 | 232 | 22.4 | 53.5 |
| H62 | 5,012 | 2 | 64 | 29.7 | 48.4 | 74 | 24.3 | 58.1 | 58 | 20.7 | 63.8 |
| T51 | 5,002 | 2 | 80 | 27.5 | 56.2 | 132 | 12.1 | 72.0 | 142 | 11.3 | 76.8 |
| M85 | 4,919 | 2 | 87 | 20.7 | 55.2 | 65 | 7.7 | 69.2 | 62 | 14.5 | 59.7 |
| S32 | 4,905 | 2 | 102 | 21.6 | 59.8 | 179 | 22.9 | 67.6 | 192 | 28.1 | 64.1 |
| G81 | 4,901 | 3 | 173 | 50.9 | 33.0 | 219 | 15.1 | 68.0 | 215 | 24.6 | 63.3 |
| J42 | 4,881 | 2 | 92 | 29.4 | 54.4 | 117 | 20.5 | 67.5 | 144 | 18.8 | 65.3 |
| O34 | 4,872 | 2 | 90 | 17.8 | 52.2 | 146 | 19.2 | 65.1 | 173 | 11.6 | 64.7 |
| N42 | 4,854 | 2 | 82 | 30.5 | 36.6 | 88 | 30.7 | 54.5 | 152 | 27.0 | 59.9 |
| R68 | 4,804 | 2 | 38 | 44.7 | 39.5 | 59 | 1.7 | 83.0 | 57 | 1.8 | 80.7 |
| K81 | 4,774 | 2 | 121 | 18.2 | 67.8 | 160 | 15.6 | 54.4 | 157 | 27.4 | 52.9 |
| R12 | 4,745 | 2 | 91 | 22.0 | 52.8 | 92 | 18.5 | 65.2 | 95 | 15.8 | 47.4 |
| R49 | 4,722 | 2 | 127 | 41.0 | 47.0 | 101 | 15.8 | 60.4 | 118 | 31.4 | 52.5 |
| F52 | 4,653 | 3 | 57 | 52.6 | 40.4 | 55 | 41.8 | 47.3 | 69 | 37.7 | 49.3 |
| B27 | 4,645 | 2 | 83 | 27.7 | 59.0 | 103 | 17.5 | 61.2 | 98 | 8.2 | 64.3 |
| T14 | 4,635 | 2 | 34 | 32.4 | 58.8 | 50 | 22.0 | 66.0 | 35 | 20.0 | 68.6 |
| H16 | 4,619 | 2 | 57 | 59.6 | 35.1 | 79 | 41.8 | 36.7 | 73 | 39.7 | 45.2 |
| E61 | 4,594 | 2 | 66 | 24.2 | 48.5 | 113 | 14.2 | 57.5 | 85 | 16.5 | 43.5 |
| J91 | 4,569 | 4 | 226 | 50.9 | 35.0 | 321 | 19.3 | 57.3 | 308 | 12.7 | 67.9 |
| Z33 | 4,561 | 2 | 28 | 39.3 | 32.1 | 72 | 38.9 | 52.8 | 82 | 31.7 | 63.4 |
| O02 | 4,544 | 2 | 90 | 47.8 | 46.7 | 98 | 32.6 | 54.1 | 134 | 26.1 | 51.5 |
| J41 | 4,496 | 2 | 93 | 39.8 | 41.9 | 117 | 27.4 | 65.8 | 116 | 13.8 | 69.8 |
| R14 | 4,440 | 2 | 55 | 18.2 | 54.5 | 88 | 10.2 | 69.3 | 75 | 9.3 | 61.3 |
| B77 | 4,316 | 5 | 54 | 25.9 | 57.4 | 46 | 13.0 | 58.7 | 42 | 9.5 | 76.2 |
| A04 | 4,283 | 2 | 153 | 25.5 | 48.4 | 168 | 7.3 | 60.3 | 181 | 17.1 | 50.3 |
| A60 | 4,260 | 2 | 56 | 17.9 | 71.4 | 64 | 15.6 | 51.6 | 53 | 17.0 | 56.6 |
| M02 | 4,246 | 2 | 95 | 52.6 | 44.2 | 119 | 33.6 | 49.6 | 133 | 19.6 | 58.6 |
| H18 | 4,236 | 3 | 82 | 30.5 | 51.2 | 78 | 28.2 | 57.7 | 79 | 24.0 | 57.0 |
| E07 | 4,224 | 2 | 53 | 15.1 | 66.0 | 90 | 11.1 | 65.6 | 88 | 14.8 | 71.6 |
| R01 | 4,182 | 2 | 41 | 12.2 | 73.2 | 64 | 10.9 | 73.4 | 68 | 14.7 | 54.4 |
| I34 | 4,161 | 2 | 120 | 25.0 | 55.8 | 206 | 18.9 | 62.1 | 214 | 23.8 | 63.5 |
| K20 | 4,150 | 2 | 108 | 25.9 | 50.9 | 162 | 15.4 | 51.9 | 166 | 14.5 | 54.2 |
| G93 | 4,146 | 2 | 137 | 54.0 | 34.3 | 190 | 18.4 | 70.0 | 170 | 17.8 | 68.0 |
| R13 | 4,126 | 2 | 168 | 39.9 | 45.8 | 210 | 25.7 | 50.5 | 201 | 25.4 | 54.2 |
| K22 | 4,121 | 2 | 125 | 28.8 | 47.2 | 165 | 18.8 | 57.6 | 174 | 15.5 | 67.2 |
| F98 | 4,119 | 2 | 51 | 43.1 | 51.0 | 68 | 17.6 | 51.5 | 72 | 25.0 | 50.0 |
| M40 | 4,085 | 2 | 54 | 35.2 | 50.0 | 61 | 8.2 | 72.1 | 62 | 8.1 | 71.0 |
| J22 | 4,067 | 2 | 68 | 38.2 | 54.4 | 117 | 30.8 | 53.9 | 94 | 26.6 | 62.8 |
| B96 | 4,045 | 2 | 114 | 51.8 | 39.5 | 161 | 24.2 | 51.5 | 118 | 22.9 | 59.3 |
| M92 | 4,042 | 2 | 37 | 35.1 | 56.8 | 38 | 29.0 | 47.4 | 53 | 15.1 | 58.5 |
| I42 | 4,002 | 2 | 122 | 41.8 | 48.4 | 203 | 22.2 | 56.6 | 211 | 22.8 | 64.4 |
| H54 | 3,975 | 3 | 75 | 52.0 | 41.3 | 98 | 46.9 | 36.7 | 82 | 45.1 | 47.6 |
| K56 | 3,965 | 2 | 167 | 44.3 | 46.7 | 233 | 15.4 | 67.0 | 230 | 17.0 | 69.6 |
| T88 | 3,940 | 2 | 142 | 54.9 | 38.0 | 134 | 28.4 | 53.7 | 154 | 24.0 | 59.1 |
| O68 | 3,934 | 2 | 104 | 30.8 | 55.8 | 113 | 12.4 | 51.3 | 118 | 11.9 | 43.2 |
| F03 | 3,905 | 2 | 126 | 37.3 | 45.2 | 174 | 9.8 | 78.7 | 169 | 21.9 | 66.3 |
| F40 | 3,902 | 2 | 69 | 30.4 | 53.6 | 73 | 32.9 | 61.6 | 95 | 21.0 | 57.9 |
| K85 | 3,881 | 2 | 115 | 20.0 | 60.9 | 189 | 15.9 | 66.1 | 204 | 15.7 | 66.7 |
| R29 | 3,880 | 2 | 39 | 30.8 | 59.0 | 54 | 22.2 | 57.4 | 57 | 15.8 | 71.9 |
| I44 | 3,877 | 2 | 112 | 30.4 | 57.1 | 185 | 7.0 | 61.6 | 174 | 7.5 | 70.7 |
| M62 | 3,859 | 2 | 60 | 25.0 | 71.7 | 93 | 8.6 | 88.2 | 77 | 9.1 | 85.7 |
| G63 | 3,815 | 2 | 119 | 31.1 | 63.9 | 156 | 27.6 | 57.0 | 148 | 25.7 | 55.4 |
| M35 | 3,805 | 2 | 118 | 36.4 | 59.3 | 121 | 28.1 | 69.4 | 137 | 20.4 | 75.2 |
| R23 | 3,803 | 2 | 36 | 13.9 | 75.0 | 51 | 11.8 | 80.4 | 52 | 19.2 | 75.0 |
| T16 | 3,799 | 2 | 42 | 40.5 | 38.1 | 43 | 9.3 | 65.1 | 38 | 10.5 | 63.2 |
| N46 | 3,793 | 4 | 36 | 47.2 | 47.2 | 40 | 25.0 | 50.0 | 66 | 18.2 | 56.1 |
| D69 | 3,737 | 2 | 165 | 42.4 | 37.0 | 196 | 22.4 | 62.2 | 211 | 17.5 | 66.3 |
| K70 | 3,690 | 2 | 119 | 29.4 | 53.8 | 208 | 19.2 | 62.0 | 224 | 20.5 | 70.5 |
| H36 | 3,662 | 2 | 89 | 27.0 | 46.1 | 81 | 23.5 | 55.6 | 86 | 29.1 | 52.3 |
| N88 | 3,649 | 3 | 88 | 23.9 | 62.5 | 100 | 28.0 | 55.0 | 105 | 17.1 | 72.4 |
| N34 | 3,604 | 2 | 68 | 38.2 | 45.6 | 72 | 29.2 | 55.6 | 82 | 24.4 | 58.5 |
| H50 | 3,550 | 2 | 57 | 47.4 | 47.4 | 43 | 30.2 | 58.1 | 51 | 29.4 | 58.8 |
| N93 | 3,529 | 2 | 89 | 39.3 | 48.3 | 103 | 46.6 | 42.7 | 111 | 40.5 | 51.4 |
| G57 | 3,524 | 2 | 92 | 26.1 | 55.4 | 95 | 17.9 | 63.2 | 97 | 19.6 | 70.1 |
| K31 | 3,523 | 2 | 122 | 24.6 | 42.6 | 161 | 16.1 | 60.9 | 173 | 17.3 | 67.0 |
| K13 | 3,478 | 2 | 53 | 41.5 | 39.6 | 63 | 19.0 | 66.7 | 48 | 27.1 | 60.4 |
| J33 | 3,473 | 2 | 91 | 38.5 | 50.5 | 100 | 19.0 | 59.0 | 121 | 17.4 | 56.2 |
| B18 | 3,459 | 2 | 99 | 22.2 | 54.5 | 121 | 19.8 | 66.9 | 159 | 22.6 | 64.8 |
| N89 | 3,421 | 2 | 67 | 23.9 | 58.2 | 75 | 32.0 | 61.3 | 66 | 33.3 | 59.1 |
| J05 | 3,385 | 3 | 43 | 30.2 | 48.8 | 70 | 7.1 | 75.7 | 59 | 17.0 | 55.9 |
| N45 | 3,373 | 3 | 91 | 39.6 | 42.9 | 88 | 30.7 | 60.2 | 96 | 27.1 | 58.3 |
| I65 | 3,314 | 2 | 118 | 16.7 | 66.7 | 177 | 13.0 | 61.6 | 189 | 12.7 | 68.2 |
| K11 | 3,281 | 2 | 59 | 32.2 | 59.3 | 81 | 23.5 | 53.1 | 80 | 13.8 | 55.0 |
| O28 | 3,269 | 2 | 91 | 68.1 | 27.5 | 110 | 38.2 | 55.5 | 172 | 33.7 | 50.6 |
| D13 | 3,260 | 2 | 109 | 33.0 | 45.9 | 147 | 13.6 | 67.3 | 164 | 14.0 | 73.8 |
| F81 | 3,245 | 2 | 46 | 32.6 | 26.1 | 54 | 9.3 | 63.0 | 69 | 5.8 | 62.3 |
| R19 | 3,234 | 2 | 50 | 32.0 | 52.0 | 83 | 30.1 | 57.8 | 82 | 28.0 | 58.5 |
| N70 | 3,211 | 2 | 93 | 43.0 | 48.4 | 128 | 30.5 | 51.6 | 130 | 23.9 | 61.5 |
| I95 | 3,179 | 2 | 87 | 35.6 | 56.3 | 129 | 5.4 | 69.8 | 108 | 4.6 | 81.5 |
| S86 | 3,177 | 2 | 33 | 39.4 | 45.5 | 62 | 29.0 | 58.1 | 63 | 25.4 | 65.1 |
| D37 | 3,176 | 2 | 159 | 33.3 | 44.6 | 220 | 25.4 | 64.6 | 265 | 26.8 | 65.3 |
| L42 | 3,158 | 2 | 40 | 15.0 | 80.0 | 38 | 10.5 | 81.6 | 39 | 2.6 | 87.2 |
| T23 | 3,143 | 3 | 21 | 38.1 | 47.6 | 42 | 26.2 | 54.8 | 36 | 22.2 | 58.3 |
| M18 | 3,131 | 2 | 45 | 28.9 | 55.6 | 54 | 31.5 | 42.6 | 53 | 24.5 | 54.7 |
| I89 | 3,126 | 2 | 90 | 25.6 | 56.7 | 95 | 5.3 | 69.5 | 98 | 14.3 | 61.2 |
| O21 | 3,120 | 2 | 83 | 36.1 | 53.0 | 79 | 15.2 | 60.8 | 171 | 8.2 | 39.2 |
| O03 | 3,112 | 2 | 77 | 37.7 | 53.2 | 85 | 20.0 | 60.0 | 127 | 17.3 | 59.1 |
| L90 | 3,063 | 2 | 89 | 21.4 | 47.2 | 80 | 18.8 | 65.0 | 85 | 16.5 | 64.7 |
| K82 | 3,005 | 2 | 73 | 15.1 | 76.7 | 105 | 0.0 | 100.0 | 119 | 17.6 | 56.3 |
| I82 | 3,004 | 2 | 160 | 46.9 | 46.9 | 174 | 17.8 | 67.2 | 193 | 13.5 | 64.8 |
| M95 | 2,984 | 3 | 71 | 31.0 | 66.2 | 62 | 32.3 | 62.9 | 78 | 25.6 | 68.0 |
| S53 | 2,973 | 4 | 28 | 39.3 | 53.6 | 36 | 27.8 | 66.7 | 40 | 27.5 | 60.0 |
| R20 | 2,973 | 2 | 51 | 35.3 | 52.9 | 75 | 22.7 | 58.7 | 70 | 10.0 | 80.0 |
| K83 | 2,959 | 2 | 122 | 35.2 | 47.5 | 167 | 15.6 | 64.7 | 202 | 16.3 | 67.3 |
| T17 | 2,956 | 5 | 33 | 54.5 | 39.4 | 40 | 42.5 | 50.0 | 33 | 33.3 | 54.5 |
| O08 | 2,955 | 4 | 117 | 45.3 | 43.6 | 88 | 34.1 | 53.4 | 108 | 34.3 | 46.3 |
| G24 | 2,943 | 2 | 36 | 13.9 | 69.4 | 60 | 11.7 | 75.0 | 62 | 9.7 | 83.9 |
| J38 | 2,898 | 2 | 108 | 30.6 | 50.0 | 111 | 23.4 | 69.4 | 116 | 18.1 | 51.7 |
| S46 | 2,898 | 2 | 48 | 29.2 | 50.0 | 63 | 19.0 | 65.1 | 75 | 24.0 | 69.3 |
| R25 | 2,850 | 2 | 39 | 25.6 | 56.4 | 64 | 21.9 | 64.1 | 58 | 24.1 | 69.0 |
| I45 | 2,806 | 2 | 74 | 27.0 | 64.9 | 110 | 8.2 | 76.4 | 104 | 8.7 | 70.2 |
| J37 | 2,797 | 2 | 82 | 37.8 | 52.4 | 89 | 23.6 | 56.2 | 112 | 23.2 | 59.8 |
| E10 | 2,787 | 2 | 104 | 24.0 | 53.9 | 142 | 19.7 | 57.8 | 134 | 24.6 | 58.2 |
| F95 | 2,785 | 2 | 31 | 12.9 | 64.5 | 46 | 15.2 | 60.9 | 42 | 14.3 | 59.5 |
| L28 | 2,780 | 2 | 51 | 56.9 | 37.2 | 48 | 31.2 | 54.2 | 46 | 45.6 | 45.6 |
| F07 | 2,774 | 2 | 73 | 37.0 | 60.3 | 97 | 42.3 | 47.4 | 84 | 36.9 | 56.0 |
| N23 | 2,766 | 2 | 66 | 31.8 | 40.9 | 103 | 25.2 | 53.4 | 107 | 21.5 | 59.8 |
| I88 | 2,750 | 2 | 61 | 36.1 | 52.5 | 92 | 16.3 | 65.2 | 97 | 18.6 | 59.8 |
| O72 | 2,729 | 3 | 117 | 35.9 | 28.2 | 140 | 19.3 | 47.1 | 118 | 16.1 | 50.9 |
| F84 | 2,713 | 2 | 69 | 30.4 | 39.1 | 81 | 22.2 | 54.3 | 93 | 21.5 | 71.0 |
| I78 | 2,705 | 4 | 36 | 22.2 | 58.3 | 36 | 16.7 | 61.1 | 34 | 14.7 | 61.8 |
| D62 | 2,700 | 5 | 135 | 42.2 | 41.5 | 217 | 24.4 | 56.2 | 205 | 27.3 | 53.7 |
| C61 | 2,700 | 2 | 98 | 32.6 | 45.9 | 115 | 26.1 | 58.3 | 167 | 28.1 | 65.9 |
| G20 | 2,681 | 2 | 70 | 31.4 | 64.3 | 95 | 13.7 | 77.9 | 96 | 17.7 | 66.7 |
| H20 | 2,671 | 2 | 86 | 43.0 | 48.8 | 89 | 28.1 | 52.8 | 82 | 30.5 | 48.8 |
| J16 | 2,667 | 3 | 108 | 59.3 | 31.5 | 146 | 21.2 | 65.1 | 121 | 23.1 | 57.9 |
| R69 | 2,635 | 2 | 31 | 29.0 | 58.1 | 45 | 2.2 | 75.6 | 51 | 2.0 | 90.2 |
| D39 | 2,601 | 3 | 110 | 30.0 | 50.0 | 173 | 22.5 | 62.4 | 202 | 14.8 | 75.7 |
| Q38 | 2,592 | 2 | 38 | 42.1 | 50.0 | 38 | 26.3 | 68.4 | 46 | 37.0 | 58.7 |
| H57 | 2,557 | 2 | 48 | 47.9 | 52.1 | 51 | 25.5 | 64.7 | 38 | 52.6 | 42.1 |
| K63 | 2,553 | 2 | 116 | 34.5 | 48.3 | 152 | 22.4 | 58.5 | 155 | 23.2 | 58.7 |
| S66 | 2,529 | 2 | 39 | 35.9 | 59.0 | 63 | 25.4 | 58.7 | 61 | 21.3 | 59.0 |
| H69 | 2,512 | 2 | 52 | 36.5 | 48.1 | 60 | 10.0 | 66.7 | 54 | 5.6 | 64.8 |
| K91 | 2,504 | 2 | 101 | 66.3 | 27.7 | 125 | 13.6 | 79.2 | 108 | 24.1 | 65.7 |
| D38 | 2,501 | 2 | 139 | 32.4 | 54.0 | 190 | 29.5 | 54.2 | 241 | 36.1 | 52.7 |
| N32 | 2,491 | 2 | 80 | 33.8 | 51.2 | 102 | 17.6 | 54.9 | 104 | 21.1 | 60.6 |
| E89 | 2,486 | 2 | 101 | 45.5 | 46.5 | 90 | 14.4 | 67.8 | 79 | 17.7 | 58.2 |
| T78 | 2,466 | 2 | 69 | 37.7 | 53.6 | 73 | 31.5 | 58.9 | 73 | 27.4 | 63.0 |
| Q82 | 2,465 | 2 | 33 | 27.3 | 45.5 | 36 | 30.6 | 52.8 | 38 | 26.3 | 57.9 |
| K14 | 2,464 | 2 | 39 | 38.5 | 59.0 | 51 | 21.6 | 62.8 | 54 | 16.7 | 68.5 |
| R18 | 2,460 | 2 | 161 | 37.9 | 52.2 | 219 | 25.1 | 62.6 | 197 | 24.4 | 65.5 |
| I86 | 2,459 | 5 | 41 | 19.5 | 46.3 | 54 | 18.5 | 51.9 | 61 | 9.8 | 49.2 |
| N71 | 2,441 | 2 | 102 | 47.1 | 46.1 | 112 | 8.3 | 91.7 | 104 | 26.0 | 53.9 |
| I73 | 2,395 | 2 | 52 | 30.8 | 61.5 | 73 | 19.2 | 71.2 | 72 | 16.7 | 63.9 |
| Q66 | 2,389 | 4 | 21 | 23.8 | 61.9 | 26 | 23.1 | 61.5 | 23 | 30.4 | 60.9 |
| Z95 | 2,340 | 2 | 164 | 36.6 | 47.6 | 174 | 17.8 | 69.5 | 183 | 20.3 | 71.4 |
| A38 | 2,315 | 2 | 36 | 33.3 | 44.4 | 43 | 27.9 | 58.1 | 43 | 30.2 | 51.2 |
| N13 | 2,299 | 2 | 115 | 46.1 | 40.0 | 164 | 22.6 | 61.0 | 155 | 21.3 | 61.3 |
| O64 | 2,297 | 2 | 103 | 13.6 | 58.2 | 112 | 22.3 | 36.6 | 105 | 17.1 | 61.0 |
| T84 | 2,284 | 3 | 141 | 36.2 | 39.7 | 151 | 33.1 | 59.6 | 150 | 30.7 | 58.0 |
| G51 | 2,279 | 2 | 62 | 29.0 | 53.2 | 90 | 15.6 | 57.8 | 87 | 19.5 | 69.0 |
| H33 | 2,243 | 2 | 70 | 31.4 | 52.9 | 83 | 24.1 | 62.6 | 73 | 12.3 | 68.5 |
| J36 | 2,241 | 2 | 76 | 38.2 | 42.1 | 102 | 14.7 | 57.8 | 98 | 13.3 | 65.3 |
| M46 | 2,224 | 2 | 111 | 37.8 | 47.8 | 118 | 18.6 | 53.4 | 125 | 16.8 | 69.6 |
| F70 | 2,218 | 2 | 28 | 35.7 | 57.1 | 36 | 19.4 | 75.0 | 33 | 18.2 | 81.8 |
| S10 | 2,203 | 2 | 40 | 27.5 | 65.0 | 36 | 13.9 | 83.3 | 29 | 6.9 | 89.7 |
| S13 | 2,202 | 2 | 29 | 31.0 | 69.0 | 36 | 17.1 | 57.1 | 43 | 16.3 | 58.1 |
| M05 | 2,189 | 3 | 86 | 34.9 | 46.5 | 94 | 23.4 | 60.6 | 96 | 31.2 | 56.2 |
| H47 | 2,179 | 2 | 78 | 28.2 | 70.5 | 81 | 35.8 | 49.4 | 82 | 25.6 | 51.2 |
| G31 | 2,177 | 2 | 85 | 23.5 | 55.3 | 130 | 12.3 | 73.1 | 126 | 17.5 | 77.8 |
| G46 | 2,173 | 5 | 85 | 32.9 | 52.9 | 152 | 34.2 | 52.0 | 157 | 29.9 | 54.1 |
| I12 | 2,137 | 2 | 69 | 43.5 | 40.6 | 110 | 12.7 | 67.3 | 97 | 17.5 | 65.0 |
| H44 | 2,118 | 2 | 48 | 45.8 | 39.6 | 55 | 43.6 | 49.1 | 51 | 47.1 | 49.0 |
| N43 | 2,118 | 2 | 53 | 20.8 | 50.9 | 60 | 10.0 | 55.0 | 73 | 6.8 | 65.8 |
| L92 | 2,115 | 4 | 46 | 21.7 | 60.9 | 47 | 23.4 | 55.3 | 44 | 27.3 | 59.1 |
| N19 | 2,102 | 2 | 106 | 31.1 | 50.9 | 179 | 6.7 | 71.5 | 151 | 18.5 | 65.6 |
| L43 | 2,086 | 2 | 43 | 32.6 | 51.2 | 43 | 30.2 | 51.2 | 40 | 45.0 | 40.0 |
| C50 | 2,029 | 2 | 74 | 35.1 | 50.0 | 115 | 45.2 | 43.5 | 145 | 41.4 | 38.6 |
| O13 | 2,026 | 2 | 90 | 17.8 | 62.2 | 128 | 8.6 | 53.1 | 139 | 11.5 | 44.6 |
| O65 | 2,026 | 4 | 100 | 11.1 | 40.4 | 96 | 9.4 | 62.5 | 103 | 21.4 | 58.2 |
| B83 | 2,022 | 2 | 22 | 22.7 | 59.1 | 20 | 30.0 | 60.0 | 16 | 31.2 | 56.2 |
| F34 | 2,021 | 2 | 43 | 39.5 | 55.8 | 51 | 35.3 | 56.9 | 52 | 38.5 | 50.0 |
| F17 | 2,005 | 2 | 57 | 19.3 | 50.9 | 63 | 19.0 | 77.8 | 61 | 18.0 | 70.5 |
| C34 | 1,985 | 2 | 127 | 30.7 | 55.1 | 192 | 30.7 | 58.3 | 214 | 31.8 | 64.5 |
| L65 | 1,983 | 2 | 31 | 32.3 | 58.1 | 38 | 23.7 | 63.2 | 33 | 24.2 | 60.6 |
| M84 | 1,981 | 2 | 87 | 34.5 | 47.1 | 104 | 24.0 | 67.3 | 104 | 26.0 | 67.3 |
| R56 | 1,975 | 4 | 46 | 26.1 | 73.9 | 81 | 11.1 | 74.1 | 84 | 22.6 | 56.0 |
| S71 | 1,960 | 2 | 33 | 69.7 | 27.3 | 42 | 23.8 | 71.4 | 34 | 29.4 | 61.8 |
| O41 | 1,952 | 2 | 99 | 29.3 | 47.5 | 126 | 12.7 | 36.5 | 124 | 28.2 | 57.3 |
| L05 | 1,939 | 2 | 36 | 36.1 | 58.3 | 56 | 35.7 | 50.0 | 67 | 13.4 | 71.6 |
| I74 | 1,928 | 2 | 118 | 40.7 | 47.5 | 173 | 25.4 | 59.0 | 164 | 23.8 | 57.3 |
| H15 | 1,918 | 3 | 29 | 44.8 | 48.3 | 42 | 28.6 | 57.1 | 37 | 35.1 | 56.8 |
| A74 | 1,893 | 2 | 38 | 36.8 | 39.5 | 44 | 18.2 | 59.1 | 35 | 22.9 | 51.4 |
| D41 | 1,892 | 2 | 91 | 23.1 | 56.0 | 148 | 30.4 | 53.4 | 189 | 28.0 | 57.7 |
| D10 | 1,886 | 2 | 61 | 39.3 | 45.9 | 55 | 23.6 | 63.6 | 68 | 23.5 | 61.8 |
| H93 | 1,879 | 2 | 37 | 35.1 | 62.2 | 49 | 26.5 | 59.2 | 46 | 37.0 | 54.4 |
| E56 | 1,845 | 2 | 26 | 15.4 | 69.2 | 57 | 14.0 | 79.0 | 40 | 22.5 | 67.5 |
| O60 | 1,842 | 2 | 98 | 43.9 | 41.8 | 126 | 6.3 | 73.8 | 121 | 12.4 | 59.5 |
| R64 | 1,836 | 2 | 158 | 43.7 | 49.4 | 159 | 4.4 | 88.7 | 110 | 6.4 | 88.2 |
| L63 | 1,835 | 2 | 29 | 27.6 | 51.7 | 41 | 24.4 | 46.3 | 41 | 26.8 | 48.8 |
| I71 | 1,827 | 2 | 91 | 44.0 | 39.6 | 157 | 10.2 | 73.2 | 169 | 22.5 | 63.9 |
| D24 | 1,825 | 2 | 46 | 43.5 | 34.8 | 41 | 34.1 | 46.3 | 52 | 30.8 | 48.1 |
| N73 | 1,806 | 2 | 118 | 33.0 | 46.6 | 129 | 18.6 | 57.4 | 125 | 21.6 | 60.0 |
| K65 | 1,789 | 2 | 155 | 38.1 | 40.6 | 184 | 18.5 | 56.5 | 197 | 22.3 | 62.9 |
| O92 | 1,778 | 4 | 112 | 20.5 | 58.0 | 48 | 37.5 | 50.0 | 32 | 15.6 | 78.1 |
| N62 | 1,776 | 2 | 31 | 32.3 | 58.1 | 36 | 5.6 | 72.2 | 41 | 4.9 | 82.9 |
| J43 | 1,775 | 2 | 98 | 22.4 | 57.1 | 138 | 15.9 | 67.4 | 142 | 15.5 | 64.8 |
| O69 | 1,774 | 2 | 90 | 21.1 | 45.6 | 107 | 11.2 | 42.1 | 93 | 10.8 | 44.1 |
| G83 | 1,761 | 2 | 94 | 43.6 | 38.3 | 105 | 31.4 | 62.9 | 97 | 21.6 | 69.1 |
| I15 | 1,758 | 2 | 124 | 30.6 | 51.6 | 124 | 33.9 | 50.8 | 147 | 27.9 | 53.7 |
| G82 | 1,741 | 2 | 111 | 35.1 | 46.9 | 138 | 42.0 | 47.8 | 143 | 24.5 | 62.9 |
| T24 | 1,736 | 2 | 22 | 50.0 | 40.9 | 51 | 17.6 | 72.6 | 43 | 20.9 | 65.1 |
| O75 | 1,719 | 5 | 81 | 34.6 | 58.0 | 84 | 29.8 | 50.0 | 68 | 35.3 | 47.1 |
| H72 | 1,697 | 3 | 55 | 30.9 | 50.9 | 51 | 17.6 | 62.8 | 51 | 29.4 | 54.9 |
| F60 | 1,697 | 2 | 59 | 61.0 | 33.9 | 66 | 12.1 | 78.8 | 67 | 10.4 | 70.2 |
| C18 | 1,681 | 2 | 112 | 27.7 | 50.9 | 158 | 30.4 | 60.8 | 199 | 32.2 | 61.8 |
| R43 | 1,680 | 2 | 43 | 39.5 | 44.2 | 53 | 17.0 | 67.9 | 63 | 38.1 | 39.7 |
| F50 | 1,680 | 2 | 54 | 35.2 | 55.6 | 69 | 26.1 | 59.4 | 87 | 28.7 | 55.2 |
| S76 | 1,669 | 4 | 28 | 39.3 | 60.7 | 37 | 24.3 | 67.6 | 33 | 24.2 | 69.7 |
| T79 | 1,662 | 2 | 75 | 57.3 | 36.0 | 135 | 47.4 | 34.8 | 118 | 39.0 | 48.3 |
| N77 | 1,661 | 2 | 34 | 47.1 | 50.0 | 45 | 51.1 | 46.7 | 37 | 56.8 | 40.5 |
| K51 | 1,650 | 2 | 96 | 25.0 | 58.3 | 94 | 19.1 | 57.5 | 102 | 17.6 | 62.8 |
| H34 | 1,632 | 3 | 68 | 39.7 | 32.4 | 77 | 18.2 | 67.5 | 84 | 28.6 | 51.2 |
| J21 | 1,606 | 2 | 55 | 25.4 | 54.5 | 75 | 10.7 | 64.0 | 58 | 1.7 | 75.9 |
| R61 | 1,602 | 2 | 16 | 6.2 | 93.8 | 36 | 8.3 | 61.1 | 27 | 11.1 | 66.7 |
| E86 | 1,596 | 2 | 63 | 30.2 | 50.8 | 115 | 13.9 | 60.0 | 90 | 11.1 | 63.3 |
| D70 | 1,574 | 2 | 184 | 17.9 | 65.5 | 159 | 23.3 | 66.7 | 192 | 25.0 | 66.7 |
| K74 | 1,569 | 2 | 108 | 37.0 | 49.1 | 146 | 24.0 | 71.9 | 169 | 32.0 | 61.5 |
| B30 | 1,564 | 2 | 21 | 19.0 | 81.0 | 27 | 18.5 | 77.8 | 18 | 11.1 | 83.3 |
| M96 | 1,563 | 2 | 71 | 52.1 | 42.2 | 68 | 8.8 | 83.8 | 51 | 5.9 | 92.2 |
| M93 | 1,563 | 2 | 39 | 38.5 | 53.9 | 31 | 29.0 | 67.7 | 39 | 23.1 | 69.2 |
| D26 | 1,549 | 2 | 25 | 20.0 | 60.0 | 57 | 17.5 | 56.1 | 63 | 19.0 | 71.4 |
| O91 | 1,533 | 3 | 117 | 15.4 | 55.6 | 49 | 42.9 | 53.1 | 44 | 45.5 | 47.7 |
| D30 | 1,523 | 3 | 74 | 14.9 | 43.2 | 96 | 24.0 | 61.5 | 105 | 13.3 | 58.1 |
| K73 | 1,512 | 2 | 60 | 28.3 | 46.7 | 84 | 26.2 | 58.3 | 89 | 23.6 | 68.5 |
| F05 | 1,496 | 2 | 107 | 54.2 | 40.2 | 183 | 23.0 | 61.8 | 191 | 46.6 | 49.2 |
| J90 | 1,496 | 2 | 117 | 40.2 | 44.4 | 190 | 16.8 | 67.4 | 178 | 19.7 | 55.6 |
| F39 | 1,496 | 2 | 54 | 25.9 | 61.1 | 51 | 27.4 | 62.8 | 60 | 33.3 | 58.3 |
| I36 | 1,485 | 2 | 94 | 20.2 | 60.6 | 158 | 20.2 | 64.6 | 148 | 15.5 | 61.5 |
| T26 | 1,483 | 2 | 22 | 59.1 | 31.8 | 28 | 46.4 | 53.6 | 24 | 25.0 | 70.8 |
| I51 | 1,468 | 2 | 68 | 26.5 | 61.8 | 107 | 18.7 | 62.6 | 115 | 16.5 | 69.6 |
| E22 | 1,455 | 2 | 61 | 18.0 | 23.0 | 61 | 16.4 | 55.7 | 95 | 10.5 | 67.4 |
| I61 | 1,452 | 3 | 78 | 21.8 | 57.7 | 149 | 36.2 | 51.7 | 147 | 22.4 | 62.6 |
| E21 | 1,444 | 2 | 62 | 14.5 | 50.0 | 84 | 21.4 | 50.0 | 71 | 25.4 | 46.5 |
| I43 | 1,438 | 2 | 54 | 42.6 | 51.9 | 51 | 19.6 | 72.6 | 48 | 16.7 | 68.8 |
| K06 | 1,432 | 5 | 11 | 81.8 | 18.2 | 13 | 23.1 | 76.9 | 8 | 37.5 | 62.5 |
| B82 | 1,394 | 5 | 16 | 31.2 | 56.2 | 11 | 36.4 | 63.6 | 12 | 33.3 | 58.3 |
| B90 | 1,384 | 2 | 62 | 24.2 | 45.2 | 75 | 26.7 | 50.7 | 75 | 14.7 | 70.7 |
| N61 | 1,379 | 2 | 83 | 30.1 | 62.6 | 51 | 35.3 | 54.9 | 44 | 11.4 | 79.6 |
| B95 | 1,357 | 2 | 65 | 35.4 | 50.8 | 67 | 46.3 | 41.8 | 46 | 39.1 | 47.8 |
| T18 | 1,354 | 2 | 33 | 36.4 | 57.6 | 40 | 27.5 | 67.5 | 33 | 27.3 | 60.6 |
| I46 | 1,350 | 3 | 127 | 27.6 | 54.3 | 180 | 22.2 | 62.2 | 152 | 19.1 | 63.8 |
| K75 | 1,347 | 2 | 83 | 27.7 | 59.0 | 112 | 22.3 | 61.6 | 112 | 17.9 | 75.0 |
| M07 | 1,343 | 2 | 54 | 27.8 | 51.9 | 52 | 34.6 | 61.5 | 58 | 25.9 | 56.9 |
| A66 | 1,337 | 2 | 12 | 33.3 | 50.0 | 17 | 17.6 | 41.2 | 16 | 12.5 | 56.2 |
| S09 | 1,333 | 2 | 31 | 51.6 | 35.5 | 40 | 17.5 | 75.0 | 36 | 16.7 | 77.8 |
| J81 | 1,332 | 2 | 98 | 32.6 | 44.9 | 176 | 15.9 | 60.2 | 149 | 20.8 | 62.4 |
| T22 | 1,329 | 2 | 20 | 25.0 | 65.0 | 35 | 17.1 | 77.1 | 30 | 16.7 | 73.3 |
| F93 | 1,328 | 4 | 31 | 41.9 | 41.9 | 37 | 27.0 | 56.8 | 41 | 29.3 | 53.7 |
| I22 | 1,327 | 2 | 95 | 33.7 | 49.5 | 140 | 30.7 | 50.0 | 156 | 27.6 | 61.5 |
| E80 | 1,316 | 5 | 47 | 21.3 | 57.5 | 65 | 21.5 | 63.1 | 68 | 14.7 | 69.1 |
| D75 | 1,314 | 2 | 60 | 33.3 | 45.0 | 85 | 24.7 | 71.8 | 102 | 34.3 | 54.9 |
| D14 | 1,291 | 3 | 93 | 35.5 | 34.4 | 95 | 30.5 | 61.0 | 111 | 28.8 | 50.5 |
| O87 | 1,291 | 5 | 87 | 28.7 | 58.6 | 70 | 10.0 | 58.6 | 65 | 7.7 | 60.0 |
| T25 | 1,289 | 2 | 17 | 52.9 | 35.3 | 37 | 27.0 | 70.3 | 30 | 33.3 | 63.3 |
| O12 | 1,281 | 2 | 68 | 10.3 | 52.9 | 102 | 9.8 | 64.7 | 120 | 8.3 | 66.7 |
| K66 | 1,275 | 5 | 97 | 33.0 | 56.7 | 132 | 22.0 | 61.4 | 107 | 17.8 | 71.0 |
| Z20 | 1,273 | 2 | 11 | 36.4 | 63.6 | 13 | 30.8 | 61.5 | 13 | 30.8 | 61.5 |
| B09 | 1,273 | 3 | 19 | 10.5 | 84.2 | 31 | 16.1 | 80.7 | 17 | 11.8 | 70.6 |
| J69 | 1,260 | 2 | 112 | 49.1 | 39.3 | 167 | 26.5 | 54.2 | 155 | 16.1 | 71.6 |
| K27 | 1,250 | 2 | 74 | 23.0 | 55.4 | 85 | 27.1 | 57.6 | 89 | 30.3 | 61.8 |
| O48 | 1,246 | 2 | 61 | 21.3 | 70.5 | 88 | 44.3 | 43.2 | 90 | 51.1 | 41.1 |
| R93 | 1,245 | 2 | 39 | 23.1 | 64.1 | 41 | 17.1 | 65.8 | 27 | 22.2 | 70.4 |
| S33 | 1,240 | 2 | 25 | 48.0 | 52.0 | 38 | 15.8 | 73.7 | 36 | 27.8 | 63.9 |
| H27 | 1,239 | 2 | 53 | 28.3 | 60.4 | 58 | 13.8 | 82.8 | 69 | 10.1 | 85.5 |
| F91 | 1,236 | 2 | 28 | 42.9 | 42.9 | 37 | 29.7 | 54.0 | 41 | 26.8 | 36.6 |
| S27 | 1,231 | 2 | 61 | 23.0 | 62.3 | 123 | 16.3 | 68.3 | 127 | 22.8 | 62.2 |
| F19 | 1,228 | 2 | 48 | 41.7 | 50.0 | 68 | 29.4 | 51.5 | 79 | 24.0 | 62.0 |
| D16 | 1,223 | 2 | 31 | 38.7 | 51.6 | 34 | 29.4 | 64.7 | 41 | 22.0 | 68.3 |
| A07 | 1,218 | 2 | 58 | 22.4 | 53.5 | 53 | 20.8 | 58.5 | 47 | 25.5 | 55.3 |
| F23 | 1,205 | 2 | 71 | 39.4 | 46.5 | 78 | 37.2 | 51.3 | 99 | 33.3 | 57.6 |
| D35 | 1,199 | 2 | 69 | 24.6 | 55.1 | 89 | 24.7 | 50.6 | 67 | 37.3 | 55.2 |
| D68 | 1,197 | 2 | 92 | 21.7 | 60.9 | 99 | 19.2 | 61.6 | 89 | 31.5 | 58.4 |
| M86 | 1,196 | 2 | 84 | 41.7 | 50.0 | 95 | 21.0 | 63.2 | 98 | 27.6 | 68.4 |
| R15 | 1,195 | 2 | 22 | 22.7 | 68.2 | 38 | 10.5 | 79.0 | 29 | 20.7 | 48.3 |
| H74 | 1,193 | 4 | 59 | 39.0 | 54.2 | 37 | 24.3 | 70.3 | 35 | 17.1 | 62.9 |
| R16 | 1,185 | 2 | 62 | 30.6 | 59.7 | 93 | 16.1 | 63.4 | 98 | 18.4 | 58.2 |
| O22 | 1,175 | 2 | 60 | 16.7 | 70.0 | 79 | 11.4 | 62.0 | 101 | 9.9 | 72.3 |
| L80 | 1,171 | 2 | 24 | 33.3 | 37.5 | 32 | 28.1 | 43.8 | 33 | 30.3 | 45.5 |
| F31 | 1,154 | 2 | 49 | 36.7 | 59.2 | 43 | 34.9 | 58.1 | 50 | 38.0 | 48.0 |
| T21 | 1,133 | 2 | 28 | 21.4 | 67.9 | 50 | 20.0 | 74.0 | 34 | 17.6 | 76.5 |
| N21 | 1,129 | 2 | 64 | 23.4 | 46.9 | 88 | 18.2 | 59.1 | 93 | 24.7 | 58.1 |
| L53 | 1,127 | 2 | 45 | 44.4 | 40.0 | 50 | 30.0 | 56.0 | 42 | 42.9 | 47.6 |
| Q21 | 1,123 | 2 | 50 | 30.0 | 60.0 | 64 | 6.2 | 71.9 | 71 | 12.7 | 42.2 |
| L95 | 1,123 | 2 | 59 | 18.6 | 71.2 | 71 | 19.7 | 69.0 | 61 | 9.8 | 75.4 |
| N51 | 1,117 | 5 | 30 | 33.3 | 60.0 | 38 | 47.4 | 44.7 | 35 | 20.0 | 74.3 |
| D04 | 1,116 | 2 | 30 | 43.3 | 53.3 | 39 | 30.8 | 59.0 | 37 | 51.4 | 37.8 |
| Z90 | 1,108 | 3 | 62 | 33.9 | 59.7 | 84 | 14.3 | 67.9 | 64 | 17.2 | 64.1 |
| S03 | 1,102 | 5 | 18 | 38.9 | 61.1 | 22 | 27.3 | 72.7 | 22 | 27.3 | 63.6 |
| E29 | 1,094 | 2 | 37 | 35.1 | 62.2 | 36 | 25.0 | 66.7 | 36 | 33.3 | 50.0 |
| M12 | 1,088 | 3 | 26 | 46.1 | 46.1 | 28 | 39.3 | 50.0 | 37 | 40.5 | 59.5 |
| O00 | 1,086 | 2 | 45 | 33.3 | 46.7 | 84 | 23.8 | 65.5 | 83 | 24.1 | 53.0 |
| D72 | 1,084 | 2 | 48 | 10.4 | 72.9 | 65 | 12.3 | 76.9 | 69 | 26.1 | 65.2 |
| F42 | 1,081 | 2 | 31 | 41.9 | 54.8 | 32 | 25.0 | 75.0 | 39 | 20.5 | 79.5 |
| E83 | 1,077 | 2 | 58 | 19.0 | 62.1 | 95 | 9.5 | 64.2 | 82 | 12.2 | 52.4 |
| L91 | 1,060 | 2 | 18 | 38.9 | 50.0 | 13 | 38.5 | 61.5 | 16 | 43.8 | 50.0 |
| D47 | 1,059 | 2 | 79 | 30.4 | 45.6 | 110 | 25.4 | 59.1 | 141 | 24.8 | 62.4 |
| S68 | 1,059 | 2 | 36 | 33.3 | 58.3 | 56 | 41.1 | 53.6 | 53 | 24.5 | 73.6 |
| T42 | 1,043 | 2 | 64 | 20.3 | 65.6 | 97 | 11.3 | 66.0 | 92 | 10.9 | 75.0 |
| F20 | 1,041 | 2 | 45 | 28.9 | 40.0 | 46 | 17.4 | 63.0 | 54 | 31.5 | 50.0 |
| B94 | 1,041 | 2 | 61 | 41.0 | 50.8 | 49 | 18.4 | 73.5 | 41 | 29.3 | 70.7 |
| C43 | 1,040 | 2 | 48 | 47.9 | 47.9 | 69 | 39.1 | 47.8 | 89 | 27.0 | 67.4 |
| B81 | 1,040 | 4 | 22 | 27.3 | 45.5 | 24 | 25.0 | 62.5 | 14 | 28.6 | 64.3 |
| C64 | 1,033 | 2 | 82 | 25.6 | 43.9 | 113 | 25.7 | 62.0 | 131 | 28.2 | 69.5 |
| O98 | 1,029 | 2 | 63 | 25.4 | 54.0 | 87 | 24.1 | 52.9 | 106 | 16.0 | 73.6 |
| O40 | 1,009 | 2 | 66 | 30.3 | 50.0 | 98 | 6.1 | 50.0 | 95 | 22.1 | 43.2 |
| F13 | 1,007 | 2 | 36 | 25.0 | 61.1 | 61 | 27.9 | 50.8 | 45 | 31.1 | 60.0 |
| R40 | 1,007 | 2 | 83 | 51.8 | 34.9 | 111 | 18.9 | 66.7 | 96 | 15.6 | 80.2 |
| S69 | 1,001 | 5 | 29 | 44.8 | 51.7 | 39 | 33.3 | 66.7 | 37 | 43.2 | 51.4 |
| K90 | 999 | 2 | 66 | 34.9 | 51.5 | 74 | 14.9 | 71.6 | 57 | 19.3 | 71.9 |
| K72 | 997 | 4 | 111 | 43.2 | 51.4 | 116 | 26.7 | 60.3 | 97 | 28.9 | 62.9 |
| N75 | 979 | 2 | 37 | 35.1 | 51.4 | 51 | 29.4 | 54.9 | 43 | 30.2 | 58.1 |
| D53 | 977 | 2 | 70 | 17.1 | 70.0 | 95 | 8.5 | 63.8 | 89 | 14.6 | 67.4 |
| K55 | 974 | 2 | 91 | 27.5 | 61.5 | 139 | 23.7 | 53.2 | 122 | 16.4 | 64.8 |
| R57 | 967 | 2 | 108 | 50.9 | 36.1 | 138 | 33.3 | 44.9 | 119 | 31.9 | 43.7 |
| S31 | 953 | 2 | 35 | 20.0 | 71.4 | 47 | 38.3 | 57.5 | 41 | 22.0 | 70.7 |
| B85 | 944 | 5 | 25 | 24.0 | 28.0 | 33 | 12.1 | 51.5 | 19 | 21.0 | 10.5 |
| H80 | 935 | 2 | 24 | 29.2 | 54.2 | 28 | 28.6 | 46.4 | 28 | 25.0 | 64.3 |
| J17 | 932 | 2 | 69 | 40.6 | 49.3 | 101 | 18.8 | 67.3 | 86 | 20.9 | 74.4 |
| T43 | 931 | 2 | 67 | 61.2 | 31.3 | 90 | 13.3 | 63.3 | 94 | 22.3 | 63.8 |
| Q67 | 930 | 3 | 24 | 37.5 | 62.5 | 24 | 45.8 | 54.2 | 28 | 39.3 | 50.0 |
| N49 | 925 | 3 | 40 | 25.0 | 47.5 | 43 | 20.9 | 65.1 | 44 | 20.4 | 70.4 |
| K50 | 923 | 2 | 82 | 34.1 | 58.5 | 87 | 18.4 | 66.7 | 88 | 22.7 | 61.4 |
| S96 | 922 | 4 | 19 | 36.8 | 63.2 | 24 | 37.5 | 62.5 | 22 | 45.5 | 54.5 |
| J13 | 921 | 2 | 64 | 25.0 | 57.8 | 127 | 20.5 | 67.7 | 102 | 24.5 | 52.9 |
| L00 | 917 | 5 | 14 | 42.9 | 35.7 | 23 | 8.7 | 69.6 | 18 | 16.7 | 72.2 |
| R82 | 914 | 2 | 51 | 29.4 | 49.0 | 61 | 27.9 | 42.6 | 45 | 37.8 | 51.1 |
| C67 | 912 | 3 | 79 | 25.3 | 63.3 | 99 | 27.3 | 55.6 | 135 | 26.7 | 61.5 |
| S41 | 911 | 2 | 23 | 39.1 | 56.5 | 29 | 44.8 | 51.7 | 25 | 40.0 | 56.0 |
| D31 | 908 | 5 | 38 | 36.8 | 55.3 | 44 | 25.0 | 70.4 | 28 | 35.7 | 50.0 |
| N99 | 906 | 2 | 66 | 59.1 | 25.8 | 62 | 29.0 | 58.1 | 30 | 40.0 | 56.7 |
| G53 | 903 | 4 | 41 | 39.0 | 56.1 | 38 | 31.6 | 63.2 | 27 | 14.8 | 85.2 |
| C16 | 903 | 2 | 83 | 34.9 | 50.6 | 123 | 19.5 | 73.2 | 158 | 33.1 | 62.4 |
| R94 | 897 | 3 | 23 | 34.8 | 60.9 | 38 | 29.0 | 60.5 | 30 | 30.0 | 60.0 |
| F02 | 894 | 2 | 70 | 35.7 | 57.1 | 78 | 18.0 | 71.8 | 59 | 15.2 | 69.5 |
| L11 | 884 | 5 | 21 | 19.0 | 71.4 | 21 | 23.8 | 42.9 | 16 | 18.8 | 62.5 |
| J84 | 881 | 2 | 99 | 24.2 | 65.7 | 114 | 17.5 | 60.5 | 135 | 14.1 | 64.4 |
| M08 | 879 | 2 | 41 | 43.9 | 48.8 | 52 | 17.3 | 69.2 | 55 | 43.6 | 41.8 |
| F22 | 864 | 2 | 39 | 35.9 | 48.7 | 52 | 28.9 | 65.4 | 50 | 32.0 | 60.0 |
| O30 | 858 | 2 | 52 | 44.2 | 46.1 | 62 | 16.1 | 72.6 | 114 | 17.5 | 78.1 |
| S99 | 856 | 4 | 11 | 36.4 | 63.6 | 13 | 23.1 | 76.9 | 10 | 30.0 | 70.0 |
| H73 | 855 | 2 | 25 | 44.0 | 56.0 | 43 | 37.2 | 60.5 | 23 | 26.1 | 69.6 |
| K61 | 852 | 2 | 59 | 37.3 | 44.1 | 79 | 16.5 | 73.4 | 82 | 18.3 | 70.7 |
| O86 | 848 | 2 | 107 | 43.0 | 35.5 | 74 | 17.6 | 68.9 | 58 | 24.1 | 56.9 |
| S73 | 845 | 2 | 30 | 60.0 | 36.7 | 40 | 32.5 | 55.0 | 26 | 50.0 | 46.1 |
| J94 | 845 | 3 | 92 | 30.4 | 45.6 | 139 | 11.6 | 66.7 | 127 | 13.4 | 71.7 |
| O14 | 844 | 2 | 75 | 22.7 | 48.0 | 103 | 22.3 | 50.5 | 104 | 26.0 | 53.9 |
| J93 | 837 | 2 | 81 | 27.2 | 49.4 | 121 | 20.8 | 58.3 | 116 | 13.8 | 66.4 |
| D32 | 827 | 3 | 47 | 19.1 | 57.5 | 71 | 18.3 | 62.0 | 79 | 19.0 | 68.3 |
| O90 | 823 | 2 | 108 | 41.7 | 45.4 | 61 | 41.0 | 49.2 | 49 | 24.5 | 67.3 |
| E16 | 823 | 2 | 59 | 8.5 | 71.2 | 81 | 7.4 | 84.0 | 53 | 7.6 | 83.0 |
| F71 | 822 | 2 | 11 | 36.4 | 54.5 | 20 | 25.0 | 75.0 | 13 | 15.4 | 84.6 |
| O10 | 821 | 2 | 61 | 24.6 | 47.5 | 78 | 24.4 | 57.7 | 117 | 8.6 | 53.9 |
| G30 | 820 | 2 | 36 | 25.0 | 55.6 | 52 | 15.4 | 73.1 | 39 | 25.6 | 66.7 |
| C25 | 817 | 2 | 91 | 34.1 | 59.3 | 140 | 27.1 | 67.9 | 177 | 31.1 | 63.8 |
| T40 | 817 | 2 | 47 | 12.8 | 76.6 | 61 | 11.5 | 70.5 | 51 | 11.8 | 72.6 |
| H21 | 816 | 2 | 47 | 38.3 | 48.9 | 51 | 41.2 | 52.9 | 47 | 10.6 | 68.1 |
| O35 | 815 | 3 | 59 | 32.2 | 40.7 | 72 | 37.5 | 58.3 | 120 | 26.7 | 57.5 |
| D44 | 813 | 4 | 60 | 28.3 | 58.3 | 78 | 19.2 | 59.0 | 80 | 25.0 | 66.2 |
| D40 | 809 | 2 | 47 | 31.9 | 53.2 | 59 | 32.2 | 64.4 | 99 | 34.3 | 59.6 |
| N96 | 807 | 2 | 50 | 44.0 | 56.0 | 19 | 47.4 | 47.4 | 69 | 31.9 | 59.4 |
| I99 | 801 | 2 | 21 | 42.9 | 57.1 | 27 | 14.8 | 85.2 | 21 | 28.6 | 71.4 |
| M45 | 800 | 2 | 40 | 40.0 | 45.0 | 40 | 40.0 | 50.0 | 38 | 36.8 | 47.4 |
| O07 | 800 | 2 | 67 | 40.3 | 53.7 | 52 | 30.8 | 61.5 | 56 | 28.6 | 60.7 |
| R17 | 800 | 2 | 58 | 36.2 | 53.5 | 131 | 25.2 | 62.6 | 133 | 22.6 | 70.7 |
| E13 | 797 | 2 | 92 | 19.6 | 63.0 | 86 | 22.1 | 69.8 | 84 | 16.7 | 77.4 |
| G90 | 794 | 2 | 52 | 17.3 | 75.0 | 56 | 7.1 | 78.6 | 54 | 11.1 | 70.4 |
| H83 | 793 | 2 | 34 | 26.5 | 64.7 | 34 | 35.3 | 55.9 | 27 | 37.0 | 48.1 |
| J47 | 790 | 2 | 61 | 34.4 | 49.2 | 72 | 27.8 | 54.2 | 63 | 19.0 | 63.5 |
| C20 | 786 | 2 | 80 | 32.5 | 55.0 | 96 | 32.3 | 60.4 | 130 | 33.9 | 59.2 |
| E74 | 785 | 2 | 21 | 28.6 | 66.7 | 30 | 13.3 | 73.3 | 18 | 27.8 | 72.2 |
| L64 | 781 | 5 | 21 | 19.0 | 81.0 | 23 | 17.4 | 60.9 | 14 | 28.6 | 42.9 |
| C54 | 759 | 2 | 79 | 27.9 | 49.4 | 83 | 32.5 | 57.8 | 138 | 34.8 | 62.3 |
| Q44 | 751 | 2 | 36 | 16.7 | 69.4 | 49 | 16.3 | 67.3 | 41 | 14.6 | 58.5 |
| L56 | 751 | 5 | 28 | 21.4 | 71.4 | 23 | 34.8 | 56.5 | 20 | 30.0 | 60.0 |
| A84 | 750 | 2 | 60 | 25.0 | 68.3 | 73 | 13.7 | 69.9 | 65 | 12.3 | 75.4 |
| S23 | 749 | 2 | 15 | 40.0 | 60.0 | 18 | 33.3 | 61.1 | 13 | 38.5 | 61.5 |
| R27 | 744 | 2 | 53 | 43.4 | 54.7 | 59 | 20.3 | 55.9 | 61 | 4.9 | 85.2 |
| B88 | 740 | 4 | 23 | 26.1 | 65.2 | 21 | 28.6 | 66.7 | 20 | 30.0 | 65.0 |
| R91 | 728 | 2 | 45 | 24.4 | 62.2 | 56 | 28.6 | 46.4 | 59 | 11.9 | 86.4 |
| O04 | 728 | 2 | 61 | 54.1 | 41.0 | 61 | 26.2 | 37.7 | 59 | 15.2 | 49.1 |
| Q51 | 726 | 2 | 28 | 21.4 | 57.1 | 35 | 14.3 | 65.7 | 51 | 13.7 | 78.4 |
| R41 | 723 | 2 | 25 | 24.0 | 76.0 | 38 | 23.7 | 73.7 | 38 | 36.8 | 63.2 |
| A48 | 717 | 2 | 63 | 33.3 | 55.6 | 88 | 35.2 | 51.1 | 62 | 22.6 | 59.7 |
| Q53 | 706 | 2 | 16 | 31.2 | 50.0 | 12 | 25.0 | 58.3 | 19 | 31.6 | 52.6 |
| T58 | 699 | 2 | 35 | 25.7 | 62.9 | 55 | 10.9 | 70.9 | 49 | 18.4 | 67.3 |
| T85 | 696 | 2 | 51 | 45.1 | 39.2 | 60 | 33.3 | 55.0 | 51 | 51.0 | 43.1 |
| Z73 | 695 | 2 | 13 | 23.1 | 61.5 | 21 | 28.6 | 66.7 | 14 | 64.3 | 21.4 |
| K09 | 687 | 2 | 26 | 42.3 | 42.3 | 23 | 21.7 | 78.3 | 33 | 24.2 | 66.7 |
| M89 | 685 | 2 | 26 | 30.8 | 69.2 | 21 | 9.5 | 81.0 | 14 | 21.4 | 71.4 |
| O66 | 673 | 2 | 70 | 21.4 | 52.9 | 73 | 19.2 | 57.5 | 62 | 25.8 | 59.7 |
| H05 | 666 | 2 | 35 | 22.9 | 74.3 | 46 | 15.2 | 58.7 | 29 | 31.0 | 65.5 |
| I27 | 666 | 4 | 70 | 18.6 | 62.9 | 121 | 17.4 | 66.1 | 102 | 19.6 | 65.7 |
| I85 | 662 | 2 | 77 | 35.1 | 58.4 | 127 | 18.9 | 66.1 | 123 | 23.6 | 64.2 |
| K71 | 658 | 2 | 52 | 19.2 | 69.2 | 96 | 15.6 | 56.2 | 98 | 12.2 | 70.4 |
| L55 | 651 | 4 | 13 | 30.8 | 69.2 | 15 | 33.3 | 60.0 | 10 | 40.0 | 60.0 |
| I72 | 646 | 2 | 60 | 20.0 | 55.0 | 80 | 50.0 | 32.5 | 72 | 18.1 | 63.9 |
| M87 | 644 | 2 | 45 | 20.0 | 64.4 | 46 | 19.6 | 69.6 | 49 | 20.4 | 67.3 |
| G52 | 640 | 4 | 28 | 25.0 | 71.4 | 35 | 22.9 | 65.7 | 17 | 35.3 | 58.8 |
| D33 | 630 | 2 | 47 | 19.1 | 57.5 | 55 | 23.6 | 49.1 | 70 | 27.1 | 54.3 |
| F11 | 630 | 2 | 32 | 40.6 | 50.0 | 39 | 28.2 | 66.7 | 38 | 26.3 | 65.8 |
| R35 | 626 | 2 | 24 | 33.3 | 66.7 | 28 | 32.1 | 46.4 | 19 | 15.8 | 57.9 |
| T20 | 625 | 2 | 21 | 23.8 | 66.7 | 32 | 18.8 | 75.0 | 25 | 28.0 | 72.0 |
| M66 | 625 | 2 | 31 | 16.1 | 80.7 | 35 | 28.6 | 68.6 | 28 | 32.1 | 60.7 |
| I60 | 622 | 3 | 61 | 23.0 | 49.2 | 105 | 20.0 | 68.6 | 105 | 19.0 | 65.7 |
| N36 | 620 | 2 | 32 | 25.0 | 59.4 | 42 | 21.4 | 54.8 | 30 | 26.7 | 56.7 |
| F44 | 611 | 3 | 35 | 31.4 | 65.7 | 49 | 18.4 | 75.5 | 41 | 19.5 | 78.0 |
| D43 | 605 | 2 | 54 | 24.1 | 64.8 | 84 | 25.0 | 69.0 | 111 | 23.4 | 69.4 |
| L87 | 605 | 5 | 8 | 50.0 | 37.5 | 14 | 21.4 | 71.4 | 7 | 28.6 | 57.1 |
| C56 | 604 | 2 | 82 | 26.8 | 69.5 | 112 | 30.4 | 64.3 | 128 | 23.4 | 68.8 |
| S12 | 603 | 2 | 40 | 35.0 | 55.0 | 68 | 10.3 | 79.4 | 78 | 11.5 | 70.5 |
| A59 | 602 | 3 | 24 | 37.5 | 50.0 | 31 | 32.3 | 64.5 | 32 | 31.2 | 50.0 |
| A02 | 600 | 2 | 47 | 31.9 | 59.6 | 60 | 26.7 | 61.7 | 50 | 26.0 | 64.0 |
| F12 | 598 | 2 | 33 | 27.3 | 54.5 | 47 | 23.4 | 63.8 | 50 | 18.0 | 48.0 |
| F99 | 597 | 2 | 45 | 31.1 | 37.8 | 81 | 7.4 | 87.7 | 80 | 16.2 | 80.0 |
| O32 | 595 | 2 | 60 | 21.7 | 68.3 | 83 | 21.7 | 69.9 | 81 | 23.5 | 59.3 |
| D86 | 592 | 2 | 65 | 29.2 | 60.0 | 63 | 27.0 | 54.0 | 75 | 25.3 | 64.0 |
| C80 | 591 | 2 | 93 | 31.2 | 62.4 | 115 | 22.6 | 73.9 | 131 | 33.6 | 64.1 |
| L86 | 587 | 5 | 8 | 37.5 | 37.5 | 9 | 22.2 | 55.6 | 11 | 18.2 | 81.8 |
| G80 | 581 | 4 | 11 | 18.2 | 81.8 | 14 | 21.4 | 78.6 | 10 | 30.0 | 70.0 |
| H17 | 580 | 2 | 26 | 42.3 | 34.6 | 26 | 7.7 | 73.1 | 16 | 31.2 | 56.2 |
| H82 | 574 | 2 | 29 | 34.5 | 58.6 | 31 | 32.3 | 61.3 | 25 | 28.0 | 60.0 |
| H70 | 570 | 2 | 52 | 40.4 | 59.6 | 64 | 32.8 | 45.3 | 55 | 34.5 | 58.2 |
| L44 | 570 | 2 | 19 | 57.9 | 42.1 | 19 | 63.2 | 36.8 | 18 | 77.8 | 22.2 |
| N12 | 566 | 2 | 44 | 38.6 | 54.5 | 55 | 5.4 | 72.7 | 43 | 9.3 | 76.7 |
| G35 | 561 | 2 | 52 | 26.9 | 57.7 | 50 | 18.0 | 70.0 | 66 | 28.8 | 56.1 |
| J14 | 546 | 3 | 63 | 38.1 | 44.4 | 98 | 18.4 | 49.0 | 87 | 24.1 | 44.8 |
| O44 | 542 | 2 | 47 | 12.8 | 59.6 | 65 | 35.4 | 52.3 | 89 | 16.9 | 56.2 |
| D11 | 541 | 2 | 38 | 31.6 | 52.6 | 31 | 45.2 | 51.6 | 48 | 31.2 | 58.3 |
| B89 | 537 | 5 | 8 | 37.5 | 62.5 | 10 | 50.0 | 50.0 | 7 | 71.4 | 28.6 |
| Q61 | 536 | 2 | 25 | 20.0 | 76.0 | 37 | 16.2 | 73.0 | 22 | 27.3 | 68.2 |
| G55 | 536 | 2 | 29 | 34.5 | 65.5 | 38 | 26.3 | 63.2 | 33 | 36.4 | 54.5 |
| L94 | 536 | 2 | 22 | 45.5 | 36.4 | 24 | 29.2 | 50.0 | 22 | 45.5 | 36.4 |
| M11 | 535 | 2 | 34 | 23.5 | 67.7 | 50 | 16.0 | 58.0 | 27 | 25.9 | 59.3 |
| S21 | 533 | 2 | 32 | 53.1 | 43.8 | 35 | 57.1 | 40.0 | 36 | 22.2 | 72.2 |
| Z89 | 530 | 2 | 73 | 50.7 | 32.9 | 66 | 12.1 | 71.2 | 48 | 22.9 | 60.4 |
| K45 | 529 | 2 | 30 | 13.3 | 70.0 | 37 | 16.2 | 81.1 | 20 | 30.0 | 65.0 |
| R44 | 526 | 2 | 26 | 19.2 | 76.9 | 33 | 18.2 | 66.7 | 24 | 12.5 | 83.3 |
| I97 | 524 | 5 | 31 | 35.5 | 48.4 | 21 | 14.3 | 71.4 | 9 | 22.2 | 77.8 |
| A40 | 522 | 2 | 58 | 51.7 | 37.9 | 93 | 14.0 | 63.4 | 87 | 11.5 | 69.0 |
| G96 | 520 | 2 | 37 | 35.1 | 64.9 | 49 | 18.4 | 65.3 | 44 | 31.8 | 63.6 |
| E23 | 515 | 2 | 45 | 24.4 | 53.3 | 52 | 23.1 | 67.3 | 51 | 31.4 | 56.9 |
| N35 | 515 | 2 | 59 | 25.4 | 50.9 | 62 | 22.6 | 66.1 | 46 | 30.4 | 60.9 |
| L68 | 505 | 2 | 14 | 15.4 | 69.2 | 27 | 7.4 | 66.7 | 24 | 25.0 | 58.3 |
| T39 | 505 | 2 | 40 | 85.0 | 2.5 | 64 | 34.4 | 48.4 | 69 | 8.7 | 71.0 |
| N02 | 504 | 3 | 38 | 31.6 | 55.3 | 41 | 24.4 | 63.4 | 27 | 25.9 | 48.1 |
| M00 | 502 | 2 | 67 | 41.8 | 43.3 | 72 | 31.9 | 55.6 | 76 | 25.0 | 55.3 |
| O73 | 502 | 2 | 61 | 19.7 | 75.4 | 51 | 13.7 | 62.8 | 55 | 10.9 | 67.3 |
| D28 | 500 | 2 | 39 | 25.6 | 56.4 | 42 | 28.6 | 69.0 | 31 | 38.7 | 61.3 |
| E34 | 494 | 2 | 18 | 50.0 | 44.4 | 22 | 13.6 | 72.7 | 17 | 23.5 | 64.7 |
| D36 | 493 | 2 | 27 | 33.3 | 51.9 | 33 | 15.2 | 84.8 | 26 | 19.2 | 80.8 |
| I05 | 492 | 2 | 37 | 24.3 | 54.0 | 59 | 6.8 | 79.7 | 59 | 15.2 | 67.8 |
| F79 | 492 | 2 | 9 | 44.4 | 55.6 | 21 | 42.9 | 57.1 | 12 | 50.0 | 50.0 |
| C53 | 489 | 4 | 66 | 36.4 | 54.5 | 77 | 36.4 | 55.8 | 108 | 27.8 | 65.7 |
| F29 | 489 | 2 | 39 | 33.3 | 43.6 | 54 | 33.3 | 59.3 | 57 | 43.9 | 52.6 |
| Q18 | 486 | 4 | 42 | 28.6 | 66.7 | 36 | 38.9 | 61.1 | 37 | 27.0 | 70.3 |
| G21 | 486 | 2 | 35 | 22.9 | 62.9 | 45 | 13.3 | 82.2 | 31 | 29.0 | 54.8 |
| Q65 | 484 | 2 | 14 | 21.4 | 50.0 | 9 | 33.3 | 66.7 | 19 | 26.3 | 73.7 |
| C71 | 481 | 2 | 53 | 30.2 | 62.3 | 86 | 26.7 | 65.1 | 104 | 30.8 | 59.6 |
| Q23 | 479 | 2 | 28 | 17.9 | 71.4 | 37 | 13.5 | 81.1 | 49 | 16.3 | 61.2 |
| S89 | 479 | 5 | 13 | 53.9 | 46.1 | 16 | 18.8 | 81.2 | 6 | 33.3 | 66.7 |
| M01 | 478 | 3 | 30 | 33.3 | 56.7 | 34 | 35.3 | 52.9 | 28 | 32.1 | 60.7 |
| N03 | 476 | 2 | 49 | 28.6 | 57.1 | 63 | 19.0 | 73.0 | 70 | 30.0 | 62.9 |
| E44 | 474 | 2 | 86 | 34.9 | 60.5 | 86 | 18.6 | 59.3 | 80 | 15.0 | 68.8 |
| E02 | 469 | 2 | 27 | 33.3 | 51.9 | 23 | 17.4 | 73.9 | 15 | 13.3 | 66.7 |
| K28 | 467 | 2 | 47 | 19.1 | 68.1 | 57 | 24.6 | 52.6 | 43 | 25.6 | 58.1 |
| Z93 | 466 | 2 | 89 | 30.3 | 61.8 | 107 | 6.5 | 84.1 | 118 | 11.9 | 82.2 |
| H67 | 466 | 2 | 12 | 41.7 | 58.3 | 27 | 25.9 | 70.4 | 11 | 54.5 | 45.5 |
| N00 | 465 | 2 | 35 | 20.0 | 68.6 | 41 | 9.8 | 80.5 | 38 | 13.2 | 76.3 |
| D29 | 465 | 2 | 21 | 19.0 | 66.7 | 26 | 26.9 | 61.5 | 28 | 14.3 | 67.9 |
| I77 | 465 | 2 | 40 | 27.5 | 67.5 | 57 | 17.5 | 77.2 | 48 | 14.6 | 79.2 |
| H49 | 465 | 2 | 46 | 37.0 | 41.3 | 55 | 14.6 | 67.3 | 55 | 23.6 | 61.8 |
| Q05 | 465 | 2 | 10 | 30.0 | 70.0 | 23 | 30.4 | 69.6 | 24 | 25.0 | 66.7 |
| T45 | 464 | 2 | 58 | 37.9 | 56.9 | 93 | 20.4 | 58.1 | 68 | 16.2 | 64.7 |
| D73 | 463 | 2 | 62 | 30.6 | 51.6 | 80 | 11.2 | 70.0 | 80 | 15.0 | 72.5 |
| E88 | 462 | 2 | 29 | 27.6 | 62.1 | 42 | 14.3 | 83.3 | 29 | 24.1 | 62.1 |
| M99 | 460 | 2 | 21 | 42.9 | 52.4 | 11 | 63.6 | 36.4 | 11 | 54.5 | 36.4 |
| R09 | 459 | 2 | 24 | 25.0 | 75.0 | 40 | 7.5 | 87.5 | 24 | 25.0 | 66.7 |
| S56 | 456 | 5 | 20 | 50.0 | 45.0 | 31 | 19.4 | 80.7 | 27 | 29.6 | 70.4 |
| C91 | 452 | 2 | 48 | 20.8 | 72.9 | 62 | 22.6 | 64.5 | 75 | 30.7 | 62.7 |
| A54 | 451 | 2 | 13 | 61.5 | 30.8 | 17 | 41.2 | 52.9 | 16 | 31.2 | 56.2 |
| A37 | 450 | 5 | 30 | 36.7 | 53.3 | 32 | 43.8 | 50.0 | 17 | 35.3 | 52.9 |
| I31 | 443 | 5 | 71 | 25.4 | 56.3 | 94 | 17.0 | 58.5 | 106 | 16.0 | 65.1 |
| S29 | 435 | 2 | 11 | 45.5 | 45.5 | 24 | 12.5 | 83.3 | 11 | 27.3 | 72.7 |
| I40 | 430 | 2 | 47 | 21.3 | 66.0 | 63 | 12.7 | 66.7 | 59 | 6.8 | 67.8 |
| I79 | 430 | 2 | 30 | 33.3 | 66.7 | 39 | 23.1 | 71.8 | 22 | 22.7 | 72.7 |
| L22 | 423 | 5 | 4 | 50.0 | 50.0 | 8 | 25.0 | 75.0 | 5 | 40.0 | 60.0 |
| S49 | 418 | 2 | 9 | 33.3 | 66.7 | 20 | 30.0 | 70.0 | 10 | 60.0 | 40.0 |
| E27 | 417 | 2 | 43 | 39.5 | 51.2 | 51 | 15.7 | 70.6 | 31 | 32.3 | 51.6 |
| G72 | 414 | 2 | 47 | 25.5 | 63.8 | 61 | 18.0 | 65.6 | 81 | 12.3 | 75.3 |
| D80 | 412 | 2 | 63 | 27.0 | 60.3 | 69 | 11.6 | 68.1 | 62 | 14.5 | 80.7 |
| D03 | 412 | 2 | 20 | 70.0 | 30.0 | 25 | 28.0 | 72.0 | 23 | 26.1 | 69.6 |
| R76 | 410 | 2 | 30 | 43.3 | 43.3 | 35 | 8.6 | 51.4 | 22 | 18.2 | 59.1 |
| S36 | 409 | 3 | 46 | 37.0 | 54.4 | 75 | 10.7 | 76.0 | 74 | 21.6 | 67.6 |
| C19 | 409 | 2 | 65 | 30.8 | 50.8 | 85 | 34.1 | 57.6 | 110 | 35.5 | 58.2 |
| H55 | 406 | 4 | 16 | 37.5 | 62.5 | 21 | 19.0 | 71.4 | 17 | 11.8 | 82.3 |
| C22 | 405 | 5 | 71 | 32.4 | 54.9 | 89 | 27.0 | 59.5 | 110 | 28.2 | 65.4 |
| C83 | 404 | 3 | 87 | 36.8 | 52.9 | 94 | 25.5 | 64.9 | 105 | 32.4 | 64.8 |
| N08 | 401 | 2 | 45 | 37.8 | 51.1 | 58 | 22.4 | 65.5 | 47 | 25.5 | 61.7 |
| F25 | 399 | 2 | 19 | 68.4 | 26.3 | 28 | 32.1 | 57.1 | 30 | 43.3 | 50.0 |
| Q63 | 398 | 2 | 28 | 28.6 | 53.6 | 33 | 12.1 | 66.7 | 29 | 17.2 | 62.1 |
| L62 | 397 | 5 | 15 | 33.3 | 66.7 | 13 | 23.1 | 76.9 | 5 | 40.0 | 60.0 |
| A15 | 393 | 2 | 58 | 32.8 | 55.2 | 58 | 34.5 | 56.9 | 67 | 23.9 | 50.8 |
| S39 | 390 | 5 | 17 | 47.1 | 47.1 | 19 | 31.6 | 68.4 | 11 | 36.4 | 63.6 |
| E72 | 389 | 2 | 31 | 19.4 | 71.0 | 51 | 5.9 | 70.6 | 25 | 20.0 | 76.0 |
| F15 | 387 | 5 | 30 | 53.3 | 40.0 | 40 | 32.5 | 55.0 | 44 | 27.3 | 63.6 |
| O61 | 379 | 4 | 55 | 25.4 | 70.9 | 62 | 14.5 | 74.2 | 56 | 19.6 | 64.3 |
| O45 | 379 | 5 | 55 | 34.5 | 49.1 | 57 | 28.1 | 47.4 | 48 | 22.9 | 58.3 |
| B49 | 378 | 5 | 9 | 55.6 | 44.4 | 12 | 50.0 | 50.0 | 8 | 75.0 | 25.0 |
| M14 | 376 | 4 | 13 | 38.5 | 53.9 | 17 | 17.6 | 47.1 | 8 | 37.5 | 50.0 |
| Z21 | 376 | 4 | 15 | 26.7 | 66.7 | 20 | 25.0 | 75.0 | 19 | 21.0 | 63.2 |
| T68 | 371 | 2 | 44 | 27.3 | 61.4 | 68 | 1.5 | 73.5 | 66 | 4.6 | 77.3 |
| L13 | 368 | 5 | 16 | 31.2 | 68.8 | 27 | 22.2 | 63.0 | 11 | 63.6 | 27.3 |
| E65 | 367 | 3 | 11 | 27.3 | 63.6 | 7 | 14.3 | 85.7 | 5 | 60.0 | 40.0 |
| G92 | 367 | 2 | 71 | 36.6 | 63.4 | 86 | 26.7 | 50.0 | 76 | 10.5 | 75.0 |
| A64 | 367 | 2 | 7 | 57.1 | 28.6 | 19 | 52.6 | 42.1 | 13 | 46.1 | 46.1 |
| T46 | 366 | 2 | 46 | 23.9 | 63.0 | 70 | 12.9 | 65.7 | 59 | 17.0 | 71.2 |
| T82 | 363 | 2 | 55 | 36.4 | 60.0 | 67 | 22.4 | 55.2 | 56 | 21.4 | 53.6 |
| F94 | 361 | 2 | 13 | 46.1 | 53.9 | 19 | 47.4 | 47.4 | 20 | 50.0 | 50.0 |
| H30 | 356 | 2 | 36 | 47.2 | 38.9 | 36 | 41.7 | 47.2 | 35 | 40.0 | 48.6 |
| A26 | 354 | 4 | 19 | 5.3 | 89.5 | 29 | 17.2 | 62.1 | 11 | 45.5 | 54.5 |
| R48 | 352 | 2 | 25 | 24.0 | 36.0 | 28 | 21.4 | 46.4 | 25 | 24.0 | 56.0 |
| N29 | 352 | 2 | 20 | 45.0 | 55.0 | 32 | 15.6 | 78.1 | 18 | 27.8 | 55.6 |
| E30 | 351 | 2 | 10 | 30.0 | 70.0 | 17 | 23.5 | 76.5 | 16 | 25.0 | 75.0 |
| K41 | 351 | 3 | 37 | 37.8 | 51.4 | 39 | 15.4 | 74.4 | 39 | 18.0 | 66.7 |
| G91 | 351 | 2 | 38 | 18.4 | 60.5 | 53 | 15.1 | 71.7 | 54 | 20.4 | 61.1 |
| L41 | 351 | 2 | 18 | 50.0 | 50.0 | 24 | 29.2 | 62.5 | 14 | 57.1 | 42.9 |
| S37 | 346 | 2 | 50 | 42.0 | 50.0 | 66 | 22.7 | 63.6 | 69 | 21.7 | 66.7 |
| K36 | 346 | 2 | 50 | 22.0 | 58.0 | 50 | 18.0 | 68.0 | 51 | 21.6 | 60.8 |
| D45 | 344 | 2 | 32 | 25.0 | 62.5 | 36 | 22.2 | 61.1 | 38 | 36.8 | 50.0 |
| B25 | 341 | 4 | 95 | 32.6 | 51.6 | 78 | 16.7 | 68.0 | 84 | 14.3 | 70.2 |
| M91 | 341 | 2 | 22 | 27.3 | 68.2 | 18 | 27.8 | 72.2 | 11 | 54.5 | 36.4 |
| M31 | 341 | 2 | 70 | 30.0 | 64.3 | 88 | 17.0 | 77.3 | 92 | 21.7 | 71.7 |
| J86 | 341 | 2 | 72 | 36.1 | 54.2 | 92 | 27.2 | 57.6 | 96 | 20.8 | 59.4 |
| D61 | 338 | 2 | 92 | 47.8 | 45.6 | 91 | 18.7 | 71.4 | 105 | 21.0 | 69.5 |
| D55 | 336 | 2 | 14 | 21.4 | 64.3 | 18 | 16.7 | 83.3 | 8 | 37.5 | 50.0 |
| B99 | 332 | 2 | 37 | 51.4 | 43.2 | 49 | 14.3 | 77.6 | 42 | 19.0 | 73.8 |
| I81 | 331 | 2 | 50 | 30.0 | 50.0 | 89 | 22.5 | 50.6 | 76 | 31.6 | 65.8 |
| S67 | 330 | 4 | 21 | 47.6 | 52.4 | 29 | 37.9 | 62.1 | 25 | 48.0 | 52.0 |
| E67 | 329 | 2 | 12 | 41.7 | 58.3 | 16 | 18.8 | 75.0 | 8 | 12.5 | 75.0 |
| A06 | 328 | 2 | 22 | 36.4 | 54.5 | 26 | 26.9 | 53.9 | 19 | 42.1 | 52.6 |
| R58 | 326 | 2 | 26 | 30.8 | 61.5 | 42 | 23.8 | 66.7 | 33 | 36.4 | 60.6 |
| N44 | 324 | 5 | 28 | 25.0 | 53.6 | 32 | 21.9 | 65.6 | 30 | 20.0 | 70.0 |
| H71 | 320 | 2 | 23 | 47.8 | 43.5 | 18 | 61.1 | 38.9 | 28 | 35.7 | 60.7 |
| H51 | 319 | 5 | 12 | 25.0 | 75.0 | 11 | 18.2 | 81.8 | 9 | 22.2 | 77.8 |
| L52 | 318 | 2 | 27 | 25.9 | 74.1 | 44 | 6.8 | 77.3 | 29 | 41.4 | 48.3 |
| G37 | 318 | 2 | 39 | 10.3 | 71.8 | 36 | 19.4 | 63.9 | 46 | 26.1 | 65.2 |
| N15 | 318 | 2 | 47 | 36.2 | 42.5 | 70 | 7.1 | 67.1 | 52 | 9.6 | 71.2 |
| Q55 | 316 | 2 | 6 | 66.7 | 33.3 | 5 | 60.0 | 40.0 | 7 | 28.6 | 71.4 |
| I66 | 315 | 2 | 36 | 41.7 | 47.2 | 75 | 22.7 | 62.7 | 61 | 37.7 | 44.3 |
| G71 | 315 | 2 | 18 | 22.2 | 77.8 | 19 | 36.8 | 57.9 | 21 | 38.1 | 57.1 |
| K46 | 315 | 4 | 14 | 14.3 | 64.3 | 26 | 15.4 | 73.1 | 13 | 38.5 | 61.5 |
| N26 | 315 | 2 | 40 | 27.5 | 60.0 | 50 | 22.0 | 52.0 | 39 | 15.4 | 61.5 |
| Q76 | 309 | 4 | 9 | 44.4 | 55.6 | 11 | 45.5 | 45.5 | 8 | 75.0 | 25.0 |
| L51 | 306 | 3 | 29 | 10.3 | 89.7 | 24 | 8.3 | 75.0 | 16 | 43.8 | 56.2 |
| H31 | 305 | 2 | 19 | 31.6 | 47.4 | 22 | 31.8 | 63.6 | 13 | 46.1 | 46.1 |
| F72 | 305 | 2 | 4 | 50.0 | 50.0 | 8 | 12.5 | 87.5 | 2 | 50.0 | 50.0 |
| S59 | 304 | 4 | 8 | 37.5 | 62.5 | 12 | 25.0 | 75.0 | 6 | 33.3 | 66.7 |
| B20 | 297 | 2 | 39 | 18.0 | 71.8 | 67 | 14.9 | 68.7 | 76 | 15.8 | 73.7 |
| Z72 | 297 | 5 | 15 | 13.3 | 53.3 | 13 | 15.4 | 69.2 | 3 | 33.3 | 66.7 |
| G61 | 295 | 2 | 43 | 30.2 | 62.8 | 56 | 21.4 | 58.9 | 61 | 23.0 | 67.2 |
| I62 | 295 | 2 | 50 | 20.0 | 72.0 | 72 | 18.1 | 66.7 | 66 | 22.7 | 66.7 |
| J80 | 294 | 2 | 59 | 55.9 | 37.3 | 77 | 35.1 | 49.4 | 76 | 29.0 | 56.6 |
| F82 | 293 | 5 | 2 | 50.0 | 50.0 | 8 | 37.5 | 62.5 | 7 | 42.9 | 57.1 |
| T75 | 292 | 4 | 21 | 33.3 | 61.9 | 21 | 19.0 | 81.0 | 16 | 12.5 | 81.2 |
| S11 | 290 | 2 | 22 | 40.9 | 59.1 | 24 | 16.7 | 83.3 | 22 | 18.2 | 81.8 |
| R63 | 282 | 2 | 19 | 26.3 | 73.7 | 33 | 9.1 | 81.8 | 19 | 5.3 | 79.0 |
| O63 | 277 | 2 | 51 | 27.4 | 52.9 | 50 | 16.0 | 66.0 | 42 | 21.4 | 61.9 |
| B17 | 277 | 2 | 27 | 22.2 | 70.4 | 39 | 20.5 | 69.2 | 39 | 23.1 | 59.0 |
| D01 | 276 | 2 | 28 | 25.0 | 64.3 | 40 | 27.5 | 60.0 | 51 | 21.6 | 72.6 |
| O43 | 273 | 2 | 43 | 23.3 | 69.8 | 59 | 28.8 | 61.0 | 57 | 29.8 | 59.6 |
| C85 | 272 | 2 | 63 | 30.2 | 57.1 | 87 | 27.6 | 67.8 | 108 | 32.4 | 62.0 |
| T31 | 271 | 2 | 30 | 66.7 | 33.3 | 45 | 22.2 | 68.9 | 48 | 20.8 | 79.2 |
| T80 | 268 | 2 | 35 | 34.3 | 57.1 | 44 | 13.6 | 75.0 | 34 | 11.8 | 79.4 |
| G70 | 267 | 2 | 40 | 25.0 | 57.5 | 30 | 30.0 | 60.0 | 45 | 31.1 | 53.3 |
| D89 | 266 | 2 | 35 | 31.4 | 57.1 | 36 | 27.8 | 72.2 | 22 | 36.4 | 63.6 |
| Q17 | 265 | 4 | 4 | 75.0 | 25.0 | 9 | 44.4 | 55.6 | 12 | 41.7 | 58.3 |
| G64 | 265 | 2 | 10 | 30.0 | 70.0 | 14 | 21.4 | 78.6 | 6 | 33.3 | 66.7 |
| D46 | 264 | 2 | 50 | 22.0 | 66.0 | 60 | 26.7 | 50.0 | 61 | 31.1 | 63.9 |
| I08 | 263 | 2 | 25 | 12.0 | 80.0 | 44 | 6.8 | 81.8 | 33 | 12.1 | 78.8 |
| O46 | 261 | 2 | 35 | 54.3 | 45.7 | 50 | 24.0 | 60.0 | 53 | 45.3 | 43.4 |
| T86 | 261 | 3 | 103 | 80.6 | 18.4 | 78 | 30.8 | 68.0 | 73 | 8.2 | 87.7 |
| F89 | 260 | 2 | 7 | 71.4 | 28.6 | 9 | 77.8 | 22.2 | 10 | 90.0 | 10.0 |
| I33 | 257 | 2 | 63 | 22.2 | 65.1 | 84 | 15.5 | 57.1 | 98 | 15.3 | 66.3 |
| Q28 | 257 | 2 | 22 | 31.8 | 59.1 | 25 | 24.0 | 68.0 | 23 | 21.7 | 73.9 |
| Q87 | 257 | 4 | 7 | 71.4 | 28.6 | 13 | 46.1 | 53.9 | 14 | 42.9 | 50.0 |
| B44 | 256 | 2 | 69 | 26.1 | 49.3 | 69 | 18.8 | 65.2 | 68 | 22.1 | 64.7 |
| C90 | 256 | 3 | 63 | 27.0 | 49.2 | 77 | 15.6 | 72.7 | 109 | 26.6 | 70.6 |
| O25 | 253 | 2 | 28 | 21.4 | 57.1 | 23 | 13.0 | 82.6 | 64 | 7.8 | 56.2 |
| J85 | 252 | 3 | 52 | 38.5 | 57.7 | 84 | 26.2 | 45.2 | 77 | 31.2 | 49.4 |
| H28 | 252 | 2 | 13 | 61.5 | 38.5 | 20 | 40.0 | 55.0 | 12 | 41.7 | 50.0 |
| G95 | 252 | 2 | 23 | 21.7 | 69.6 | 44 | 11.4 | 75.0 | 46 | 15.2 | 67.4 |
| L10 | 252 | 4 | 14 | 28.6 | 50.0 | 27 | 22.2 | 77.8 | 15 | 33.3 | 66.7 |
| C26 | 251 | 2 | 34 | 20.6 | 67.7 | 67 | 28.4 | 62.7 | 79 | 31.6 | 63.3 |
| Q43 | 251 | 2 | 28 | 32.1 | 53.6 | 43 | 11.6 | 74.4 | 32 | 3.1 | 84.4 |
| N98 | 250 | 2 | 56 | 46.4 | 42.9 | 44 | 38.6 | 59.1 | 75 | 21.3 | 60.0 |
| L66 | 249 | 5 | 6 | 66.7 | 33.3 | 8 | 50.0 | 50.0 | 5 | 80.0 | 20.0 |
| S64 | 249 | 2 | 27 | 51.9 | 48.1 | 28 | 42.9 | 57.1 | 28 | 42.9 | 57.1 |
| S79 | 247 | 5 | 12 | 25.0 | 75.0 | 15 | 26.7 | 73.3 | 7 | 42.9 | 57.1 |
| R77 | 244 | 2 | 45 | 17.8 | 66.7 | 67 | 16.4 | 64.2 | 68 | 16.2 | 76.5 |
| I06 | 242 | 2 | 24 | 12.5 | 70.8 | 30 | 13.3 | 63.3 | 21 | 19.0 | 71.4 |
| B70 | 240 | 2 | 12 | 41.7 | 58.3 | 10 | 40.0 | 60.0 | 7 | 57.1 | 42.9 |
| T33 | 239 | 5 | 8 | 25.0 | 75.0 | 11 | 27.3 | 72.7 | 6 | 33.3 | 66.7 |
| C32 | 237 | 5 | 33 | 36.4 | 60.6 | 46 | 47.8 | 47.8 | 85 | 34.1 | 57.6 |
| T19 | 227 | 5 | 7 | 14.3 | 85.7 | 10 | 30.0 | 70.0 | 7 | 28.6 | 71.4 |
| F21 | 225 | 5 | 4 | 50.0 | 50.0 | 6 | 66.7 | 33.3 | 5 | 80.0 | 20.0 |
| L93 | 224 | 5 | 13 | 61.5 | 38.5 | 20 | 45.0 | 55.0 | 16 | 50.0 | 43.8 |
| C73 | 223 | 2 | 35 | 37.1 | 54.3 | 35 | 28.6 | 65.7 | 46 | 37.0 | 47.8 |
| F28 | 223 | 3 | 20 | 25.0 | 65.0 | 23 | 43.5 | 47.8 | 18 | 38.9 | 61.1 |
| C15 | 222 | 2 | 35 | 20.0 | 68.6 | 53 | 37.7 | 58.5 | 84 | 35.7 | 61.9 |
| B60 | 222 | 4 | 11 | 36.4 | 63.6 | 9 | 33.3 | 66.7 | 6 | 50.0 | 50.0 |
| M63 | 220 | 2 | 28 | 14.3 | 85.7 | 26 | 15.4 | 80.8 | 24 | 16.7 | 75.0 |
| G04 | 219 | 3 | 54 | 40.7 | 51.9 | 67 | 38.8 | 47.8 | 70 | 17.1 | 68.6 |
| A68 | 219 | 4 | 6 | 33.3 | 66.7 | 8 | 25.0 | 75.0 | 3 | 66.7 | 33.3 |
| T65 | 218 | 2 | 13 | 38.5 | 61.5 | 20 | 15.0 | 75.0 | 14 | 21.4 | 78.6 |
| I41 | 214 | 2 | 22 | 27.3 | 59.1 | 27 | 22.2 | 74.1 | 23 | 26.1 | 60.9 |
| G59 | 213 | 5 | 4 | 75.0 | 25.0 | 15 | 33.3 | 66.7 | 4 | 100.0 | 0.0 |
| I30 | 212 | 2 | 37 | 24.3 | 59.5 | 54 | 13.0 | 59.3 | 53 | 15.1 | 71.7 |
| D34 | 212 | 2 | 26 | 26.9 | 65.4 | 24 | 25.0 | 62.5 | 27 | 37.0 | 51.9 |
| E01 | 212 | 5 | 10 | 10.0 | 90.0 | 16 | 25.0 | 68.8 | 7 | 57.1 | 42.9 |
| S19 | 212 | 4 | 8 | 37.5 | 62.5 | 10 | 30.0 | 70.0 | 8 | 37.5 | 62.5 |
| O85 | 211 | 2 | 67 | 43.3 | 44.8 | 57 | 28.1 | 57.9 | 44 | 27.3 | 56.8 |
| F30 | 210 | 2 | 5 | 40.0 | 60.0 | 15 | 40.0 | 60.0 | 16 | 25.0 | 75.0 |
| C49 | 209 | 2 | 28 | 46.4 | 50.0 | 34 | 47.1 | 52.9 | 60 | 35.0 | 60.0 |
| G12 | 209 | 2 | 25 | 16.0 | 76.0 | 36 | 19.4 | 63.9 | 44 | 9.1 | 77.3 |
| Q25 | 207 | 2 | 8 | 50.0 | 50.0 | 9 | 33.3 | 55.6 | 10 | 30.0 | 70.0 |
| A05 | 207 | 2 | 10 | 10.0 | 90.0 | 13 | 7.7 | 92.3 | 10 | 10.0 | 90.0 |
| T50 | 206 | 2 | 23 | 43.5 | 47.8 | 31 | 16.1 | 67.7 | 25 | 24.0 | 68.0 |
| H95 | 204 | 2 | 7 | 71.4 | 28.6 | 10 | 30.0 | 70.0 | 7 | 42.9 | 57.1 |
| G99 | 204 | 2 | 24 | 33.3 | 62.5 | 33 | 18.2 | 78.8 | 18 | 11.1 | 83.3 |
| M32 | 204 | 2 | 25 | 32.0 | 48.0 | 28 | 46.4 | 50.0 | 21 | 47.6 | 52.4 |
| F09 | 204 | 2 | 25 | 24.0 | 72.0 | 29 | 48.3 | 48.3 | 11 | 9.1 | 90.9 |
| L12 | 202 | 2 | 24 | 41.7 | 41.7 | 28 | 28.6 | 60.7 | 28 | 39.3 | 46.4 |
| H59 | 201 | 4 | 20 | 55.0 | 35.0 | 16 | 37.5 | 56.2 | 8 | 75.0 | 12.5 |
| R80 | 201 | 2 | 13 | 38.5 | 61.5 | 29 | 17.2 | 75.9 | 20 | 30.0 | 45.0 |
| I68 | 199 | 2 | 16 | 37.5 | 62.5 | 26 | 23.1 | 73.1 | 16 | 25.0 | 75.0 |
| A00 | 199 | 3 | 1 | 100.0 | 0.0 | 6 | 16.7 | 83.3 | 2 | 50.0 | 50.0 |
| C21 | 199 | 2 | 38 | 39.5 | 47.4 | 40 | 27.5 | 72.5 | 68 | 33.8 | 60.3 |
| L99 | 199 | 5 | 3 | 33.3 | 66.7 | 11 | 9.1 | 90.9 | 1 | 100.0 | 0.0 |
| J95 | 196 | 2 | 36 | 33.3 | 47.2 | 52 | 13.5 | 71.2 | 34 | 17.6 | 73.5 |
| A31 | 194 | 2 | 18 | 50.0 | 50.0 | 24 | 54.2 | 37.5 | 14 | 57.1 | 35.7 |
| A87 | 194 | 2 | 37 | 35.1 | 48.6 | 51 | 9.8 | 78.4 | 46 | 19.6 | 58.7 |
| B19 | 191 | 2 | 16 | 31.2 | 68.8 | 38 | 21.0 | 60.5 | 30 | 26.7 | 66.7 |
| L74 | 190 | 5 | 1 | 100.0 | 0.0 | 2 | 50.0 | 50.0 | 1 | 100.0 | 0.0 |
| C92 | 190 | 2 | 60 | 31.7 | 50.0 | 72 | 27.8 | 62.5 | 81 | 23.5 | 64.2 |
| Q10 | 187 | 5 | 6 | 50.0 | 50.0 | 8 | 37.5 | 62.5 | 9 | 44.4 | 55.6 |
| S44 | 187 | 2 | 21 | 38.1 | 61.9 | 24 | 29.2 | 70.8 | 20 | 30.0 | 70.0 |
| I24 | 185 | 2 | 31 | 58.1 | 29.0 | 42 | 23.8 | 59.5 | 21 | 19.0 | 71.4 |
| I09 | 185 | 3 | 7 | 42.9 | 57.1 | 16 | 18.8 | 81.2 | 6 | 66.7 | 33.3 |
| G97 | 184 | 2 | 34 | 26.5 | 67.7 | 35 | 11.4 | 68.6 | 32 | 21.9 | 53.1 |
| L59 | 184 | 3 | 4 | 50.0 | 25.0 | 8 | 37.5 | 62.5 | 3 | 66.7 | 33.3 |
| D15 | 182 | 2 | 31 | 16.1 | 71.0 | 38 | 21.0 | 63.2 | 40 | 25.0 | 75.0 |
| Q62 | 182 | 2 | 20 | 30.0 | 60.0 | 23 | 26.1 | 69.6 | 27 | 29.6 | 59.3 |
| D59 | 181 | 2 | 40 | 30.0 | 62.5 | 51 | 25.5 | 64.7 | 38 | 26.3 | 63.2 |
| H46 | 181 | 2 | 22 | 36.4 | 50.0 | 33 | 30.3 | 60.6 | 34 | 38.2 | 58.8 |
| N04 | 179 | 2 | 45 | 35.6 | 51.1 | 66 | 27.3 | 59.1 | 71 | 31.0 | 54.9 |
| Q04 | 179 | 2 | 7 | 57.1 | 42.9 | 11 | 45.5 | 54.5 | 7 | 71.4 | 28.6 |
| T83 | 177 | 3 | 42 | 38.1 | 35.7 | 34 | 35.3 | 61.8 | 25 | 36.0 | 60.0 |
| G98 | 177 | 2 | 10 | 20.0 | 80.0 | 20 | 25.0 | 75.0 | 14 | 42.9 | 57.1 |
| R90 | 176 | 2 | 12 | 25.0 | 75.0 | 16 | 12.5 | 81.2 | 8 | 25.0 | 75.0 |
| T30 | 176 | 3 | 10 | 30.0 | 60.0 | 10 | 40.0 | 60.0 | 9 | 44.4 | 55.6 |
| T28 | 176 | 2 | 15 | 13.3 | 86.7 | 25 | 24.0 | 72.0 | 16 | 25.0 | 75.0 |
| I07 | 173 | 2 | 24 | 12.5 | 75.0 | 32 | 6.2 | 78.1 | 33 | 6.1 | 87.9 |
| R46 | 173 | 2 | 4 | 50.0 | 25.0 | 9 | 22.2 | 33.3 | 2 | 100.0 | 0.0 |
| Q27 | 172 | 3 | 14 | 7.1 | 92.9 | 13 | 30.8 | 69.2 | 6 | 33.3 | 66.7 |
| N74 | 171 | 4 | 13 | 38.5 | 53.9 | 28 | 39.3 | 39.3 | 9 | 55.6 | 22.2 |
| F53 | 169 | 3 | 26 | 23.1 | 50.0 | 9 | 66.7 | 33.3 | 7 | 57.1 | 42.9 |
| Q85 | 166 | 5 | 5 | 20.0 | 80.0 | 6 | 33.3 | 66.7 | 13 | 38.5 | 53.9 |
| K37 | 163 | 3 | 26 | 46.1 | 53.9 | 30 | 23.3 | 73.3 | 22 | 31.8 | 68.2 |
| G11 | 162 | 2 | 8 | 12.5 | 87.5 | 22 | 13.6 | 68.2 | 17 | 23.5 | 70.6 |
| Q60 | 162 | 2 | 14 | 28.6 | 50.0 | 20 | 20.0 | 70.0 | 14 | 21.4 | 64.3 |
| C65 | 160 | 2 | 24 | 25.0 | 58.3 | 35 | 28.6 | 68.6 | 50 | 32.0 | 62.0 |
| M34 | 160 | 2 | 7 | 28.6 | 71.4 | 19 | 31.6 | 63.2 | 9 | 44.4 | 55.6 |
| B76 | 159 | 3 | 4 | 50.0 | 50.0 | 6 | 33.3 | 66.7 | 4 | 50.0 | 50.0 |
| C76 | 159 | 2 | 23 | 39.1 | 47.8 | 48 | 27.1 | 72.9 | 52 | 32.7 | 61.5 |
| N37 | 158 | 4 | 6 | 66.7 | 33.3 | 9 | 44.4 | 44.4 | 3 | 100.0 | 0.0 |
| C81 | 156 | 2 | 32 | 31.2 | 65.6 | 50 | 30.0 | 70.0 | 66 | 30.3 | 65.2 |
| I28 | 156 | 2 | 16 | 31.2 | 62.5 | 24 | 12.5 | 75.0 | 23 | 17.4 | 78.3 |
| A51 | 155 | 2 | 21 | 42.9 | 47.6 | 12 | 50.0 | 41.7 | 13 | 15.4 | 76.9 |
| F63 | 155 | 2 | 3 | 33.3 | 66.7 | 2 | 50.0 | 50.0 | 2 | 50.0 | 50.0 |
| K38 | 155 | 3 | 22 | 27.3 | 50.0 | 28 | 21.4 | 75.0 | 18 | 38.9 | 55.6 |
| A53 | 153 | 4 | 10 | 60.0 | 30.0 | 11 | 45.5 | 45.5 | 8 | 50.0 | 37.5 |
| S97 | 152 | 5 | 8 | 37.5 | 62.5 | 8 | 37.5 | 62.5 | 7 | 42.9 | 57.1 |
| M33 | 152 | 2 | 19 | 52.6 | 47.4 | 30 | 33.3 | 60.0 | 36 | 33.3 | 52.8 |
| K77 | 150 | 2 | 11 | 45.5 | 54.5 | 17 | 41.2 | 58.8 | 6 | 83.3 | 16.7 |
| M61 | 149 | 5 | 5 | 20.0 | 80.0 | 5 | 40.0 | 60.0 | 1 | 100.0 | 0.0 |
| S14 | 149 | 2 | 25 | 28.0 | 68.0 | 32 | 21.9 | 71.9 | 38 | 26.3 | 68.4 |
| R86 | 148 | 2 | 10 | 30.0 | 60.0 | 11 | 36.4 | 63.6 | 11 | 45.5 | 54.5 |
| S34 | 147 | 4 | 5 | 60.0 | 40.0 | 10 | 50.0 | 50.0 | 7 | 71.4 | 28.6 |
| Q74 | 147 | 3 | 3 | 33.3 | 66.7 | 4 | 25.0 | 75.0 | 2 | 50.0 | 50.0 |
| C24 | 146 | 2 | 40 | 32.5 | 42.5 | 57 | 31.6 | 64.9 | 68 | 27.9 | 69.1 |
| E75 | 145 | 5 | 7 | 28.6 | 71.4 | 11 | 36.4 | 63.6 | 3 | 33.3 | 66.7 |
| G60 | 145 | 5 | 3 | 66.7 | 33.3 | 3 | 33.3 | 66.7 | 5 | 80.0 | 20.0 |
| A28 | 144 | 5 | 6 | 33.3 | 66.7 | 10 | 20.0 | 80.0 | 4 | 25.0 | 75.0 |
| Q80 | 143 | 5 | 3 | 66.7 | 33.3 | 7 | 42.9 | 57.1 | 4 | 75.0 | 25.0 |
| T67 | 143 | 2 | 14 | 50.0 | 42.9 | 14 | 14.3 | 71.4 | 14 | 21.4 | 71.4 |
| E51 | 143 | 2 | 21 | 14.3 | 81.0 | 48 | 14.6 | 72.9 | 40 | 10.0 | 72.5 |
| G23 | 142 | 2 | 19 | 15.8 | 79.0 | 20 | 25.0 | 65.0 | 13 | 15.4 | 76.9 |
| M30 | 142 | 2 | 10 | 40.0 | 60.0 | 26 | 11.5 | 84.6 | 11 | 45.5 | 54.5 |
| C02 | 141 | 2 | 14 | 35.7 | 50.0 | 30 | 36.7 | 60.0 | 49 | 42.9 | 55.1 |
| A42 | 140 | 2 | 13 | 23.1 | 76.9 | 10 | 30.0 | 70.0 | 10 | 40.0 | 50.0 |
| D07 | 138 | 2 | 23 | 30.4 | 69.6 | 28 | 46.4 | 50.0 | 32 | 34.4 | 65.6 |
| O33 | 138 | 3 | 33 | 21.2 | 69.7 | 42 | 21.4 | 61.9 | 39 | 20.5 | 69.2 |
| M90 | 138 | 3 | 4 | 50.0 | 50.0 | 11 | 18.2 | 81.8 | 2 | 100.0 | 0.0 |
| B58 | 138 | 2 | 14 | 28.6 | 42.9 | 18 | 33.3 | 66.7 | 11 | 27.3 | 72.7 |
| L83 | 137 | 2 | 8 | 50.0 | 50.0 | 18 | 33.3 | 66.7 | 13 | 30.8 | 53.9 |
| R34 | 137 | 2 | 27 | 33.3 | 63.0 | 40 | 17.5 | 65.0 | 31 | 19.4 | 71.0 |
| K23 | 137 | 2 | 3 | 33.3 | 66.7 | 7 | 28.6 | 71.4 | 2 | 100.0 | 0.0 |
| Q68 | 137 | 4 | 2 | 50.0 | 50.0 | 6 | 50.0 | 50.0 | 3 | 66.7 | 33.3 |
| I52 | 137 | 4 | 6 | 50.0 | 50.0 | 12 | 33.3 | 66.7 | 5 | 80.0 | 20.0 |
| Q24 | 136 | 2 | 13 | 23.1 | 61.5 | 10 | 20.0 | 50.0 | 6 | 33.3 | 50.0 |
| J70 | 136 | 2 | 22 | 22.7 | 72.7 | 26 | 26.9 | 61.5 | 21 | 23.8 | 57.1 |
| A98 | 135 | 2 | 29 | 13.8 | 82.8 | 51 | 17.6 | 66.7 | 41 | 19.5 | 68.3 |
| M49 | 135 | 5 | 1 | 100.0 | 0.0 | 4 | 50.0 | 50.0 | 1 | 100.0 | 0.0 |
| R85 | 134 | 2 | 8 | 37.5 | 62.5 | 14 | 21.4 | 50.0 | 11 | 27.3 | 27.3 |
| C69 | 134 | 2 | 9 | 55.6 | 44.4 | 22 | 36.4 | 50.0 | 22 | 36.4 | 59.1 |
| T59 | 134 | 4 | 13 | 30.8 | 61.5 | 17 | 11.8 | 76.5 | 13 | 15.4 | 76.9 |
| T87 | 133 | 2 | 27 | 59.3 | 40.7 | 17 | 11.8 | 88.2 | 9 | 33.3 | 66.7 |
| S65 | 133 | 2 | 16 | 68.8 | 31.2 | 26 | 7.7 | 92.3 | 17 | 17.6 | 82.3 |
| D00 | 131 | 2 | 13 | 15.4 | 69.2 | 18 | 33.3 | 66.7 | 12 | 58.3 | 41.7 |
| Q83 | 130 | 4 | 3 | 66.7 | 33.3 | 8 | 25.0 | 75.0 | 6 | 50.0 | 50.0 |
| C17 | 130 | 2 | 21 | 23.8 | 52.4 | 41 | 26.8 | 73.2 | 42 | 26.2 | 71.4 |
| C62 | 129 | 2 | 12 | 33.3 | 66.7 | 27 | 33.3 | 66.7 | 30 | 36.7 | 63.3 |
| G09 | 129 | 3 | 7 | 28.6 | 71.4 | 4 | 75.0 | 25.0 | 3 | 66.7 | 33.3 |
| E64 | 129 | 2 | 3 | 33.3 | 66.7 | 6 | 33.3 | 66.7 | 2 | 100.0 | 0.0 |
| B71 | 128 | 5 | 7 | 42.9 | 57.1 | 7 | 57.1 | 42.9 | 4 | 100.0 | 0.0 |
| Q12 | 127 | 5 | 5 | 80.0 | 20.0 | 7 | 71.4 | 28.6 | 6 | 50.0 | 33.3 |
| S54 | 126 | 2 | 15 | 40.0 | 60.0 | 18 | 27.8 | 72.2 | 14 | 28.6 | 64.3 |
| G00 | 125 | 2 | 37 | 32.4 | 54.0 | 54 | 24.1 | 53.7 | 53 | 22.6 | 60.4 |
| I23 | 125 | 2 | 33 | 36.4 | 54.5 | 55 | 25.4 | 61.8 | 60 | 26.7 | 63.3 |
| R89 | 124 | 2 | 3 | 66.7 | 33.3 | 8 | 50.0 | 50.0 | 2 | 100.0 | 0.0 |
| G03 | 124 | 2 | 30 | 26.7 | 60.0 | 43 | 27.9 | 62.8 | 37 | 35.1 | 51.4 |
| T62 | 123 | 2 | 15 | 60.0 | 40.0 | 21 | 14.3 | 81.0 | 17 | 11.8 | 88.2 |
| F04 | 122 | 4 | 17 | 17.6 | 70.6 | 22 | 18.2 | 81.8 | 14 | 21.4 | 78.6 |
| E63 | 121 | 3 | 3 | 50.0 | 50.0 | 12 | 8.3 | 91.7 | 4 | 25.0 | 75.0 |
| D42 | 120 | 2 | 13 | 38.5 | 53.9 | 22 | 18.2 | 68.2 | 17 | 47.1 | 52.9 |
| E24 | 119 | 2 | 17 | 35.3 | 64.7 | 16 | 12.5 | 87.5 | 11 | 18.2 | 72.7 |
| C96 | 117 | 2 | 36 | 36.1 | 61.1 | 43 | 37.2 | 58.1 | 54 | 33.3 | 64.8 |
| D06 | 116 | 2 | 16 | 31.2 | 56.2 | 30 | 40.0 | 46.7 | 35 | 40.0 | 45.7 |
| B50 | 116 | 4 | 4 | 50.0 | 50.0 | 5 | 60.0 | 40.0 | 3 | 100.0 | 0.0 |
| Q52 | 115 | 5 | 3 | 66.7 | 33.3 | 6 | 50.0 | 50.0 | 5 | 60.0 | 40.0 |
| Q64 | 115 | 2 | 11 | 27.3 | 63.6 | 9 | 33.3 | 66.7 | 9 | 33.3 | 55.6 |
| S16 | 114 | 5 | 6 | 33.3 | 66.7 | 7 | 28.6 | 71.4 | 6 | 16.7 | 83.3 |
| C10 | 114 | 2 | 23 | 52.2 | 47.8 | 27 | 44.4 | 55.6 | 45 | 42.2 | 55.6 |
| Q78 | 114 | 5 | 3 | 66.7 | 33.3 | 6 | 33.3 | 66.7 | 3 | 66.7 | 33.3 |
| D05 | 113 | 2 | 20 | 35.0 | 45.0 | 22 | 54.5 | 45.5 | 28 | 46.4 | 42.9 |
| O16 | 113 | 5 | 25 | 52.0 | 36.0 | 34 | 17.6 | 64.7 | 34 | 20.6 | 55.9 |
| E85 | 112 | 2 | 33 | 21.2 | 66.7 | 43 | 25.6 | 60.5 | 46 | 28.3 | 58.7 |
| C57 | 112 | 2 | 18 | 22.2 | 72.2 | 46 | 26.1 | 67.4 | 54 | 34.0 | 62.3 |
| B15 | 111 | 2 | 18 | 11.1 | 77.8 | 28 | 17.9 | 64.3 | 23 | 17.4 | 65.2 |
| F14 | 110 | 2 | 13 | 23.1 | 69.2 | 15 | 33.3 | 60.0 | 16 | 25.0 | 75.0 |
| C95 | 110 | 2 | 17 | 17.6 | 76.5 | 52 | 25.0 | 69.2 | 56 | 32.1 | 57.1 |
| Q72 | 109 | 5 | 3 | 66.7 | 33.3 | 6 | 50.0 | 50.0 | 3 | 66.7 | 33.3 |
| N05 | 109 | 2 | 8 | 50.0 | 50.0 | 32 | 21.9 | 75.0 | 25 | 24.0 | 72.0 |
| C51 | 108 | 2 | 15 | 40.0 | 60.0 | 27 | 40.7 | 59.3 | 39 | 48.7 | 51.3 |
| N25 | 107 | 5 | 7 | 42.9 | 57.1 | 16 | 37.5 | 56.2 | 6 | 16.7 | 83.3 |
| T34 | 106 | 3 | 15 | 20.0 | 73.3 | 16 | 12.5 | 75.0 | 25 | 16.0 | 72.0 |
| S85 | 106 | 5 | 6 | 16.7 | 83.3 | 10 | 20.0 | 80.0 | 5 | 40.0 | 60.0 |
| S84 | 105 | 4 | 9 | 33.3 | 66.7 | 10 | 30.0 | 70.0 | 8 | 37.5 | 62.5 |
| F73 | 105 | 2 | 2 | 100.0 | 0.0 | 4 | 50.0 | 50.0 | 2 | 50.0 | 50.0 |
| N82 | 103 | 2 | 11 | 18.2 | 54.5 | 17 | 35.3 | 58.8 | 10 | 50.0 | 50.0 |
| T54 | 103 | 4 | 7 | 100.0 | 0.0 | 9 | 33.3 | 66.7 | 10 | 90.0 | 10.0 |
| Q99 | 102 | 2 | 1 | 100.0 | 0.0 | 2 | 50.0 | 50.0 | 1 | 100.0 | 0.0 |
| Q79 | 100 | 5 | 1 | 100.0 | 0.0 | 4 | 50.0 | 50.0 | 4 | 50.0 | 50.0 |
