## Appendix 6 for "A national-scale digital atlas of recorded care patterns across more than 1000 diagnostic categories using real-world health data: a retrospective observational study"

**Appendix 6: Characterization of condition–concept pair relationship classifications**

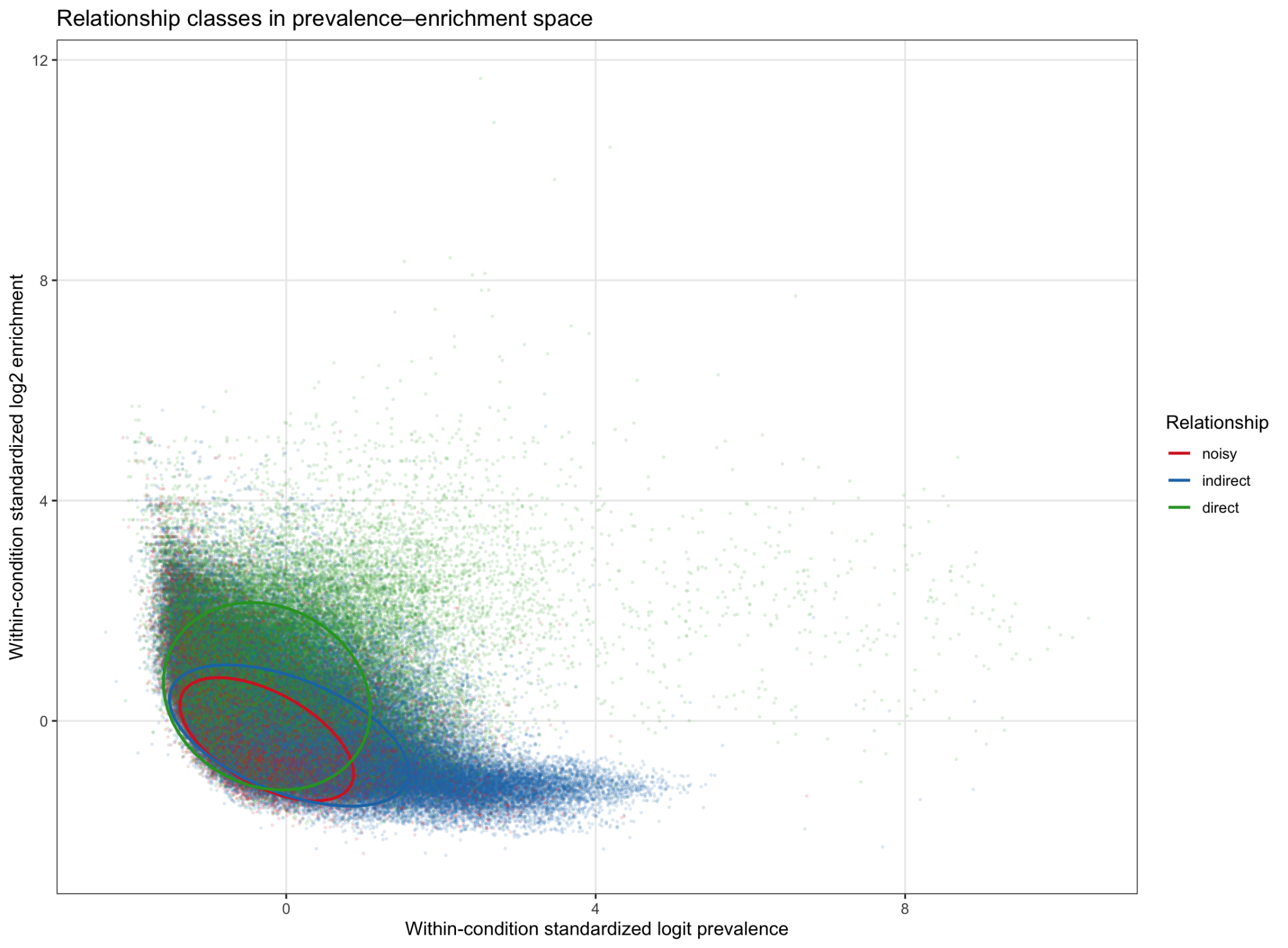

**Figure S1.** Relationship classes in within-condition prevalence–enrichment space

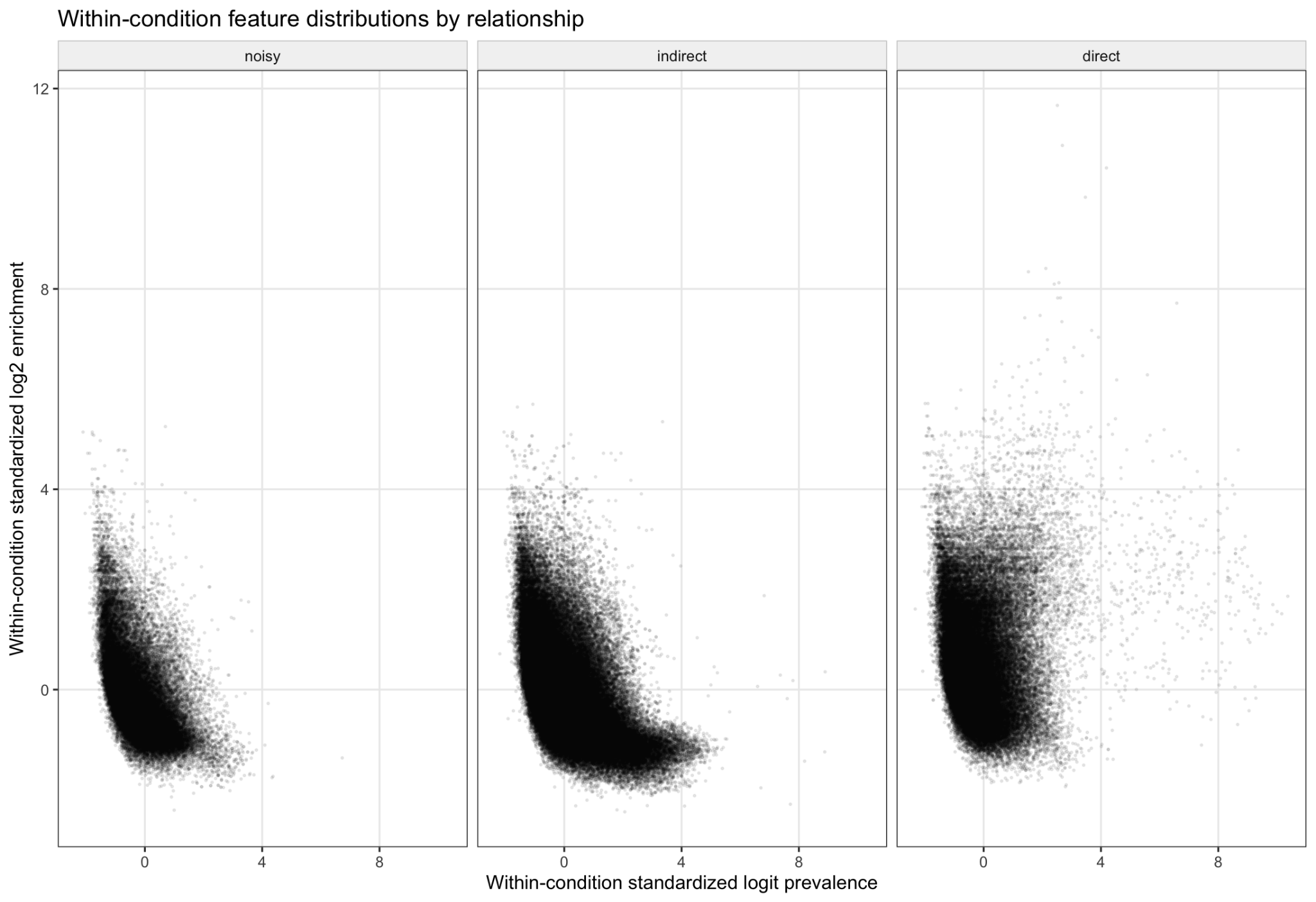

**Figure S2.** Class-specific distributions of within-condition standardized prevalence and enrichment

**Table S4.** Concept prevalence and enrichment distribution with regards to relationship

| **Group** | **Relationship** | **Prev Q25** | **Prev Med** | **Prev Q75** | **Enr Q25** | **Enr Med** | **Enr Q75** |
| --- | --- | --- | --- | --- | --- | --- | --- |
| PRE90 | Noisy | 0.011 | 0.031 | 0.091 | 1.775 | 2.738 | 5.378 |
| PRE90 | Indirect | 0.028 | 0.096 | 0.319 | 1.811 | 3.006 | 5.927 |
| PRE90 | Direct | 0.012 | 0.040 | 0.134 | 3.105 | 6.124 | 16.063 |
| POST30 | Noisy | 0.007 | 0.019 | 0.061 | 2.651 | 5.101 | 12.386 |
| POST30 | Indirect | 0.013 | 0.047 | 0.178 | 2.818 | 6.012 | 15.933 |
| POST30 | Direct | 0.012 | 0.044 | 0.165 | 7.205 | 19.365 | 100.000 |
| POST365 | Noisy | 0.012 | 0.039 | 0.113 | 1.476 | 1.993 | 3.446 |
| POST365 | Indirect | 0.025 | 0.090 | 0.284 | 1.564 | 2.382 | 4.539 |
| POST365 | Direct | 0.019 | 0.070 | 0.230 | 2.945 | 5.888 | 20.258 |
