## Appendix 7 for "A national-scale digital atlas of recorded care patterns across more than 1000 diagnostic categories using real-world health data: a retrospective observational study"

**Appendix 7: Top separators of the treatment patterns across observation periods**

**Table S5.** Top separators of the treatment patterns across observation periods

| **Rank** | **Concept name** | **Windows** | **Top-10 count** | **Mean rank** |
| --- | --- | --- | --- | --- |
| 1 | Immunology laboratory test | 3 | 1,516 | 4.53 |
| 2 | Enzyme measurement | 3 | 1,457 | 4.23 |
| 3 | Glucose measurement | 3 | 1,447 | 5.05 |
| 4 | Electrolytes measurement | 3 | 1,333 | 5.23 |
| 5 | CBC W Auto Differential panel - Blood | 3 | 1,222 | 4.41 |
| 6 | C-reactive protein measurement | 3 | 1,188 | 4.71 |
| 7 | Electrocardiographic monitoring | 3 | 1,107 | 6.20 |
| 8 | Emergency Room and Inpatient Visit (visit_occurrence) | 3 | 836 | 3.85 |
| 9 | Emergency Room and Inpatient Visit | 3 | 833 | 4.00 |
| 10 | Bilirubin measurement | 3 | 743 | 6.81 |
| 11 | Inpatient Visit | 3 | 716 | 4.72 |
| 12 | Emergency Room Visit (visit_occurrence) | 3 | 644 | 4.14 |
| 13 | Emergency Room Visit | 3 | 627 | 4.23 |
| 14 | Test strip urinalysis | 3 | 600 | 7.37 |
| 15 | Coagulation pathway screening | 3 | 560 | 6.16 |
| 16 | Inpatient Visit (visit_occurrence) | 3 | 546 | 4.76 |
| 17 | Hemoglobin A1c measurement | 3 | 530 | 7.95 |
| 18 | Admission by general nurse | 3 | 491 | 7.63 |
| 19 | Admission by physician | 3 | 487 | 6.88 |
| 20 | Albumin measurement | 3 | 384 | 6.35 |
| 21 | Telephone consultation | 3 | 348 | 7.66 |
| 22 | Procalcitonin [Mass/volume] in Serum or Plasma | 3 | 309 | 5.56 |
| 23 | Follow-up visit | 3 | 309 | 7.49 |
| 24 | Evaluation of acid-base balance | 3 | 308 | 5.45 |
| 25 | Blood group antibody screen.cells I+II+III [Presence] in Serum or Plasma | 3 | 294 | 6.03 |
| 26 | Standard level emergency care | 3 | 249 | 5.72 |
| 27 | Aerobic microbial culture | 3 | 243 | 6.23 |
| 28 | Anesthesia duration | 3 | 231 | 3.46 |
| 29 | Anesthesia duration + Recovery room monitoring, anesthesia | 3 | 222 | 4.13 |
| 30 | Fibrin-fibrinogen split products assay | 3 | 207 | 7.36 |
| 31 | Emergency procedure | 3 | 205 | 6.62 |
| 32 | Planned procedure | 3 | 195 | 5.94 |
| 33 | CT of brain without contrast + Radiology of two body areas | 3 | 170 | 5.16 |
| 34 | HIV 1+2 Ab+HIV1 p24 Ag [Presence] in Serum or Plasma by Immunoassay | 3 | 145 | 6.11 |
| 35 | Intensive Care | 3 | 138 | 4.82 |
| 36 | CT with contrast + CT without contrast + Radiology of two body areas | 3 | 138 | 5.97 |
| 37 | Chromosome analysis, cytogenetic procedure AND/OR molecular biology method | 3 | 132 | 4.72 |
| 38 | Erythrocyte sedimentation rate measurement | 3 | 130 | 7.48 |
| 39 | Plain X-ray of chest | 3 | 123 | 7.59 |
| 40 | Polymerase chain reaction analysis | 3 | 122 | 4.58 |
