## Appendix 8 for "A national-scale digital atlas of recorded care patterns across more than 1000 diagnostic categories using real-world health data: a retrospective observational study"

**Appendix 8: Clustering ICD-10 subchapters over all the results**

**Figure S3.** Top 20 subchapters (n>3) with the highest silhouette widths over all diseases for the window of POST365

**Figure S4.** Cross-disease embedding of diagnosis-window summaries by ICD-10 chapter and subchapter for PRE90 view. Each point represents an eligible ICD-10 three-character diagnosis category embedded using retained-concept presence and frequency, together with cohort-level median age and male proportion. Colours indicate ICD-10 chapters and point shapes indicate ICD-10 subchapters. The two-dimensional projection was generated using UMAP for visual exploration of whether diagnoses with similar retained care-pattern summaries formed local groupings. Axis orientation and absolute distances are arbitrary; local proximity should be interpreted as approximate similarity in the atlas-derived feature space.

**Figure S5.** Top 20 subchapters (n>3) with the highest silhouette widths over all diseases for the window of PRE90

**Figure S6.** Cross-disease embedding of diagnosis-window summaries by ICD-10 chapter and subchapter for POST30 view. Each point represents an eligible ICD-10 three-character diagnosis category embedded using retained-concept presence and frequency, together with cohort-level median age and male proportion. Colours indicate ICD-10 chapters and point shapes indicate ICD-10 subchapters. The two-dimensional projection was generated using UMAP for visual exploration of whether diagnoses with similar retained care-pattern summaries formed local groupings. Axis orientation and absolute distances are arbitrary; local proximity should be interpreted as approximate similarity in the atlas-derived feature space.

**Figure S7.** Top 20 subchapters (n>3) with the highest silhouette widths over all diseases for the window of POST30

**Table S6.** ICD subchapters (n>3) ranked by mean silhouette width

| **Rank** | **ICD range** | **Subchapter** | **N (conditions)** | **POST365** | **POST30** | **PRE90** |
| --- | --- | --- | --- | --- | --- | --- |
| 1 | T20-T25 | Burns and corrosions of external body surface, specified by site | 6 | 0.844 | 0.893 | 0.834 |
| 2 | K80-K87 | Disorders of gallbladder, biliary tract and pancreas | 6 | 0.766 | 0.753 | 0.822 |
| 3 | O00-O08 | Pregnancy with abortive outcome | 6 | 0.740 | 0.736 | 0.718 |
| 4 | N60-N64 | Disorders of breast | 5 | 0.790 | 0.755 | 0.342 |
| 5 | N40-N51 | Diseases of male genital organs | 12 | 0.172 | 0.481 | 0.486 |
| 6 | S60-S69 | Injuries to the wrist, hand and fingers | 10 | -0.108 | 0.679 | 0.533 |
| 7 | J00-J06 | Acute upper respiratory infections | 7 | 0.799 | 0.551 | -0.324 |
| 8 | O10-O16 | Edema, proteinuria and hypertensive disorders in pregnancy, childbirth and the puerperium | 5 | 0.408 | 0.243 | 0.312 |
| 9 | B65-B83 | Helminthiases | 8 | 0.455 | 0.659 | -0.332 |
| 10 | J90-J94 | Other diseases of the pleura | 4 | 0.496 | 0.141 | 0.097 |
| 11 | C81-C96 | Malignant neoplasms of lymphoid, hematopoietic and related tissue | 8 | 0.249 | 0.557 | -0.135 |
| 12 | K00-K14 | Diseases of oral cavity and salivary glands | 13 | 0.426 | 0.564 | -0.330 |
| 13 | O60-O75 | Complications of labor and delivery | 14 | 0.156 | 0.292 | 0.089 |
| 14 | O85-O92 | Complications predominantly related to the puerperium | 6 | 0.156 | -0.394 | 0.755 |
| 15 | H00-H06 | Disorders of eyelid, lacrimal system and orbit | 5 | 0.330 | 0.073 | 0.022 |
| 16 | F40-F48 | Anxiety, dissociative, stress-related, somatoform and other nonpsychotic mental disorders | 7 | 0.248 | 0.199 | -0.091 |
| 17 | C15-C26 | Malignant neoplasms of digestive organs | 11 | 0.662 | 0.441 | -0.759 |
| 18 | L10-L14 | Bullous disorders | 4 | 0.439 | 0.236 | -0.388 |
| 19 | M20-M25 | Other joint disorders | 6 | -0.057 | 0.346 | -0.044 |
| 20 | O20-O29 | Other maternal disorders predominantly related to pregnancy | 8 | 0.175 | 0.063 | -0.084 |
| 21 | Q65-Q79 | Congenital malformations and deformations of the musculoskeletal system | 9 | -0.232 | 0.252 | 0.077 |
| 22 | L20-L30 | Dermatitis and eczema | 10 | 0.360 | -0.523 | 0.249 |
| 23 | H30-H36 | Disorders of choroid and retina | 6 | 0.118 | -0.078 | -0.151 |
| 24 | M50-M54 | Other dorsopathies | 4 | -0.779 | 0.377 | 0.276 |
| 25 | K35-K38 | Diseases of appendix | 4 | -0.102 | 0.584 | -0.718 |
| 26 | F90-F98 | Behavioral and emotional disorders with onset usually occurring in childhood and adolescence | 6 | -0.426 | -0.123 | 0.306 |
| 27 | H15-H22 | Disorders of sclera, cornea, iris and ciliary body | 6 | -0.339 | -0.017 | 0.044 |
| 28 | H65-H75 | Diseases of middle ear and mastoid | 10 | -0.354 | 0.488 | -0.538 |
| 29 | I10-I15 | Hypertensive diseases | 5 | -0.429 | -0.094 | 0.095 |
| 30 | M15-M19 | Osteoarthritis | 5 | 0.176 | -0.768 | 0.132 |
| 31 | B00-B09 | Viral infections characterized by skin and mucous membrane lesions | 6 | -0.167 | -0.262 | -0.077 |
| 32 | T36-T50 | Poisoning by, adverse effects of and underdosing of drugs, medicaments and biological substances | 7 | -0.002 | -0.194 | -0.329 |
| 33 | N80-N98 | Noninflammatory disorders of female genital tract | 19 | -0.115 | -0.296 | -0.188 |
| 34 | H49-H52 | Disorders of ocular muscles, binocular movement, accommodation and refraction | 4 | -0.145 | -0.323 | -0.184 |
| 35 | J40-J47 | Chronic lower respiratory diseases | 7 | -0.341 | -0.679 | 0.368 |
| 36 | F20-F29 | Schizophrenia, schizotypal, delusional, and other non-mood psychotic disorders | 7 | -0.058 | -0.159 | -0.482 |
| 37 | D50-D53 | Nutritional anemias | 4 | -0.374 | 0.113 | -0.465 |
| 38 | G20-G26 | Extrapyramidal and movement disorders | 5 | -0.460 | 0.344 | -0.641 |
| 39 | H25-H28 | Disorders of lens | 4 | -0.348 | -0.167 | -0.321 |
| 40 | M45-M49 | Spondylopathies | 4 | -0.166 | -0.717 | 0.015 |
| 41 | N70-N77 | Inflammatory diseases of female pelvic organs | 8 | -0.540 | -0.190 | -0.257 |
| 42 | B15-B19 | Viral hepatitis | 4 | -0.651 | 0.373 | -0.731 |
| 43 | I60-I69 | Cerebrovascular diseases | 9 | -0.157 | -0.286 | -0.581 |
| 44 | G40-G47 | Episodic and paroxysmal disorders | 6 | -0.446 | -0.219 | -0.416 |
| 45 | D60-D64 | Aplastic and other anemias and other bone marrow failure syndromes | 4 | -0.261 | -0.627 | -0.196 |
| 46 | F30-F39 | Mood [affective] disorders | 6 | -0.377 | -0.272 | -0.439 |
| 47 | D70-D77 | Other disorders of blood and blood-forming organs | 4 | -0.412 | -0.196 | -0.481 |
| 48 | N10-N16 | Renal tubulo-interstitial diseases | 5 | -0.477 | -0.126 | -0.496 |
| 49 | Q60-Q64 | Congenital malformations of the urinary system | 5 | -0.561 | -0.737 | 0.183 |
| 50 | M40-M43 | Deforming dorsopathies | 4 | -0.026 | -0.245 | -0.866 |
| 51 | O30-O48 | Maternal care related to the fetus and amniotic cavity and possible delivery problems | 15 | -0.400 | -0.365 | -0.427 |
| 52 | C51-C58 | Malignant neoplasms of female genital organs | 5 | 0.039 | -0.446 | -0.790 |
| 53 | I05-I09 | Chronic rheumatic heart diseases | 5 | -0.713 | -0.523 | -0.004 |
| 54 | S40-S49 | Injuries to the shoulder and upper arm | 7 | -0.506 | -0.489 | -0.254 |
| 55 | E00-E07 | Disorders of thyroid gland | 7 | -0.499 | -0.505 | -0.272 |
| 56 | J30-J39 | Other diseases of upper respiratory tract | 10 | -0.757 | -0.447 | -0.129 |
| 57 | N00-N08 | Glomerular diseases | 6 | -0.187 | -0.534 | -0.682 |
| 58 | L40-L45 | Papulosquamous disorders | 5 | -0.604 | -0.225 | -0.581 |
| 59 | S10-S19 | Injuries to the neck | 7 | -0.576 | -0.537 | -0.332 |
| 60 | J09-J18 | Influenza and pneumonia | 9 | -0.414 | -0.669 | -0.386 |
| 61 | L50-L54 | Urticaria and erythema | 4 | -0.510 | -0.424 | -0.535 |
| 62 | L60-L75 | Disorders of skin appendages | 11 | -0.540 | -0.494 | -0.442 |
| 63 | H80-H83 | Diseases of inner ear | 4 | -0.749 | -0.885 | 0.143 |
| 64 | R30-R39 | Symptoms and signs involving the genitourinary system | 6 | -0.275 | -0.527 | -0.695 |
| 65 | Q10-Q18 | Congenital malformations of eye, ear, face and neck | 4 | -0.358 | -0.592 | -0.548 |
| 66 | Q80-Q89 | Other congenital malformations | 5 | -0.569 | -0.454 | -0.495 |
| 67 | I20-I25 | Ischemic heart diseases | 6 | -0.676 | -0.482 | -0.361 |
| 68 | S50-S59 | Injuries to the elbow and forearm | 7 | -0.408 | -0.610 | -0.523 |
| 69 | K55-K64 | Other diseases of intestines | 9 | -0.383 | -0.549 | -0.635 |
| 70 | L00-L08 | Infections of the skin and subcutaneous tissue | 7 | -0.409 | -0.590 | -0.602 |
| 71 | B85-B89 | Pediculosis, acariasis and other infestations | 4 | -0.635 | -0.597 | -0.378 |
| 72 | S00-S09 | Injuries to the head | 7 | -0.536 | -0.541 | -0.541 |
| 73 | S90-S99 | Injuries to the ankle and foot | 7 | -0.489 | -0.605 | -0.554 |
| 74 | S20-S29 | Injuries to the thorax | 6 | -0.521 | -0.516 | -0.646 |
| 75 | H90-H95 | Other disorders of ear and mastoid process | 5 | -0.494 | -0.615 | -0.612 |
| 76 | I30-I52 | Other forms of heart disease | 19 | -0.650 | -0.498 | -0.599 |
| 77 | L80-L99 | Other disorders of the skin and subcutaneous tissue | 18 | -0.592 | -0.563 | -0.600 |
| 78 | L55-L59 | Radiation-related disorders of the skin and subcutaneous tissue | 4 | -0.544 | -0.627 | -0.592 |
| 79 | G00-G09 | Inflammatory diseases of the central nervous system | 4 | -0.648 | -0.535 | -0.592 |
| 80 | G90-G99 | Other disorders of the nervous system | 9 | -0.653 | -0.440 | -0.688 |
| 81 | F10-F19 | Mental and behavioral disorders due to psychoactive substance use | 8 | -0.562 | -0.524 | -0.710 |
| 82 | Z80-Z99 | Persons with potential health hazards related to family and personal history and certain conditions influencing health status | 4 | -0.700 | -0.611 | -0.489 |
| 83 | I70-I79 | Diseases of arteries, arterioles and capillaries | 8 | -0.622 | -0.523 | -0.671 |
| 84 | T51-T65 | Toxic effects of substances chiefly nonmedicinal as to source | 7 | -0.733 | -0.650 | -0.493 |
| 85 | R90-R94 | Abnormal findings on diagnostic imaging and in function studies, without diagnosis | 4 | -0.716 | -0.418 | -0.756 |
| 86 | S80-S89 | Injuries to the knee and lower leg | 8 | -0.553 | -0.751 | -0.594 |
| 87 | F80-F89 | Pervasive and specific developmental disorders | 5 | -0.610 | -0.628 | -0.670 |
| 88 | K40-K46 | Hernia | 7 | -0.429 | -0.700 | -0.789 |
| 89 | Q20-Q28 | Congenital malformations of the circulatory system | 6 | -0.719 | -0.664 | -0.547 |
| 90 | R10-R19 | Symptoms and signs involving the digestive system and abdomen | 10 | -0.566 | -0.665 | -0.699 |
| 91 | I80-I89 | Diseases of veins, lymphatic vessels and lymph nodes, not elsewhere classified | 9 | -0.647 | -0.734 | -0.568 |
| 92 | S70-S79 | Injuries to the hip and thigh | 6 | -0.610 | -0.614 | -0.737 |
| 93 | S30-S39 | Injuries to the abdomen, lower back, lumbar spine, pelvis and external genitals | 8 | -0.640 | -0.693 | -0.654 |
| 94 | R20-R23 | Symptoms and signs involving the skin and subcutaneous tissue | 4 | -0.665 | -0.674 | -0.680 |
| 95 | T15-T19 | Effects of foreign body entering through natural orifice | 5 | -0.595 | -0.703 | -0.723 |
| 96 | D37-D48 | Neoplasms of uncertain behavior, polycythemia vera and myelodysplastic syndromes | 12 | -0.783 | -0.730 | -0.571 |
| 97 | F00-F09 | Organic, including symptomatic, mental disorders | 8 | -0.765 | -0.690 | -0.633 |
| 98 | M30-M36 | Systemic connective tissue disorders | 6 | -0.728 | -0.553 | -0.809 |
| 99 | A30-A49 | Other bacterial diseases | 9 | -0.707 | -0.676 | -0.724 |
| 100 | N30-N39 | Other diseases of the urinary system | 8 | -0.706 | -0.740 | -0.671 |
| 101 | D00-D09 | In situ neoplasms | 7 | -0.728 | -0.704 | -0.707 |
| 102 | D10-D36 | Benign neoplasms, except benign neuroendocrine tumors | 25 | -0.687 | -0.754 | -0.718 |
| 103 | F70-F79 | Intellectual disabilities | 5 | -0.675 | -0.774 | -0.727 |
| 104 | G80-G83 | Cerebral palsy and other paralytic syndromes | 4 | -0.657 | -0.778 | -0.745 |
| 105 | K20-K31 | Diseases of esophagus, stomach and duodenum | 11 | -0.663 | -0.721 | -0.799 |
| 106 | A50-A64 | Infections with a predominantly sexual mode of transmission | 8 | -0.796 | -0.742 | -0.647 |
| 107 | R50-R69 | General symptoms and signs | 14 | -0.705 | -0.746 | -0.736 |
| 108 | M80-M94 | Osteopathies and chondropathies | 12 | -0.700 | -0.725 | -0.767 |
| 109 | K70-K77 | Diseases of liver | 8 | -0.743 | -0.717 | -0.734 |
| 110 | T66-T78 | Other and unspecified effects of external causes | 4 | -0.752 | -0.720 | -0.744 |
| 111 | G50-G59 | Nerve, nerve root and plexus disorders | 10 | -0.771 | -0.671 | -0.797 |
| 112 | T80-T88 | Complications of surgical and medical care, not elsewhere classified | 9 | -0.815 | -0.700 | -0.752 |
| 113 | G60-G64 | Polyneuropathies and other disorders of the peripheral nervous system | 5 | -0.789 | -0.768 | -0.713 |
| 114 | R40-R46 | Symptoms and signs involving cognition, perception, emotional state and behavior | 6 | -0.669 | -0.840 | -0.772 |
| 115 | B35-B49 | Mycoses | 5 | -0.801 | -0.722 | -0.756 |
| 116 | E20-E35 | Disorders of other endocrine glands | 9 | -0.723 | -0.798 | -0.767 |
| 117 | N25-N29 | Other disorders of kidney and ureter | 4 | -0.745 | -0.732 | -0.819 |
| 118 | E70-E90 | Metabolic disorders | 13 | -0.746 | -0.754 | -0.804 |
| 119 | Q50-Q56 | Congenital malformations of genital organs | 4 | -0.758 | -0.777 | -0.772 |
| 120 | M05-M14 | Inflammatory polyarthropathies | 9 | -0.825 | -0.642 | -0.839 |
| 121 | M60-M79 | Soft tissue disorders | 14 | -0.714 | -0.781 | -0.818 |
| 122 | R83-R89 | Abnormal findings on examination of other body fluids, substances and tissues, without diagnosis | 4 | -0.812 | -0.799 | -0.701 |
| 123 | R00-R09 | Symptoms and signs involving the circulatory and respiratory systems | 8 | -0.787 | -0.784 | -0.746 |
| 124 | E50-E64 | Other nutritional deficiencies | 7 | -0.728 | -0.793 | -0.827 |
| 125 | A00-A09 | Intestinal infectious diseases | 8 | -0.788 | -0.878 | -0.687 |
| 126 | B25-B34 | Other viral diseases | 5 | -0.875 | -0.849 | -0.733 |
| 127 | F50-F59 | Behavioral syndromes associated with physiological disturbances and physical factors | 4 | -0.884 | -0.860 | -0.779 |
